# Resolving inflammatory bowel disease risk variants to genes and cell types

**DOI:** 10.64898/2026.05.13.26352926

**Authors:** Laura Fachal, Rui Zhang, Kyle Gettler, Talin Haritunians, Isabelle Cleynen, Christine R. Stevens, Qian Zhang, Christopher Tastad, Catalina Medici, Ron Do, Maria T. Abreu, Jean-Paul Achkar, Tariq Ahmad, Klaartje Bel Kok, Charles Bernstein, Johanne Brooks, Luis Bujanda, Jeffrey Butterworth, Katie Clark, Fraser Cummings, Mauro D’Amato, Joanne Del Buono, Richard H. Duerr, David Ellinghaus, Stephen Foley, Denis Franchimont, Andre Franke, Laura Hancock, Ailsa Hart, Patricia Hooper, Peter Irving, Mark Jarvis, Emma Johnston, Antonio Julià, Cheryl Kemp, Nick Kennedy, Juozas Kupcinskas, Anna Latiano, James Lewis, Andy Li, Jimmy Limdi, Edouard Louis, John McLaughlin, Paul Moayyedi, Gordon Moran, Craig Mowat, Rodney D. Newberry, Bill Newman, Arabis Oglesby, Joel Pekow, Kate J. Perez, Richard Pollok, Natalie Prescott, Graham Radford-Smith, Laura Raffals, Tim Raine, Arvind Ramadas, Subramaniam Ramakrishnan, Walter Reinisch, Gerhard Rogler, Bruce E. Sands, Jack Satsangi, Michael Scharl, Stefan Schreiber, Shaji Sebastian, Christian Selinger, Sophy Shedwell, Mark S. Silverberg, Salil Singh, Helen Steed, Alan Steel, Holm H. Uhlig, Ajay Verma, Séverine Vermeire, Rinse K. Weersma, Ramnik Xavier, Jonas Halfvarson, Mark J. Daly, Christopher A. Lamb, Miles Parkes, Hailiang Huang, James C. Lee, Dermot P.B. McGovern, John D. Rioux, Carl A. Anderson, Judy H. Cho

## Abstract

Inflammatory bowel diseases (IBD), principally Crohn’s disease (CD) and ulcerative colitis (UC), are common chronic disorders involving inflammation and often progressive tissue damage. Genome-wide association studies have mapped many risk signals, but the causal variants, effector genes and relevant cellular contexts remain difficult to resolve, limiting mechanistic interpretation and therapeutic translation. Here we performed a multi-ancestry GWAS meta-analysis of 125,992 individuals with IBD and more than 1.2 million controls, identifying 619 independent association signals (374 novel) at 420 IBD regions that account for 77–80% of SNP-based heritability. Fine-mapping resolved 81 high-confidence variants, 41 not previously reported. Although most signals were shared between CD and UC, 39% showed IBD subtype specificity, with UC signals showing stronger enrichment in functional annotations from intestinal epithelial, secretory and enteroendocrine cells, and CD showing stronger genetic correlations with circulating inflammatory biomarkers, including C-reactive protein and glycoprotein acetylation. Latent causal modelling supported a causal effect of decreased high-density lipoprotein on CD risk. By integrating bulk and single-cell eQTL and pQTL resources using colocalisation and Mendelian randomisation, together with coding-variant evidence from exome sequencing, we prioritised 664 candidate effector genes across 341 signals, including 390 newly implicated IBD genes, revealing new biological mechanisms and candidate therapeutic targets supported by human genetics.

---

Inflammatory bowel disease (IBD) comprises two major chronic inflammatory disorders of the gastrointestinal tract: Crohn’s disease (CD) and ulcerative colitis (UC). CD can affect any part of the gut and is characterised by transmural inflammation, often leading to stricturing and fistulising complications, whereas UC is confined to the colon and typically limited to the mucosa. IBD is increasingly prevalent worldwide and imposes a lifelong burden on patients and healthcare systems^1^.

IBD susceptibility arises from the interplay of environmental exposures, stochastic factors and inherited risk. Rare, high-penetrance variants can cause severe, monogenic forms of IBD, but the majority of cases reflect a polygenic architecture^2–4^. Genome-wide association and sequencing studies have mapped more than 320 IBD risk signals, most driven by non-coding variation^5–8^. However, these signals account for only part of inherited susceptibility. Larger and more diverse genetic studies are needed both to capture remaining genetic risk and to increase power to fine-map causal variants, resolve effector genes and pinpoint the cellular contexts in which disease-associated variants act. These steps are crucial for mechanistic insight and therapeutic prioritisation. Although newly identified variants are expected to explain progressively smaller fractions of disease risk, genetic effect size is a poor predictor of the biological and therapeutic value of the genes and pathways they implicate^9^.

Here we conducted the largest multi-ancestry GWAS meta-analysis of IBD to date, comprising genetic data from more than 1.3 million individuals. We integrated these association results with bulk and single-cell expression QTL datasets, plasma protein QTL resources and coding-variant evidence from exome sequencing to refine likely causal variants, prioritise candidate effector genes and resolve the cell types, pathways and candidate therapeutic targets implicated by genetic risk for IBD.

## GWAS meta-analysis

We performed a multi-ancestry GWAS meta-analysis across 70 cohorts from European (EUR), East Asian (EAS) and South Asian (SAS) ancestry groups (PCA-inferred ancestry; Methods and Supplementary Table 1). The final dataset included 125,992 individuals with IBD (59,734 CD; 57,565 UC; 8,693 IBD-unclassified, included only in the combined IBD analyses) and 1,262,229 controls. Among these, there were 1,207 IBD cases and 6,992 controls of SAS ancestry, and 14,393 IBD cases and 15,456 controls of EAS ancestry (Fig. 1a, Extended Data Fig. 1). Using conditional analysis in our cohorts with access to in-sample linkage disequilibrium (LD) data (EUR and SAS datasets), we identified 619 independent genome-wide significant association signals for CD, UC or the combined IBD phenotype, 374 of which have not been reported previously (Methods, Fig. 1b,c, Extended Data Fig. 1, Supplementary Table 2). These signals included 128 associated with CD and 118 with UC, whereas the remainder were associated with both subtypes, either with consistent effect sizes (IBD unsaturated, N = 269) or significantly different effect sizes (IBD saturated, N = 104).

**Figure 1.**
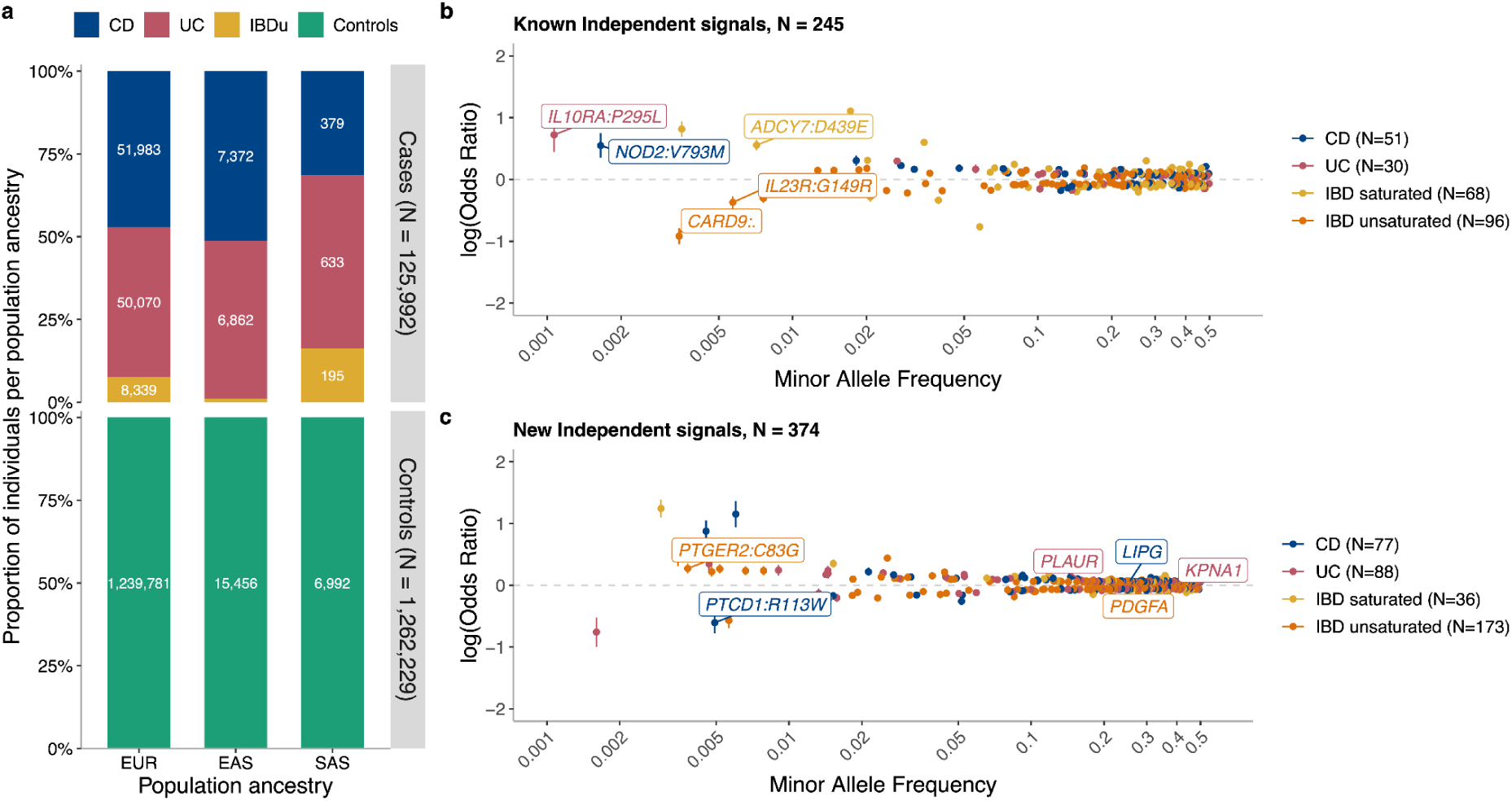
Multi-ancestry GWAS meta-analysis identifies 619 independent IBD association signals. a, Numbers of individuals included as inflammatory bowel disease (IBD) cases and controls, stratified by principal component (PC)-inferred genetic ancestry: European (EUR), East Asian (EAS) and South Asian (SAS). Case counts are partitioned into Crohn’s disease (CD), ulcerative colitis (UC) and IBD-unclassified (IBDu). b,c, Independent association signals plotted by effect size (log(odds ratio), with 95% confidence intervals) against minor allele frequency for the index variant at each signal. Colours indicate disease classification of the signal: CD-specific, UC-specific, IBD-unsaturated, or IBD-saturated. b, Previously reported independent signals validated in this study. c, Independent signals not reported previously.

All independent signals showed no sex-specific effects (Supplementary Table 3, Supplementary Note) and consistent effects in both EUR and multi-ancestry meta-analyses (Supplementary Fig. 2a). However, the lead variant for 20 signals showed significant heterogeneity in effect size attributed to ancestry (MR-MEGA ancestry-specific Het P < 8.08 x 10^-5^, Fig. 2b; Extended Data Fig. 2a, Supplementary Table 2). These results likely reflect a failure to capture the causal variant as the lead variant, and instead capturing tag variants with different LD patterns with the causal variant across ancestries (Supplementary Note). In line with previous studies focused on EAS populations ^8^ (which contributes to our total EAS cohort), we did not detect any SAS ancestry-specific significant associations.

**Figure 2.**
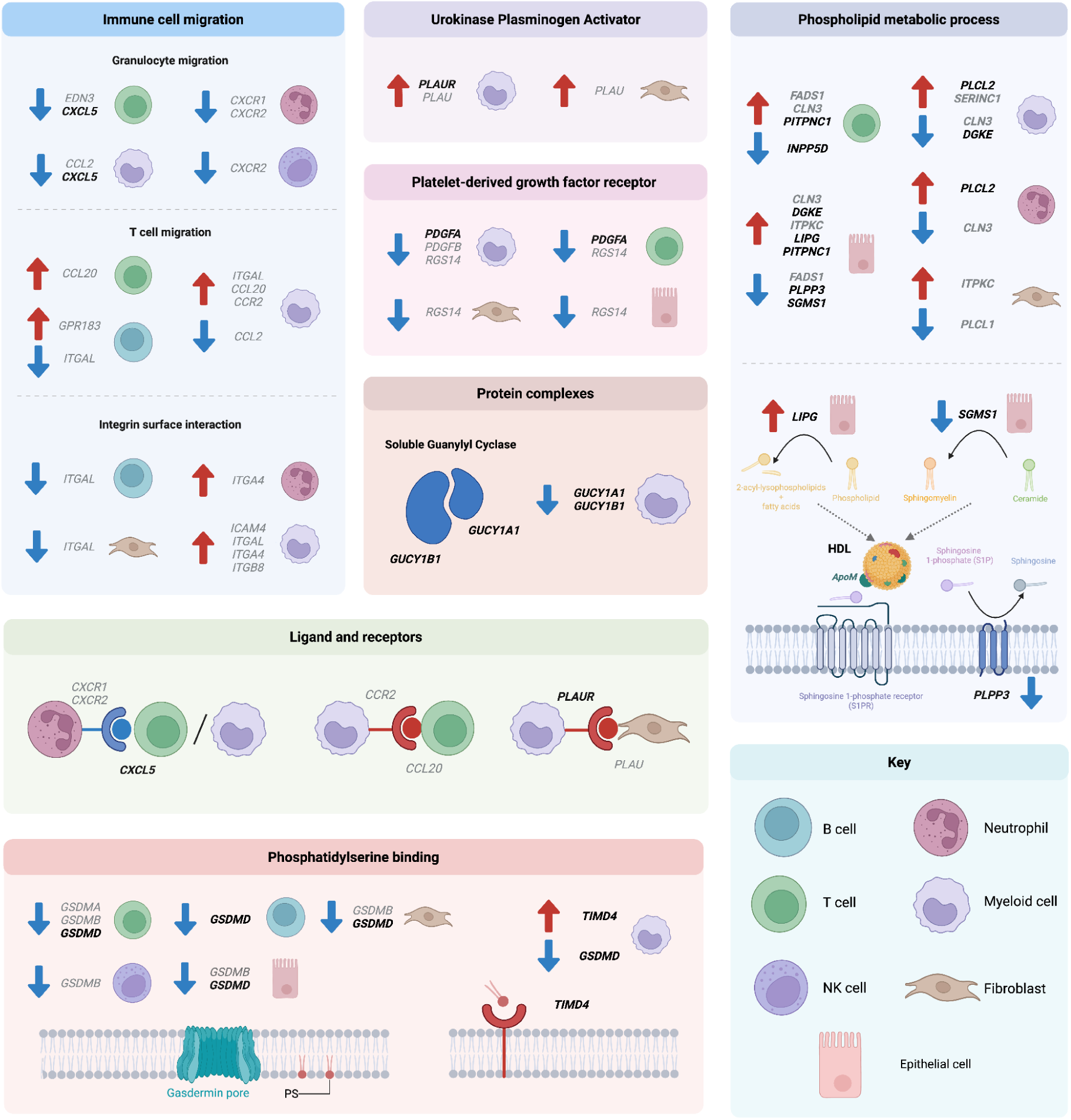
Nominated candidate effector genes converge on selected biological modules. Arrows indicate whether the IBD risk allele is associated with higher (red) or lower (blue) expression of the indicated gene in the specified cell type. Bold text denotes newly implicated genes, defined as genes without previous genetic association evidence for IBD, CD orUC in the Open Targets Platform (OTAR). Modules shown summarise selected pathways and ligand-receptor interactions supported by nominated candidate effector genes and direction-of-effect analyses. PS, phosphatidylserine.

## Identifying likely causal variants via fine-mapping

To identify the most likely causal variants underlying the independent association signals, we performed fine-mapping analyses using SuSiE^10^. We first collapsed the independent signals into genomic regions (N IBD regions = 420, Extended Data Fig. 1, Methods). Within each defined region, fine-mapping analyses were conducted in the subset of cohorts for which individual-level genotyping array data were available (EUR Tier 1, Methods, Supplementary Note), enabling accurate estimation of in-sample LD^11^. However, this also limited the number of IBD regions that could be fine-mapped (N = 170; Methods; Extended Data Fig. 3), as the lead variants did not reach genome-wide significance in this subset or had insufficient sample coverage.

Among these regions, we defined 268 credible sets (Supplementary Table 4), of which 81 were classified as high-confidence, defined by including variants with high posterior inclusion probability (PIP ≥ 0.5) and high sample coverage (≥ 95%). Among these 81 sets, 25 resolved to a single variant, including 7 located in coding regions and 18 in non-coding regions. Of the 81 lead variants from high-confidence credible sets, 24 had been reported previously^5,6^, whereas 41 were classified as novel after excluding variants in linkage disequilibrium with earlier signals (r² > 0.25) (Table 1; Methods; Supplementary Note).

**Table 1.** Novel IBD fine-mapped causal variants.

| MarkerName | Phe | Prob | BETA | SE | p_val | MAF | Annotation | Effector gene(s)* |
| --- | --- | --- | --- | --- | --- | --- | --- | --- |
| chr1:66934185:C:T | IBD | 0.8637 | -0.7425 | 0.1305 | 1.29E-08 | 0.002 | MIER1_intronic |  |
| chr1:146860876:G:T | CD | 0.5735 | -0.1316 | 0.0194 | 1.20E-11 | 0.4169 | HYDIN2;NBPF12_intergenic | *NBPF12* (T cells) |
| chr1:146878365:A:G | CD | 0.7936 | -0.1281 | 0.019 | 1.70E-11 | 0.4523 | HYDIN2;NBPF12_intergenic | *NBPF12* (T cells) |
| chr1:151816019:G:A | UC | 0.8439 | 0.1268 | 0.0188 | 1.69E-11 | 0.1527 | RORC_intronic |  |
| chr1:206935894:C:T | IBD | 0.8885 | -0.3276 | 0.0522 | 3.59E-10 | 0.0103 | PIGR_intronic |  |
| chr1:212839129:T:TTA | CD | 0.6978 | -0.0808 | 0.0135 | 2.30E-09 | 0.3543 | SPATA45_intronic |  |
| chr2:27508073:T:C | CD | 0.5852 | -0.1388 | 0.0128 | 1.36E-27 | 0.4382 | GCKR_exonic | *GCKR* (exonic)<br>*PPM1G* (Macrophages) |
| chr2:159820864:C:G | IBD | 0.6137 | -0.0646 | 0.0116 | 2.41E-08 | 0.3027 | LY75;LY75-CD302_intronic |  |
| chr2:162404181:T:C | UC | 0.6904 | 0.1165 | 0.0186 | 3.46E-10 | 0.1462 | KCNH7_intronic |  |
| chr2:217051329:T:G | CD | 0.5224 | -0.0765 | 0.0127 | 1.84E-09 | 0.4442 | LINC01921;DIRC3-AS1_intergenic |  |
| chr2:240630275:C:T | IBD | 0.6883 | 0.1174 | 0.0137 | 1.08E-17 | 0.1815 | GPR35:T108M | *GPR35* (exonic) |
| chr2:241816853:C:T | IBD | 0.6048 | 0.1162 | 0.0171 | 1.03E-11 | 0.1353 | NEU4_exonic | *NEU4* (exonic)<br>*PDCD1* (Macrophages) |
| chr4:3445027:G:A | CD | 0.9053 | 0.1587 | 0.0216 | 2.28E-13 | 0.1033 | HGFAC_intronic |  |
| chr4:3447925:G:A | IBD | 0.5684 | 0.1488 | 0.0205 | 3.81E-13 | 0.0753 | HGFAC:R509H |  |
| chr5:38867630:C:T | CD | 0.6417 | 0.0957 | 0.0126 | 2.98E-14 | 0.4926 | OSMR_intronic |  |
| chr5:130724057:G:A | CD | 0.505 | 0.1316 | 0.0139 | 2.38E-21 | 0.3476 | CHSY3;HINT1_intergenic | *P4HA2* (B-cells, Macrophages, Monocytes, Neutrophils, Secretory cells, Colonocytes) |
| chr5:159498514:GA:G | UC | 0.5936 | -0.2553 | 0.0262 | 1.92E-22 | 0.0776 | LINC01845;LINC01847_intergenic |  |
| chr6:44264337:A:G | IBD | 0.6725 | 0.0631 | 0.0111 | 1.33E-08 | 0.4305 | NFKBIE_intronic | *NFKBIE* (Macrophages) |
| chr7:27192143:C:T | UC | 0.5332 | -0.0963 | 0.0169 | 1.11E-08 | 0.1969 | HOXA11-AS;HOXA13_intergenic | *ENSG00000253508 HOXA13* (Colonocytes) |
| chr8:144103704:G:A | CD | 0.9997 | 0.2225 | 0.0253 | 1.61E-18 | 0.0919 | SHARPIN:S17F | *SHARPIN* (exonic) |
| chr9:274641:T:C | IBD | 0.8851 | -0.0732 | 0.011 | 2.69E-11 | 0.3845 | DOCK8_intronic |  |
| chr9:34710087:A:C | CD | 0.5793 | -0.1131 | 0.0195 | 6.97E-09 | 0.1242 | CCL21_UTR5 | *CCL21* (blood plasma)<br>*SPATA31F1* (exonic) |
| chr9:114881082:C:T | IBD | 0.7126 | 0.0934 | 0.0122 | 2.27E-14 | 0.2525 | TNFSF15;TNFSF8_intergenic | *TNFSF15* (Macrophages, Monocytes, Dendritic Cells 2; Enterocytes) |
| chr10:62710915:C:T | IBD | 0.9933 | -0.1407 | 0.0146 | 4.42E-22 | 0.1509 | ZNF365;ADO_intergenic |  |
| chr10:96798744:T:A | UC | 0.9503 | 0.1337 | 0.0215 | 5.15E-10 | 0.1138 | RPL13AP5;MIR607_intergenic | *LCOR* (Colonocytes) |
| chr10:110426390:C:T | CD | 0.7706 | 0.1157 | 0.0134 | 6.20E-18 | 0.3409 | SMNDC1;DUSP5_intergenic |  |
| chr11:2299865:C:A | IBD | 0.9888 | -0.0764 | 0.011 | 3.79E-12 | 0.4999 | C11orf21_intronic | *ASCL2* (B cells, NK Cells, T cells) |
| chr12:6395607:T:G | CD | 0.9693 | 0.1018 | 0.0138 | 1.50E-13 | 0.4456 | LTBR;CD27-AS1_intergenic | *ENSG00000256433 LBTR1* (Macrophages, Secretory cells, Colonocytes, Enterocytes) |
| chr13:40222015:G:A | IBD | 0.6573 | -0.1506 | 0.026 | 6.79E-09 | 0.0418 | LINC00548;LINC00598_intergenic |  |
| chr16:50712018:C:T | CD | 1 | 0.5762 | 0.0798 | 5.15E-13 | 0.0072 | NOD2:R676C | *NOD2* (exonic) |
| chr16:85980635:T:C | UC | 0.5278 | 0.1452 | 0.0171 | 1.72E-17 | 0.1855 | IRF8;LINC01082_intergenic | *IRF8* (Colonocytes, Enterocytes, Intestinal Epithelial Stem cells) |
| chr17:40713238:A:T | CD | 0.6575 | 0.3519 | 0.0451 | 6.30E-15 | 0.0263 | KRT24;KRT25_intergenic |  |
| chr17:42365698:C:T | IBD | 0.7545 | -0.372 | 0.0526 | 1.53E-12 | 0.0102 | STAT3_intronic |  |
| chr19:10354167:G:A | CD | 0.5318 | -0.5817 | 0.0814 | 8.98E-13 | 0.0065 | TYK2:A928V | *TYK2* (exonic) |
| chr19:33264218:T:C | UC | 0.7216 | 0.1621 | 0.0155 | 1.56E-25 | 0.2726 | SLC7A10;CEBPA_intergenic | *CEBPA* (Macrophages) |
| chr19:33268130:T:G | CD | 0.6201 | 0.1199 | 0.0141 | 2.17E-17 | 0.3285 | SLC7A10;CEBPA_intergenic | *CEBPA* (Macrophages) |
| chr19:40712706:G:A | CD | 0.7798 | -0.0735 | 0.0128 | 1.01E-08 | 0.4989 | COQ8B_intronic | *ITPKC* (Colonocytes, Enterocytes, Fibroblasts) |
| chr20:6084192:T:C | IBD | 0.9564 | 0.0675 | 0.0114 | 3.48E-09 | 0.3223 | FERMT1_intronic |  |
| chr20:64080106:C:T | UC | 0.5391 | 0.076 | 0.0138 | 3.72E-08 | 0.4805 | OPRL1_UTR5 |  |
| chr21:33397315:TAA:T | CD | 0.5802 | 0.1043 | 0.013 | 1.05E-15 | 0.4346 | IFNAR1;IFNGR2_intergenic | *IFNGR2* (Colonocytes, Enterocytes, Secretory cells, Intestinal Epithelial Stem cells) |
| chr21:44095466:T:G | CD | 0.9911 | -0.0831 | 0.0149 | 2.51E-08 | 0.352 | TRAPPC10_intronic | *ENSG00000260256* (Macrophages) |

## IBD-associated regions capture most SNP-based heritability

To quantify how much heritability is now captured by the expanded maps of disease-associated regions, we estimated SNP-based heritability using LD score regression^12^ and EUR summary statistics (Methods). On the liability scale, SNP-based heritability was 0.24 for IBD, 0.31 for CD, and 0.25 for UC (Extended Data Fig. 4a,b). Using stratified LD score regression^13^, variants within the IBD-associated regions (N = 420) captured 0.77–0.80 of SNP-based heritability (0.77, 0.80 and 0.77 for CD, IBD and UC, respectively), a 20% increase compared with previous reports (Extended Data Fig. 4c; Methods). Thus, most SNP-based heritability for IBD is now captured within IBD-associated regions. Further variant discovery in European populations will require substantially larger sample sizes, because additional loci are likely to explain progressively smaller fractions of heritability (Supplementary Note).

## Nominating candidate effector genes

To nominate candidate effector genes mediating disease risk, we integrated EUR GWAS summary statistics with bulk and single-cell expression QTL datasets (N = 492), protein QTL datasets (N = 3) and results from the IIBDGC exome sequencing study^14^, applying complementary approaches including colocalisation analysis and Mendelian randomization (MR).

To maximize IBD relevance, peripheral blood pQTL were also calculated using data from people with IBD. The SPARC IBD cohort includes 1,021 IBD cases (643 CD and 378 UC) with both plasma protein levels and genotype data (Methods)^15^. In total, 43,387 pQTL with P < 1.7 x 10^-11^ (5 x 10^−8^ adjusted for the 2,923 unique proteins in the Olink panel) were identified, of which 4,531 overlap significant (P < 5 x 10^-7^) GWAS results. The pQTL are also highly consistent with those identified in UK Biobank, with 406 out of 407 exact SNP-protein matches showing the same direction in both datasets when comparing beta coefficients and allele order^16^. This indicates that inflammation does not alter the directionality of pQTLs, underscoring their potential utility as biomarkers. Reflecting the small IBD cohort size, we did not observe any trans-pQTLs.

Colocalisation analyses prioritised 625 candidate effector genes across 305 independent association signals. A ranking framework informed by cell-type–specific QTL enrichment of IBD variants designated 322 genes as highest priority (defined as ‘rank 1’; Methods; Supplementary Note). MR prioritised 117 genes across 89 IBD regions whose encoded proteins are implicated in susceptibility to IBD, CD or UC; 18 genes harboured cis-pQTL variants in strong LD with the corresponding GWAS index variant (r² ≥ 0.6) at 12 signals, enabling direct signal–gene assignment. Integration with curated exonic and splice-altering variants from the IIBDGC exome sequencing study^14^ revealed that 8% of independent signals (51 variants across 37 genes) were likely driven by coding variation (Supplementary Table 5). Increased GWAS power and improved imputation enabled detection, for the first time through array-based association analysis, of signals driven by three well-established low-frequency coding variants previously identified only through sequencing (*CCR7*:p.M7V, *DOK2*:p.P274L, *IL10RA*:p.P295L), as well as 14 novel coding-variant signals.

Across all approaches, we nominated 664 candidate effector genes across 341 independent association signals (Supplementary Table 6). Using coding-burden heritability as an orthogonal benchmark, our combined nomination strategy outperformed a nearest-gene approach (Extended Data Fig. 5). Of the 341 signals, 172 were assigned a single gene, whereas 169 were assigned to multiple candidate effector genes, most frequently through colocalisation. Signals without nominated genes had significantly lower minor allele frequencies than those with nominated genes (P = 3.7 × 10⁻⁴), particularly for colocalisation-based assignments (P = 8.9 × 10⁻^7^) (Supplementary Note).

The three approaches captured complementary mechanisms linking genetic variation to disease, including regulatory effects identified through QTL colocalisation and MR, and protein-altering variation identified through ExWAS. Accordingly, for 314 of 341 signals (92%), candidate effector genes were supported by only a single approach, most frequently colocalisation (N = 279), highlighting the complementary value of regulatory and coding-based strategies. At the gene level, 39 of 664 candidate effector genes were nominated by more than one independent association signal, providing independent support for prioritization, including newly implicated genes such as *TIMD4* and *PLAAT3*.

To assess prior support for the nominated candidate effector genes in IBD, we queried the Open Targets Platform (OTAR)^17^. Of the 664 candidate effector genes, 274 had previous genetic association evidence for IBD. Among the remaining 390 genes, 203 were designated high-priority candidates based on rank 1 colocalisation, MR or exonic evidence, whereas 187 were supported by lower-priority colocalisation. High-priority genes were enriched for additional non-genetic evidence in OTAR compared with lower-priority colocalisation genes (odds ratio = 2.25, χ² test P = 3.88 × 10⁻^4^). Together, these analyses provide further support to the higher-confidence subset of newly nominated candidate effector genes for downstream functional validation.

Finally, we assessed the direction of effect of associated variants on gene expression and plasma protein levels. Of the 625 genes prioritised through colocalisation, 604 were supported by more than one QTL dataset, with 526 of them showing consistent directions of effect across tissues, cell types or datasets (Supplementary Table 5; Methods).

## Biological context of nominated candidate effector genes

Candidate disease effector genes were significantly enriched in pathways related to immune-cell activation and trafficking, cytokine and integrin signalling, and downstream kinase cascades (Supplementary Table 7). These genes were also enriched among curated ligand–receptor pairs from CellTalkDB (OR = 5.9; P = 2.2 × 10⁻^6^), genes implicated in monogenic forms of IBD^4^ (OR = 3.8; P = 2.4 × 10^⁻3^; N = 7 out of 64) and genes underpinning primary immunodeficiencies (OR = 2.3; P < 2 × 10⁻⁶; N = 42 out of 999; Genomics England Panel 398), supporting the biological coherence of the nominated gene set.

Genes involved in granulocyte migration (Fig. 2) showed reduced expression associated with IBD risk alleles, including the neutrophil chemokine receptors *CXCR1* and *CXCR2* and their ligand *CXCL5*, a gene not previously associated with IBD. In contrast, IBD risk alleles were associated with increased expression of genes involved in T cell and monocyte trafficking and integrin-mediated cell-surface interactions, including ligand-receptor pairs such as *CCR2* and *CCL20*. Together, these observations further support immune-cell trafficking as a central mechanism through which genetic risk contributes to IBD.

Several additional biological modules were supported by coherent direction-of-effect patterns across nominated candidate effector genes (Fig. 2). These included the urokinase plasminogen activator pathway, soluble guanylyl cyclase signalling, phosphatidylserine binding, phospholipid metabolic processes and interferon signalling. Among these, two newly implicated genes, *GUCY1A1* and *GUCY1B1*, encode the two subunits of soluble guanylyl cyclase, and IBD risk alleles were associated with decreased expression of both genes in macrophages and monocytes, driven by the same association signal. In addition, three independent risk alleles affected expression of *LIPG, PLPP3* (LPP3) and *SGMS1*, newly implicated genes linked to phospholipid metabolism and HDL-related biology^18,19^. Together, these analyses nominate a set of genetically supported biological modules through which risk alleles may contribute to IBD pathogenesis.

## Shared genetic architecture highlights candidate biomarkers and causal mediators

To place IBD genetics in a broader biological and clinical context, we asked which complex traits share genetic architecture with IBD, and whether any of these relationships are consistent with causal mediation rather than correlation alone.

We used cross-trait LD score regression and EUR summary statistics to estimate genetic correlations between IBD, CD and UC and 347 complex traits spanning immune-mediated diseases, circulating biomarkers and blood cell traits (Methods). In total, 87 traits showed significant genetic correlation with IBD, CD or UC (Extended Data Fig. 6). As expected, the strongest correlations were observed with immune-mediated diseases, including ankylosing spondylitis, psoriasis and primary sclerosing cholangitis. Among circulating traits, HDL- and Apolipoprotein A-I (ApoA-I)-related measures showed significant negative genetic correlations with IBD, whereas systemic inflammatory biomarkers, including C-reactive protein, phenylalanine and glycoprotein acetylation, showed positive and predominantly stronger genetic correlations with CD than with UC.

To assess whether any of these correlations were consistent with causal mediation, we applied a latent causal variable (LCV) model to trait pairs showing significant genetic correlation (Methods). HDL-related traits and ApoA-I showed evidence consistent with near-complete genetic causality (as estimated by the LCV model), supporting a protective effect on IBD and CD risk, respectively (|GCP| ≥ 0.6; q < 0.05) (Fig. 3). The LCV model also supported a causal effect of CD liability on increased circulating phenylalanine (Fig. 3), consistent with prior work suggesting phenylalanine as a potential biomarker for CD^20^. Together, these analyses highlight circulating biomarkers that either reflect shared inflammatory biology with IBD or may lie on causal pathways influencing disease susceptibility.

**Figure 3.**
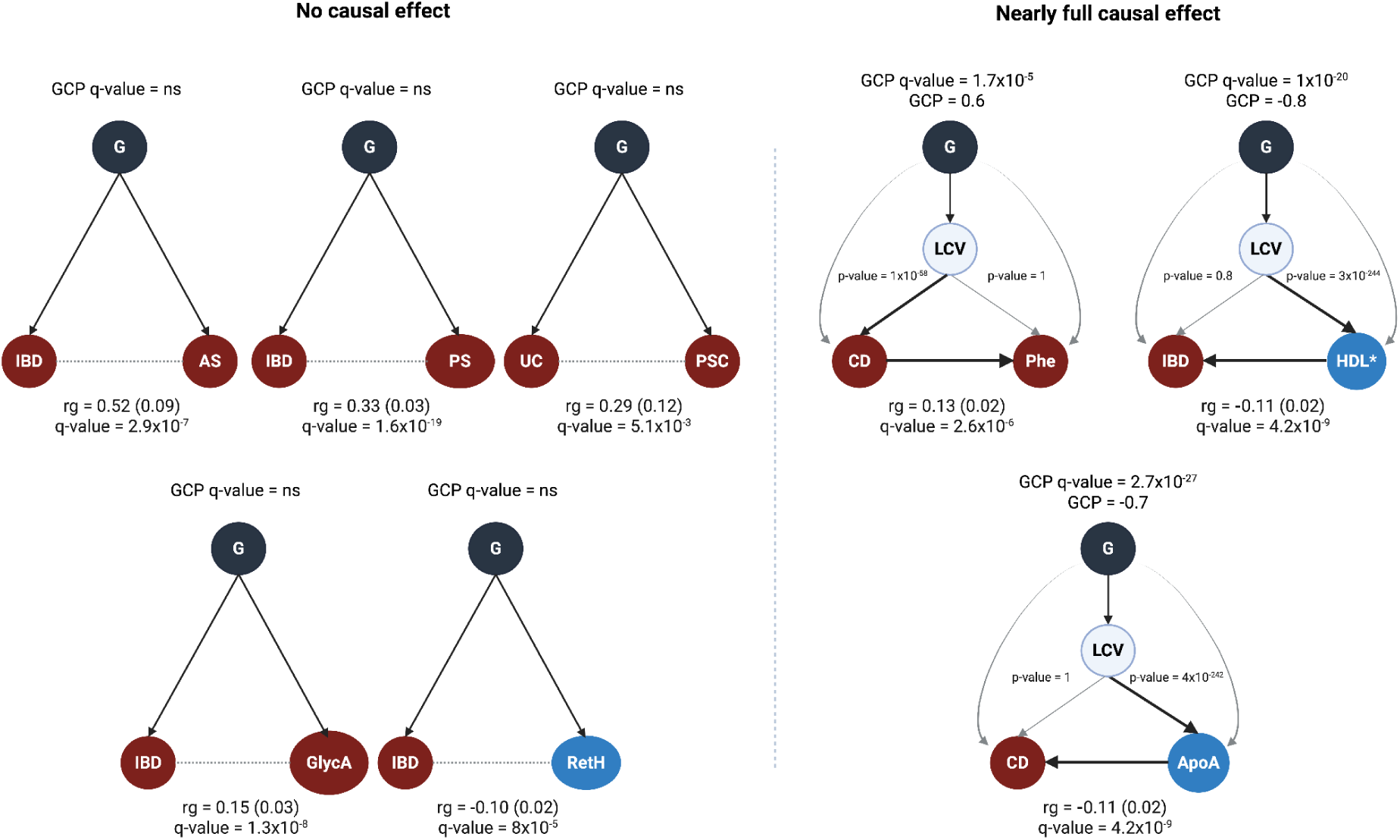
Shared genetic architecture and latent causal variable analyses highlight candidate biomarkers and mediators. Genetic correlation estimates (rg) are shown for selected traits remaining significant after multiple-testing correction (q < 0.05), with standard errors in parentheses. Directed acyclic graphs illustrate LCV results for trait pairs with either no evidence of genetic causality (GCP q-value not significant) or evidence consistent with near-complete genetic causality (|GCP| ≥ 0.6). Black arrows indicate the inferred direction of effect between traits. P values shown next to the arrows correspond to tests of the inferred causal direction under the LCV model. G, genetic component; AS, ankylosing spondylitis; PS, Psoriasis; PSC, primary sclerosing cholangitis; GlycA, glycoprotein acetylation; RetH, reticulocyte haemoglobin equivalent; Phe, phenylalanine; HDL, total cholesterol in high-density lipoprotein; ApoA, apolipoprotein A-I. * Total cholesterol levels in HDL

## Cell-type context of IBD-associated genetic variation

We applied stratified LD score regression (S-LDSC) to compare enrichment of CD- and UC-associated genetic risk across annotations derived from different cell types and states. Across all significant results, the strongest enrichments were observed for immune-derived annotations, including those from B cells, T cells and myeloid cells. In datasets profiling immune cells under both resting and stimulated conditions, stimulated states consistently showed higher enrichment estimates than resting states (Supplementary Note).

Immune cell subsets, including T cells and macrophages, as well as fibroblasts, showed significant and broadly comparable enrichment for both CD and UC (Fig. 4a). In contrast, S-LDSC coefficient z-scores were, on average, 1.5-fold higher for UC than for CD in epithelial cell types, including enterocytes, goblet cells and tuft cells, consistent with a stronger contribution of intestinal epithelial annotations to UC than to CD susceptibility (Fig. 4b).

**Figure 4.**
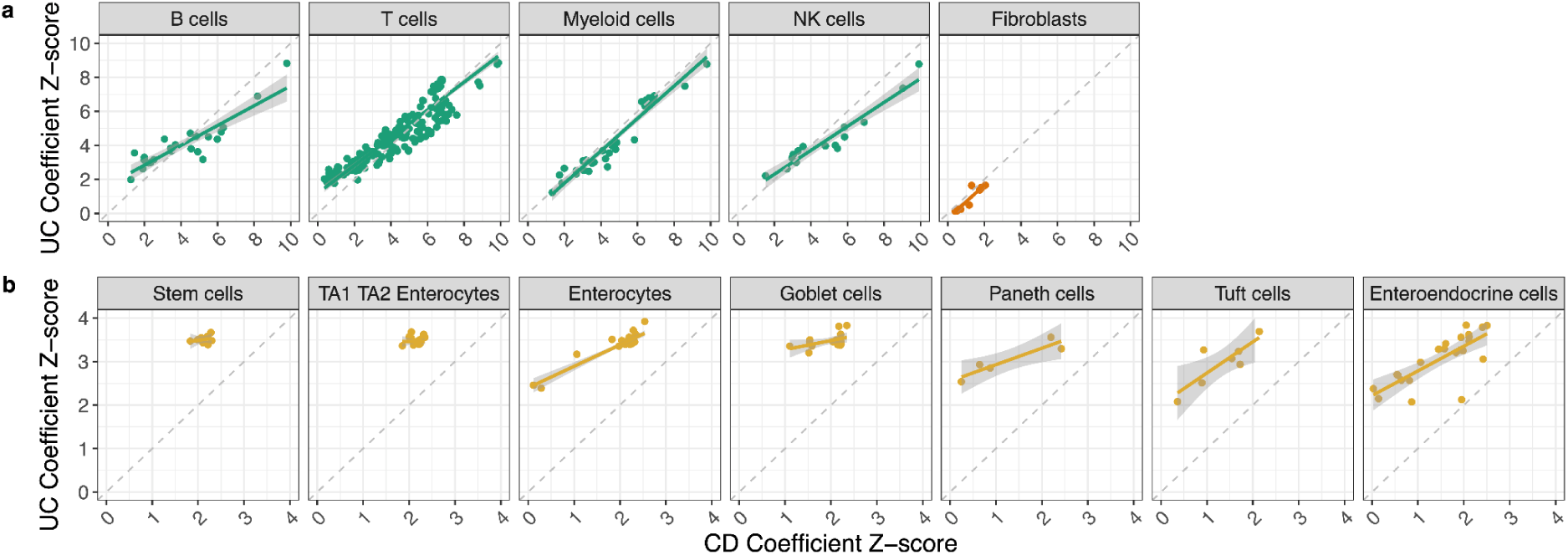
Comparative enrichment of CD and UC genetic risk across cell-type-derived annotations. Pairwise comparison of stratified LD score regression (S-LDSC) coefficient z-scores across annotations derived from (a) immune cells and fibroblasts and (b) intestinal epithelial cell types, aggregated across studies. Each point represents an independent estimate for a major cell type with more than four observations. The x axis shows the S-LDSC coefficient z-score for Crohn’s disease (CD); the y axis shows the corresponding z-score for ulcerative colitis (UC). Axis limits differ between panels. Immune cell annotations include both peripheral blood-derived and tissue-resident populations. TA1 and TA2 denote transit-amplifying enterocyte states positioned between stem cells and mature enterocytes, as defined in^21^

## Therapeutic implications of nominated candidate effector genes

To assess the therapeutic relevance of the nominated candidate effector genes, we asked whether they overlapped known drug targets, supported drugs in clinical development for IBD, and highlighted opportunities for repurposing or anticipating adverse effects. We therefore mapped prioritised candidate effector genes to therapeutic targets and associated drugs using the OTAR platform^17^.

Overall, we identified 283 known drugs investigated in clinical trials for IBD, CD or UC, involving 365 drug–target pairs (Supplementary Table 8). Forty-four pairs were supported by our study, 39 with high confidence, and these were 1.9-fold higher odds to reach phase ≥ 3 than unsupported pairs (Fisher’s exact test P = 0.03). This included support for the inferred direction of effect of several established IBD treatments, including IL-23 inhibitors (*IL23R*), IL-12/23 inhibitors (*IL12B*), the anti-integrin vedolizumab (*ITGA4*–*ITGB7*), and JAK inhibitors (*JAK2*) (Fig. 5). We also identified additional genetic support for additional agents currently in phase 3 IBD trials, including inhibitors of *TNFSF15* (TL1A) and targeted agents against its receptor *TNFRSF25* (DR3).

**Figure 5.**
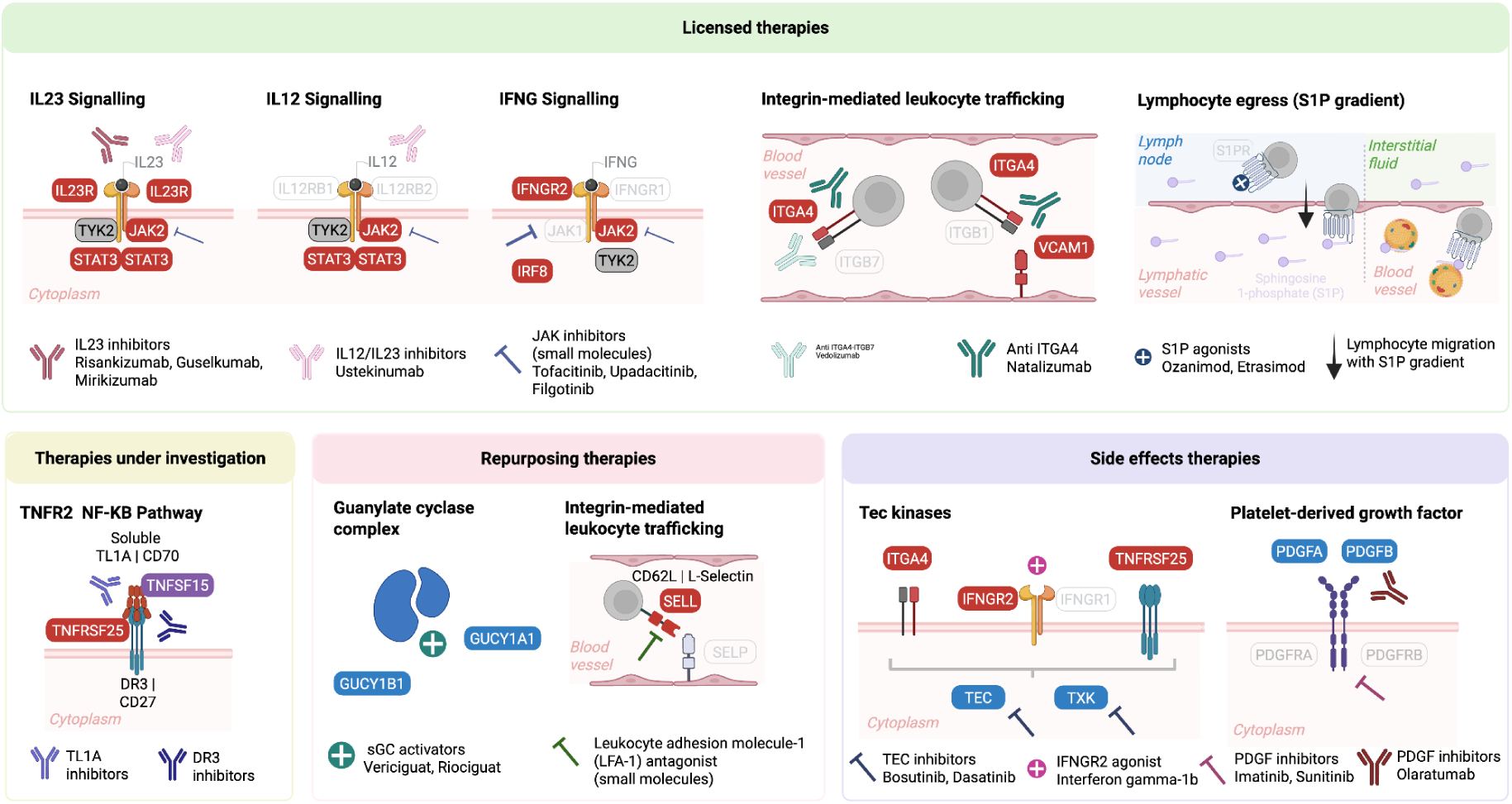
Genetic support for therapeutic targets, repurposing opportunities and adverse effects in IBD. Modules are grouped into licensed therapies, therapies under investigation, repurposing opportunities, and pathways in which target inhibition or activation is associated with adverse effects. Background colour indicates the inferred direction of genetic support for the highlighted gene or target: red, IBD risk allele associated with higher expression; blue, IBD risk allele associated with lower expression; purple, risk allele associated with both higher and lower expression in different cell types; grey, coding-variant support; white, no IBD-associated gene. Drug classes or representative compounds are shown below each module. Although not listed among the effector genes, protein-truncating variants in *IL12RB1* (IL12 signalling) show significant protective effect on IBD^14^

We next asked whether the same framework could highlight drug repositioning opportunities. In total, 44 IBD-prioritised genes with high confidence are targets for 242 known drugs evaluated for other diseases (269 drug-target pairs; Supplementary Table 9). Assessing concordance between the inferred direction of pharmacological modulation and the direction suggested by our genetic analyses identified 102 compounds linked to 19 high confidence IBD genes with concordant effects. This set includes soluble guanylate cyclase activators targeting *GUCY1A1* and *GUCY1B1*, approved for pulmonary hypertension and coronary artery disease. In our data, the IBD risk allele is associated with reduced expression of both genes in myeloid cells, suggesting that agonists of this pathway may also be beneficial in IBD. Among the concordant compounds were 16 agents that have progressed beyond phase II, including a leukocyte adhesion molecule-1 antagonist targeting L-Selectine (*SELL*), evaluated in sickle cell disease.

Genetic data can also anticipate adverse effects. Modern oncology agents frequently induce immune-mediated toxicities, including colitis. Multiple anticancer agents target PDGF signalling, either by inhibiting PDGF receptors directly or by disrupting PDGFA/PDGFB-driven signalling through their tyrosine kinase receptors, with varying specificity and differing reported risks of colitis^22,23^. In our data, IBD risk alleles were associated with lower expression of *PDGFA* and *PDGFB*, providing genetic support for clinical observations that inhibition of this pathway may promote colitis.

Similarly, members of the Tec kinase family (*TEC* and *TXK*) were associated with IBD. Tec-deficient models develop T-cell-mediated intestinal inflammation^24^, and inhibitors that non-specifically target Tec-family kinases, including bosutinib and dasatinib, are associated with gastrointestinal complications in oncology patients^25,26^. In our data, IBD risk alleles were linked to decreased expression of both genes in macrophages, consistent with these observations. Together, these analyses show that human genetics can help prioritise therapeutic targets, identify repurposing opportunities, and anticipate mechanism-based adverse effects in IBD.

## Discussion

In this study, we assembled the largest multi-ancestry GWAS meta-analysis of inflammatory bowel disease to date and identified 374 previously unreported association signals. Together with previously reported loci validated here, these explain approximately 77% of the heritability attributable to the IBD regions captured in this study (MAF ≥ 0.001). The expanded map also identified 51 association signals likely to be driven by coding variation, supported by orthogonal evidence from exome sequencing, including 11 not previously reported. Although individuals of non-European ancestry comprised up to 13% of IBD cases, power remained insufficient to detect ancestry-specific signals, emphasising the need for substantially larger and more diverse studies.

By integrating plasma proteomic and transcriptomic QTL data with our GWAS results, we nominated 664 candidate effector genes across 343 association signals. These analyses improve resolution of the genes likely to mediate genetic risk, but also show that an important remaining challenge is to define the genes, cell types and molecular effects through which associated variants act. Most signals were assigned genes through QTL integration, whereas signals without nominated genes had significantly lower minor allele frequencies, suggesting that regulatory effects driven by lower-frequency variants remain underpowered in current QTL datasets^27^. Additional signals are also likely to act through cell types, states or disease contexts that are not yet adequately represented in available molecular atlases^28,29^.

The fact that we nominated substantially more candidate effector genes than association signals should not be interpreted solely as over-assignment. CRISPR-based perturbation studies have shown that cis-regulatory elements can affect the expression of multiple nearby genes, indicating that a single regulatory association signal may legitimately map to more than one effector gene^30–32^. At the same time, the variable strength of evidence across genes at the same locus emphasises the need for prioritisation and experimental validation.

Combined monogenic and GWAS-implicated genes increase the relative success of gene-target mappings in clinical trials^9^. Our results show that continued GWAS discovery remains biologically and therapeutically informative, with enhanced sample sizes improving both locus discovery and fine-mapping resolution. Newly implicated candidate effector genes include targets of licensed and emerging IBD therapies, indicating that therapeutic relevance is only weakly related, if at all, to common-variant effect size^9^. These observations argue against a strong interpretation of the omnigenic model, where later-discovered common-variant associations implicate increasingly peripheral biology^33–35^. Instead, they suggest that increasingly well-powered GWAS can identify biologically central genes despite modest individual effect sizes.

CD and UC showed broadly similar genetic architectures, reflected in the large proportion of shared signals and their strong genetic correlation. Despite this shared background, we detected subtype-specific enrichment in distinct cellular programmes. Our findings refine previous work^36,37^ by showing stronger enrichment in UC across several epithelial cell types, including enteroendocrine cells and secretory lineages such as Paneth, goblet and tuft cells, whereas immune-cell and fibroblast enrichments were broadly comparable between CD and UC. Rather than indicating wholly distinct disease architectures, these results suggest differences in the relative contribution of shared programmes, with epithelial biology contributing more strongly to UC than to CD.

Our cross-trait analyses demonstrate that human genetics can help distinguish circulating biomarkers that primarily reflect downstream inflammatory burden from traits that may lie closer to pathways influencing disease susceptibility. We identified shared genetic architecture between IBD and several inflammatory and metabolic traits, including C-reactive protein (CRP), phenylalanine and glycoprotein acetylation (GlycA). The stronger genetic overlap between CRP and CD mirrors clinical observations that CRP is less reliable for monitoring disease activity in UC^38^, suggesting a genetic basis for this difference. Our results also extended the set of circulating traits genetically linked to CD to include phenylalanine and GlycA, supporting their relevance as biomarkers of disease-associated inflammatory burden^20,39,40^. We further identified a shared genetic component between IBD and circulating HDL levels and ApoA-I, with causal modelling suggesting that genetically determined lower HDL levels increase IBD susceptibility. Our findings support the view that some circulating traits identified through cross-trait analyses may index pathways closer to disease causation rather than simply downstream inflammation. Consistent with this, we identify multiple IBD-associated genes involved in phospholipid metabolism, including three genes expressed in intestinal epithelial cells: *LIPG*, *PLPP3* and *SGMS1*. These genes implicate HDL-related lipid metabolism in the disease process^18,19^, suggesting a mechanistic link to lipid signalling pathways converging around sphingosine-1-phosphate receptors (S1PR)^41^, a family of receptors targeted by therapies approved for UC. This link to S1P signalling arises from the genetic evidence itself rather than from drug target overlap, providing orthogonal support for a therapeutically validated pathway. HDL has an established role as the main plasma chaperone for S1P. However, a mechanistic link for this association, potentially mediated through the effects on gut immune cell trafficking, will need to be functionally validated.

Our drug-target analyses show that this increasingly resolved genetic map has direct translational relevance. Genetics supported the inferred direction of effect for several established or late-stage IBD therapies, highlighted additional repurposing opportunities, and in some cases anticipated mechanism-based adverse effects. Taken together, enrichment of the nominated candidate effector genes for established drug targets and monogenic IBD genes, together with the ability of the framework to recover expected therapeutic directions and anticipate adverse effects, increases confidence that our nominated genes capture biologically important effectors of disease. This supports their use in conceptualising and prioritising novel therapeutic targets for IBD.

Several limitations remain. Despite the multi-ancestry design of the discovery meta-analysis, downstream fine-mapping and many mechanistic analyses were still driven primarily by European-ancestry data. Many candidate effector genes were supported by only a single line of evidence, and some signal-to-gene assignments will therefore remain provisional until tested experimentally. Similarly, causal inference for circulating traits is model-based and should not be interpreted as equivalent to evidence from intervention. Taken together, our results suggest that the next phase of IBD genetics will be defined less by discovering additional loci and more by resolving how associated variants act through genes, cell states and biomarkers to identify therapeutic targets.

## Supporting information

Named_Authors

Banner_Authors

Cohorts_Descriptions

## Methods

### GWAS summary statistics quality control and meta-analysis

We used a combination of in-house scripts to perform quality control centrally for the GWAS summary statistics in each cohort (Supplementary Note). In summary, we excluded (1) variants with low call rate (<0.95 for variants with MAF > 0.01; <0.98 for variants with MAF ≤ 0.01); (2) variants with a significant difference in genotype call rate missingness (P <1×10^-4^) between cases and controls; (3) variants with allele frequency differences versus those from the frequency reported in gnomAD (release v3.0)^1^ non-Finish Europeans, or TOPMed^2^ global MAF (using the criterion ((p1 − p0)^2/((p1+p0)*(2-p1-p0)) > 0.025 and > 0.125, respectively), where p0 is the MAF in the reference panel and p1 the observed MAF in the study); (4) variants with a Hardy–Weinberg Equilibrium (HWE) P < 10-5 among controls and 10-12 among cases, estimated in EUR ancestry samples; and (5) monomorphic variants.

Imputation of non-directly genotyped data was carried out per cohort, using the multi-ancestry TOPMed as reference panel (r2@1.0.0), through the TOPMed imputation server (imputationserver@1.5.7)^3^. Post-imputation, we excluded variants with strong deviation from HWE (P HWE ≤ 10-5 in controls; or alternatively ≤ 10-12 in cohorts that only included cases).

The analyses were carried out using Regenie^4^ (v3.2.5), applying Firth approximation correction to variants with P ≤ 0.1, and including as covariates per-continental population ancestry PCs and sex.

We used METAL (version released 2011-03-25)^5^ to perform per-PC inferred ancestry fixed-effects (standard error weighted) meta-analysis (EUR, SAS). We combined the summary statistics from the SAS and EUR cohorts together with the EAS IBD summary stats released in Liu et al.^6^. We additionally ran a random-effects meta-analysis as implemented in MR-MEGA (v0.1.5)^7^. These analyses included 125,992 IBD cases and 1,262,229 population controls (Extended Data Table 1), from three different population ancestries, EUR, EAS and SAS.

### Independent signals

To define the newly associated regions we followed two different approaches: informed by previously defined signals and agnostic to these (Extended Data Fig. 1). We started by selecting all known index variants from published fine-mapped independent signals (extracted from refs^8,9^, defined as the variant with the largest posterior probability), or the GWAS index variant if the known region had not been previously fine-mapped^9^. Thus, we included 370 known index variants present in our EUR cohorts (out of 378, Supplementary Table 10). These analyses were run separately for CD, UC and IBD, and per continental ancestry (EUR and SAS). As the LD reference, we used in-sample genotyping array data for the European (EUR tier 1) and South Asian datasets to which we had access (see Supplementary Note). We first used COJO^10^ to define variants showing conditional genome-wide significance (P < 5×10-8) either in EUR tier 2 or in SAS, after accounting for the effect of any known signal. In addition, we ran the EUR and SAS conditional analyses via COJO, without conditioning on any previously reported signal. Variants reported in less than 50% of the total Neff were excluded, as well as variants showing significant heterogeneity in effect size (Het P value < 1×10-15). The total Neff was estimated as follows:

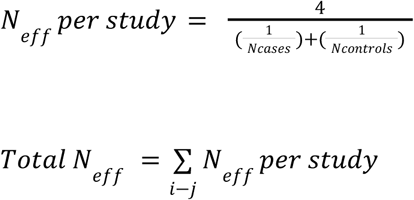

This retained 13,656,989 variants for IBD analyses, 13,909,224 for CD and 13,694,540 for UC.

We outlined the regions in the genome that included the genome-wide significant variants prioritised above via bedtools (v2.29.2)^11^, aggregating those into a single region when they were located <1Mb from each other, and expanding the intervals by +/- 500 kb from the most upstream and downstream variant per new region. Regions were also merged if they overlapped. The HLA region (as defined in https://www.ncbi.nlm.nih.gov/grc/human/regions/MHC ± 5 Mb) was excluded from this definition.

We finally used COJO^10^ to estimate the joint effects of all the selected variants, retaining only those showing genome-wide significance (P < 5×10^-8^) in the joint model (Extended Data Fig. 1). We further investigated those previously reported variants that could not be replicated in this study (Supplementary Note).

### Disease likelihood modeling

Each association signal was reclassified according to its relative association strength with each disease subtype (CD and UC) using a multinomial regression–based likelihood modelling approach, as previously described by^12^, applied to the two larger datasets comprising the EUR tier 1 cohort. For this analysis, we used the lead variant for each signal as defined above.

For each variant, we fitted four logistic regression models parameterized by two effect sizes (β_CD_; β_UC_) (R package *multinomRob*). Three constrained models were specified: a CD-specific model (β_UC_ = 0; 1 d.f.), a UC-specific model (β_CD_ = 0; 1 d.f.), and an IBD-unsaturated model (β_CD_ = β_UC_; 1 d.f.). We additionally fitted an unconstrained IBD-saturated model allowing both parameters to vary independently (2 d.f.). Likelihood-ratio tests were performed to compare each constrained model to the unconstrained model.

Variants for which all three likelihood-ratio tests showing P < 0.05 were classified as IBD-saturated, interpreted as associated with both CD and UC but with different effect sizes. Otherwise, the variant was assigned to the constrained model with the highest likelihood. Variants best explained by the IBD-unsaturated model were interpreted as associated with both CD and UC but with no evidence for different effect sizes. For variants where the model could not converge (N = 1) or not reaching P < 10-4 in the EUR tier 1 cohort (N = 308), we assigned the phenotype (IBD, CD or UC) based on the most significant result in the EUR tier 2 summary statistics.

### Heritability analyses

We used LD Score Regression (LDSC)^13^ to estimate the total heritability explained by the variants included in this study (hg2). We first estimated univariate LD Scores for all the variants reported in >50% of the total effective sample size (Neff), MAF >= 0.001 and imputed with INFO ≥ 0.9 in the EUR tier 2 analysis (IBD N = 12,674,691; CD N = 12,840,263; UC N = 12,833,584). The union of these variants (N = 12,966,826) is referred to hereafter as the EUR tier 2 variant set.

As an LD reference panel, we used genotype data from EUR participants (tier 1), applying a 1 Mb window for LD score calculation. To validate the IIBDGC-derived LD scores, we compared them with LD scores generated from UK Biobank (UKB) data released through the Pan-UKB project^14^, restricting the comparison to HapMap3 variants. The LD scores showed excellent concordance (Pearson r = 0.99). We additionally compared LD scores computed using all EUR samples with scores derived solely from individuals genotyped on the GSA array, observing similarly high concordance (Pearson r = 0.99).

To infer hg2 on the liability scale, we used disease prevalence estimates of 0.7%, 0.3% and 0.4% for IBD, CD and UC, respectively, as previously reported^4^, and specified the sample size as the Neff, such that the sample prevalence was set to 0.5. We evaluated hg2 across multiple minor allele frequency (MAF) thresholds to assess whether inclusion of lower-frequency variants might inflate heritability estimates. These analyses showed no evidence of inflation attributable to rare variant inclusion and indicated that most of the observed genetic variance is explained by common variants (Extended Data Fig. 4). LDSC intercepts and ratios ((intercept – 1) / (mean(χ²) – 1)) also showed no evidence of genomic inflation beyond that expected from polygenicity (Source data)^13^.

To estimate the proportion of SNP heritability explained by variants within IBD-associated regions, we generated two genomic annotations: one containing variants located within the IBD regions defined above (N = 420) and a second containing all remaining variants in the genome. These IBD regions encompassed 19% of the genome (assuming a total genome length of 3,039 Mb, GRCh38.p14) and contained 21% of all variants. We repeated this analysis with the regions that only contained newly reported association signals (‘new regions’); whereas regions harbouring either only known or known and newly identified independent signals were conservatively classified as ‘known regions’. We then applied stratified LD Score Regression (S-LDSC)^15^, using the implementation provided within PolyFun, to partition SNP heritability between these two annotations.

We additionally estimated the genetic variance explained per variant, and evaluated the sample size required to detect additional variants that explain lower ranges in genetic variance (Supplementary Note 5).

### Genetic correlation analysis

We estimated genetic correlations between pairs of traits using the LDSC implementation for cross-trait LD Score Regression. For this analysis, we restricted to a subset of EUR tier 2 variants that met the following criteria: variants located outside the HLA region, single nucleotide polymorphisms (SNPs), and non-palindromic sites to minimise potential strand alignment errors across datasets. This resulted in 9,985,018 variants for IBD, 10,107,872 for CD and 10,106,917 for UC.

We next selected traits with publicly available GWAS summary statistics from the GWAS Catalog^16^. For each trait, we included the largest available GWAS or meta-analysis based on European (or predominantly European) ancestry and required that summary statistics included effect and non-effect alleles, allele frequency, effect size and standard error. Source URLs for all studies are provided in the Source Data.

Because many non-IBD traits exhibited substantial correlation (e.g. HDL) we controlled for multiple testing using false discovery rate (FDR) adjustment applied to LDSC-derived P values (R package *qvalue*). Traits were considered to show significant genetic correlation when q < 0.01.

### Latent causal modelling to establish causal relationships between genetic correlated traits

We implemented a latent causal variable (LCV) modelling approach^17^ to infer whether the pairs of traits with significant genetic correlation exhibited full or partial genetic causality. For this analysis, we used the same subset of variants and LD scores applied in the cross-trait LDSC analyses, excluding traits with weak heritability estimates (LCV h² Z-score < 7), as recommended by the authors. Pairs of traits showing significant evidence of full or near-full genetic causality were defined as those with q < 0.05 and |gĉp| > 0.6.

### Enrichment analysis

To identify cell types and tissues that play a role in IBD, CD and UC disease susceptibility, we applied S-LDSC (the version provided with PolyFun^18^) to partition heritability across a number of cell-type specific genomic annotations selected in Weisbrood et al.^18^ as the baseline model (Supplementary Note). In addition to these annotations, we incorporated ATAC-seq data from immune cell types under resting and stimulated conditions^19^; cis candidate regulatory elements (cCRE) derived from single cell ATAC-seq data from 30 adult tissues^20^; and single-cell open chromatin assays across eight different intestinal sites^21^, as well as ATAC-seq, intact-HiC and DNase data from ENCODE portal Immune Cell collection. For each annotation, we estimated per-annotation LD scores using the EUR tier 2 variants (as defined above) and the genotype data from the EUR participants, setting up the LD window to 1Mb, as LD reference.^22,23^

The additional cell-type specific features were incorporated into the baseline model as quantitative traits one at a time, so we could infer and compare the amount of heritability captured by each cell type and annotation, independently. To correct for multiple testing, we applied bonferroni correction, accounting for the total number of features used (N = 689; 103 of them from the baseline model), and the number of disease phenotypes (N = 3 ;P ≤ 2.42×10^-5^).

We used the coefficient Z-scores estimated by S-LDSC to compare the relative contribution of different cell types to the two major IBD subtypes, CD and UC (Supplementary Note).

### Fine-mapping

To identify regions with sufficient power to yield high-confidence fine-mapping results, we evaluated all 420 regions and assessed whether any genome-wide significant variants (from meta analysis of the subset of studies with genotyping array data available, named EUR tier 1) remained after excluding variants that failed two criteria: effective sample coverage below 95% and MAF below 1% (Extended Data Fig. 3). A more stringent sample coverage and MAF threshold was applied for fine-mapping than for GWAS to minimize potential biases, as variants with low frequency or insufficient sample representation can distort the fine-mapping posterior distribution and lead to false positive signals. After this filtering step, 187 regions still contained significant variants and were taken forward for fine-mapping. The remaining regions were not excluded due to lack of GWAS evidence, but rather because limited power in the current dataset precluded reliable fine-mapping, and therefore they were not expected to yield well-resolved credible sets.

Fine-mapping was conducted separately for each phenotype (CD, UC, and IBD) within each region using SuSiE^24^. After fine-mapping, we excluded credible sets whose lead variants had both low sample coverage (<95%) and low minor allele frequency (MAF) (<1%). In addition, we excluded credible sets whose lead variants didn’t reach genome-wide significance (p>=5E-8).

For regions retaining credible sets after filtering, we performed a credible set alignment analysis following the method described by ref^25^. Briefly, we calculated an alpha-weighted Jaccard similarity index between all pairs of credible sets, where the alpha values are derived from SuSiE and measure the posterior inclusion probability of each variant. Next, we performed hierarchical clustering using 1 minus the similarity index as the distance metric and cut the dendrogram at a height of 0.9. Any pairs of credible sets with a similarity index greater than 0.1 were merged. We further merged credible sets if the linkage disequilibrium (LD; r^2) between their lead variants was greater than or equal to 0.8. Phenotypes were reassigned for merged credible sets using the following rules:

- If only one phenotype (IBD, CD, or UC) was present, the original phenotype was retained.
- If both CD (or UC) and IBD were present, the phenotype was reassigned to CD (or UC).
- If all three phenotypes were present, the phenotype was reassigned to IBD.

Next, we used GCTA-COJO^10^ to calculate the conditional P for the lead variant of each credible set, using GSA genotype data as the reference panel, as this dataset represents the largest proportion of our EUR tier 1 samples.

Of the 187 regions in which fine-mapping was performed, one didn’t produce any credible set because it was automatically filtered out by SuSiE due to the purity requirement. In an additional 16 regions, although fine-mapping signals were initially detected, they were removed after further filtering out variants with low sample coverage and low MAF (Supplementary Data Fig. 3). Across the remaining 170 regions, we identified a total of 268 credible sets.

We also evaluated how well the independent signals identified by fine mapping are captured by the COJO conditional analyses. Of the 268 credible sets, 249 were captured by conditional analysis (lead variants r² > 0.1; Supplementary Table 11), 216 of them in high linkage disequilibrium (r² > 0.6).

### SPARC IBD pQTL analyses

Matched Olink Explore 3072 Panel and GSA data for 1,021 IBD cases from the SPARC IBD cohort1 were used to calculate pQTL. After TOPMed imputation^3^, genetic data QC matched the methods from Sun et al., leaving 6.7 million variants and 1,021 individuals that passed filters. Regenie^4^ was next used to identify pQTL including age, age of IBD diagnosis, sex, IBD classification, race, duration of disease, disease severity, and the first 20 genetic PCs as covariates in each step. For the first step a more stringent LD pruning was applied which resulted in 120,000 variants, then step 2 was run using batches containing 100 genes and 1 chromosome each for the full set of 6.7 million variants and 2,923 Olink genes which passed QC.

### Effector gene determination

#### Co-localization analysis

We ran pairwise colocalisation analyses between our IBD CD and UC GWAS summary statistics (EUR tier 2) and eQTL, sc-eQTL and cis-pQTL summary statistics using coloc v5.2.2^26–28^ and the coloc.abf function. We included gut and blood sc-eQTL data^29^, eQTL studies carried out in immune related cell types^30^, or relevant tissues from eQTL catalogue^31^, and three pQTL datasets, SPARC IBD, analysed specifically as part of this study, and UKB and Decode described in^32^. For the pQTL datasets, we restricted the analyses to the 2Mb cis window around the TSS from each protein coding gene targeted in the assay. We only reported colocalisations between GWAS and e/pQTLs (i) showing a posterior probability (PP) of both traits sharing the same causal variant (PP H4) ≥ 0.8; and (ii) where the lead variant for our independent IBD, CD and UC signals were in strong LD (r2 > 0.6) with the lead e/pQTL variant.

#### Prioritization of co-localization results

To weight each gene nomination according to tissue or cell-type specificity and biological consequence (eQTL or pQTL), we estimated heritability-based enrichment scores for each cell type or condition within each IBD subtype. For each eQTL or pQTL dataset (one condition at a time), we computed enrichment Z-scores using S-LDSC, annotating the lead index variant significantly associated with gene expression or plasma protein levels and extending this annotation to variants in LD with the index variant. To assess robustness to LD definition, we evaluated the stability of the Z-score estimates at different LD thresholds (Supplementary Note). On this basis, we selected an LD threshold of r² = 0.25 for all subsequent analyses.

We assigned a priority rank to each signal-gene pair based on the maximum tissue enrichment heritability Z-score observed across all colocalizing QTLs. To benchmark this prioritization strategy, we used burden heritability estimates as an orthogonal reference metric^33^.

For signals with multiple colocalized genes, the concordance between tissue based and burden heritability based rankings was assessed using a Fisher’s exact test (Extended Data Fig. 5a). We separately assessed the performance of our combined approach, that is genes assigned the highest tissue-based rank (rank 1), together with genes nominated by confirmed exonic variants, and genes nominated by MR with regard to the closest gene. We additionally compared the mean burden heritability of all colocalized genes, and colocalized genes assigned the highest tissue-based rank (rank 1). For each group, a background distribution was generated by randomly selecting the same number of genes (except for *NOD2*) within ±1 Mb of the original set and recalculating mean burden heritability 1,000,000 times; the proportion of replicates exceeding the observed value was taken as the empirical P value.

#### Evaluating the directionality of effect across different tissues

Although colocalisation can infer shared causal signals with different levels of missingness in the GWAS and QTL studies, aligning effect direction requires variants present in both datasets, and may be complicated by allele-orientation mismatches (particularly at palindromic variants). To systematically address these limitations, we (i) expanded the variant set to include those in strong LD (r2 ≥ 0.6) with the GWAS lead variant; (ii) estimated, for each variant, the relationship between the eQTL effect (βxy) and the GWAS effect (βxz) using a Wald ratio test (βwald = βxy / βxz); and (iii) calculated the mean βwald and 95% across all variants in LD.

We also examined colocalized signals where the GWAS lead variant was present in the QTL dataset (85% of results), and observed strong agreement between the mean βwald and the lead variant βwald (Pearson r = 0.97; P < 1×10⁻^16^) with only 0.13% showing discordant directions, supporting the robustness of this approach.

#### Mendelian Randomization

To nominate additional effector genes for our GWAS, we implemented a Mendelian randomization approach via GSMR2^10^, integrating 3 pQTL datasets, SPARC IBD, and UKB and Decode^32^.

For this approach we excluded palindromic variants to ensure that, through comparing the MAF, the effect allele from both datasets aligned. We also only focused on protein coding genes within the 420 IBD regions as defined above, that were assayed by the pQTL datasets (N Decode = 1872, N UKB and SPARC IBD = 1140).

We defined the GSMR2 parameters as follows, P threshold of 0.01 for single and multiple SNP-based HEIDI-outlier analysis; a minimum number of instruments required for the analysis to 10; an LD r2 threshold of 0.01 to remove SNPs in high LD; and the FDR threshold of 0.01 to remove the chance correlations between the SNP instruments. Only variants with MAF ≥ 0.0025 were included. GSMR2 was run with the new HEIDI-outlier filtering method, designed to be more robust against directional pleiotropy than the original implementation.

We also applied a reverse MR, informed by the IBD independent index variants, and only reported as candidate effector genes those with significant evidence in the forward analyses (q value MR < 0.05) and no evidence of reverse causation (q value reverse MR ≥ 0.05).

#### Exome-wide association results integration

We annotated the independent association signals as likely driven by coding variants, when those variants reported previously by the previous IIBDGC Exome Sequencing study^33^ were also included in (i) lead index variants as reported through COJO; (ii) when the lead variant from the independent signals was in high LD (Supplementary Table 5).

#### Gene enrichment analyses

To annotate the list of prioritised genes into biological programmes, we ran a hypergeometric test to identify a significant overrepresentation of our list of genes among pathways and Gene Ontology terms as annotated in the pathway gene set database (http://download.baderlab.org/EM_Genesets/; release dated March 2026), subsetting this full list to pathways including between 2 and 2000 genes. We manually curated the list of genes reported in ref, as significantly dysregulated upon *ETS2* overexpression^34^.

We annotated whether IBD genes participate in protein complexes [https://mips.helmholtz-muenchen.de/corum/download].

We further evaluated whether the prioritised genes were enriched for: (i) curated receptor–ligand pairs from CellTalkDB (http://tcm.zju.edu.cn/celltalkdb/); (ii) genes in which high-penetrance mutations have been implicated in monogenic forms of inflammatory bowel disease (IBD)^35^; and (iii) an extended set of clinically relevant IBD monogenic and primary immunodeficiency (PID) genes curated by Genomics England PanelApp (panel 398 v8.0; https://panelapp.genomicsengland.co.uk/panels/398/). Enrichment was assessed using a permutation-based framework in which an equal number of protein-coding genes were randomly sampled from GENCODE v44, repeated 500,000 times. Empirical P values were defined as the proportion of permutations in which the number of receptor–ligand pairs or gene overlaps (monogenic or PID genes) exceeded or equalled that observed in the prioritised gene set. Odds ratios were calculated as the observed number of pairs or overlaps divided by the mean number observed across permutations.

#### Previous evidence from Open Targets

We retrieved information on existing genetic evidence supporting IBD genes through https://platform.opentargets.org/ (association_by_datasource_direct; release “March 2026 (26.03)”).

#### Drug target identification

Drug target information was compiled using the ChEMBL Evidence and Drug-Mechanism of Action databases available from https://platform.opentargets.org/ (release 26.03). Drug names for the top clinical trial for each gene were then matched against the Drug - Mechanism of Action database to identify the mechanisms of action for each reported in Supplementary Table 10. Genetic prediction scores extracted from Chen et al.^36^, were assigned to the genes.

## Acknowledgements

A complete list of acknowledgments, funding sources and relevant Ethics Committee or Institutional Review Board Information appears in the Supplementary Note.

## Data availability

Genotype data from UK IBDGC that support this study have been deposited in the European Genome-phenome Archive (EGA), which is hosted by the EBI and the CRG, and are available under accessions EGAD00000000005, EGAS00000000084, and EGAS00001000924. Further information about EGA can be found at https://ega-archive.org and The European Genome-phenome Archive in 2021: 10.1093/nar/gkab1059. These data can be accessed following approval by the Sanger Data Access Committee.

Genotype data used in this study has been made publicly available in dbGaP Study Accession: phs001642.v1.p1 - Center for Common Disease Genomics [CCDG] - Autoimmune: Inflammatory Bowel Disease (IBD) Exomes and Genomes (https://www.ncbi.nlm.nih.gov/projects/gap/cgi-bin/study.cgi?study_id=phs001642.v1.p1).

Genotype and phenotype participant data with identifiers from IBDBR are available via the NIHR BioResource (https://www.ibdbioresource.nihr.ac.uk/), subject to approval of a research proposal by the NIHR BioResource Data Access Committee and the completion of a signed data access agreement.

Access to the International IBDGC (IIBDBC) phenotype data collection can be requested following approval by the IIBDGC Management Committee (https://www.ibdgenetics.org/).

## Main References

1. Hracs, L. et al. Global evolution of inflammatory bowel disease across epidemiologic stages. Nature 642, 458–466 (2025).

2. Azabdaftari, A., Jones, K. D. J., Kammermeier, J. & Uhlig, H. H. Monogenic inflammatory bowel disease-genetic variants, functional mechanisms and personalised medicine in clinical practice. Hum Genet 142, 599–611 (2023).

3. Wellcome Trust Case Control Consortium. Genome-wide association study of 14,000 cases of seven common diseases and 3,000 shared controls. Nature 447, 661–678 (2007).

4. Bolton, C. et al. An Integrated Taxonomy for Monogenic Inflammatory Bowel Disease. Gastroenterology 162, 859–876 (2022).

5. de Lange, K. M. et al. Genome-wide association study implicates immune activation of multiple integrin genes in inflammatory bowel disease. Nat Genet 49, 256–261 (2017).

6. Huang, H. et al. Fine-mapping inflammatory bowel disease loci to single-variant resolution. Nature 547, 173–178 (2017).

7. Sazonovs, A. et al. Large-scale sequencing identifies multiple genes and rare variants associated with Crohn’s disease susceptibility. Nat Genet 54, 1275–1283 (2022).

8. Liu, Z. et al. Genetic architecture of the inflammatory bowel diseases across East Asian and European ancestries. Nat Genet 55, 796–806 (2023).

9. Minikel, E. V., Painter, J. L., Dong, C. C. & Nelson, M. R. Refining the impact of genetic evidence on clinical success. Nature 629, 624–629 (2024).

10. Wang, G., Sarkar, A., Carbonetto, P. & Stephens, M. A simple new approach to variable selection in regression, with application to genetic fine mapping. J R Stat Soc Series B Stat Methodol 82, 1273–1300 (2020).

11. Zou, Y., Carbonetto, P., Wang, G. & Stephens, M. Fine-mapping from summary data with the ‘Sum of Single Effects’ model. PLoS Genet 18, e1010299 (2022).

12. Bulik-Sullivan, B. K. et al. LD Score regression distinguishes confounding from polygenicity in genome-wide association studies. Nat Genet 47, 291–295 (2015).

13. Finucane, H. K. et al. Partitioning heritability by functional annotation using genome-wide association summary statistics. Nat Genet 47, 1228–1235 (2015).

14. Zhu, R. et al. Exome sequencing directly implicates 68 genes in inflammatory bowel disease. medRxiv (2026) doi:10.64898/2026.05.08.26352648.

15. Raffals, L. E. et al. The Development and Initial Findings of A Study of a Prospective Adult Research Cohort with Inflammatory Bowel Disease (SPARC IBD). Inflamm Bowel Dis 28, 192–199 (2022).

16. Sun, B. B. et al. Plasma proteomic associations with genetics and health in the UK Biobank. Nature 622, 329–338 (2023).

17. Buniello, A. et al. Open Targets Platform: facilitating therapeutic hypotheses building in drug discovery. Nucleic Acids Res 53, D1467–D1475 (2025).

18. Ishida, T. et al. Endothelial lipase is a major determinant of HDL level. J Clin Invest 111, 347–355 (2003).

19. Dong, J. et al. Adenovirus-mediated overexpression of sphingomyelin synthases 1 and 2 increases the atherogenic potential in mice. J Lipid Res 47, 1307–1314 (2006).

20. Mossotto, E. et al. Evidence of a genetically driven metabolomic signature in actively inflamed Crohn’s disease. Sci Rep 12, 14101 (2022).

21. Hickey, J. W. et al. Organization of the human intestine at single-cell resolution. Nature 619, 572–584 (2023).

22. Del Sordo, R., Lupinacci, G., Tanzi, G., Bassotti, G. & Villanacci, V. Imatinib and Dasatinib-induced Ulcerative Colitis: Case Report. Inflamm Bowel Dis 28, e1–e2 (2022).

23. Ma, Z. et al. Imatinib-induced ulcerative colitis. J Oncol Pharm Pract 30, 1111–1117 (2024).

24. Sandner, L. et al. The Tyrosine Kinase Tec Regulates Effector Th17 Differentiation, Pathogenicity, and Plasticity in T-Cell-Driven Intestinal Inflammation. Front Immunol 12, 750466 (2021).

25. Khoury, H. J., Gambacorti-Passerini, C. & Brümmendorf, T. H. Practical management of toxicities associated with bosutinib in patients with Philadelphia chromosome-positive chronic myeloid leukemia. Ann Oncol 29, 578–587 (2018).

26. Yamauchi, K. et al. Dasatinib-induced colitis: clinical, endoscopic and histological findings. Scand J Gastroenterol 57, 449–456 (2022).

27. Rosen, J. D., Broadaway, K. A., Brotman, S. M., Mohlke, K. L. & Love, M. I. Higher eQTL power reveals signals that boost GWAS colocalization. bioRxiv (2025) doi:10.1101/2025.08.05.668745.

28. Henry, A. et al. Single-cell genetics identifies cell type-specific causal mechanisms in complex traits and diseases. medRxiv (2025) doi:10.1101/2025.08.28.25334614.

29. Alegbe, T. et al. Cell-type-resolved genetic regulatory variation shapes inflammatory bowel disease risk. medRxiv (2025) doi:10.1101/2025.06.24.25330216.

30. Fulco, C. P. et al. Systematic mapping of functional enhancer-promoter connections with CRISPR interference. Science 354, 769–773 (2016).

31. Xie, S., Armendariz, D., Zhou, P., Duan, J. & Hon, G. C. Global Analysis of Enhancer Targets Reveals Convergent Enhancer-Driven Regulatory Modules. Cell Rep 29, 2570–2578.e5 (2019).

32. Gasperini, M. et al. A Genome-wide Framework for Mapping Gene Regulation via Cellular Genetic Screens. Cell 176, 377–390.e19 (2019).

33. Boyle, E. A., Li, Y. I. & Pritchard, J. K. An Expanded View of Complex Traits: From Polygenic to Omnigenic. Cell 169, 1177–1186 (2017).

34. Liu, X., Li, Y. I. & Pritchard, J. K. Trans Effects on Gene Expression Can Drive Omnigenic Inheritance. Cell 177, 1022–1034.e6 (2019).

35. Wray, N. R., Wijmenga, C., Sullivan, P. F., Yang, J. & Visscher, P. M. Common Disease Is More Complex Than Implied by the Core Gene Omnigenic Model. Cell 173, 1573–1580 (2018).

36. Jagadeesh, K. A. et al. Identifying disease-critical cell types and cellular processes by integrating single-cell RNA-sequencing and human genetics. Nat Genet 54, 1479–1492 (2022).

37. Krzak, M. et al. Single-Cell RNA Sequencing of Terminal Ileal Biopsies Identifies Signatures of Crohn’s Disease Pathogenesis. medRxiv 2023.09.06.23295056 (2024) doi:10.1101/2023.09.06.23295056.

38. Vermeire, S., Van Assche, G. & Rutgeerts, P. Laboratory markers in IBD: useful, magic, or unnecessary toys? Gut 55, 426–431 (2006).

39. Ritchie, S. C. et al. The Biomarker GlycA Is Associated with Chronic Inflammation and Predicts Long-Term Risk of Severe Infection. Cell Syst 1, 293–301 (2015).

40. Dierckx, T., Verstockt, B., Vermeire, S. & van Weyenbergh, J. GlycA, a Nuclear Magnetic Resonance Spectroscopy Measure for Protein Glycosylation, is a Viable Biomarker for Disease Activity in IBD. J Crohns Colitis 13, 389–394 (2019).

41. Cartier, A. & Hla, T. Sphingosine 1-phosphate: Lipid signaling in pathology and therapy. Science 366, (2019).

## Methods references

1. Karczewski, K. J. et al. The mutational constraint spectrum quantified from variation in 141,456 humans. Nature 581, 434–443 (2020).

2. Taliun, D. et al. Sequencing of 53,831 diverse genomes from the NHLBI TOPMed Program. Nature 590, 290–299 (2021).

3. Das, S. et al. Next-generation genotype imputation service and methods. Nat Genet 48, 1284–1287 (2016).

4. Mbatchou, J. et al. Computationally efficient whole-genome regression for quantitative and binary traits. Nat Genet 53, 1097–1103 (2021).

5. Willer, C. J., Li, Y. & Abecasis, G. R. METAL: fast and efficient meta-analysis of genomewide association scans. Bioinformatics 26, 2190–2191 (2010).

6. Liu, Z. et al. Genetic architecture of the inflammatory bowel diseases across East Asian and European ancestries. Nat Genet 55, 796–806 (2023).

7. Mägi, R. et al. Trans-ethnic meta-regression of genome-wide association studies accounting for ancestry increases power for discovery and improves fine-mapping resolution. Hum Mol Genet 26, 3639–3650 (2017).

8. Huang, H. et al. Fine-mapping inflammatory bowel disease loci to single-variant resolution. Nature 547, 173–178 (2017).

9. de Lange, K. M. et al. Genome-wide association study implicates immune activation of multiple integrin genes in inflammatory bowel disease. Nat Genet 49, 256–261 (2017).

10. Yang, J. et al. Conditional and joint multiple-SNP analysis of GWAS summary statistics identifies additional variants influencing complex traits. Nat Genet 44, 369–75, S1–3 (2012).

11. Quinlan, A. R. & Hall, I. M. BEDTools: a flexible suite of utilities for comparing genomic features. Bioinformatics 26, 841–842 (2010).

12. Liu, J. Z. et al. Association analyses identify 38 susceptibility loci for inflammatory bowel disease and highlight shared genetic risk across populations. Nat Genet 47, 979–986 (2015).

13. Bulik-Sullivan, B. K. et al. LD Score regression distinguishes confounding from polygenicity in genome-wide association studies. Nat Genet 47, 291–295 (2015).

14. Karczewski, K. J. et al. Pan-UK Biobank genome-wide association analyses enhance discovery and resolution of ancestry-enriched effects. Nat Genet 57, 2408–2417 (2025).

15. Finucane, H. K. et al. Partitioning heritability by functional annotation using genome-wide association summary statistics. Nat Genet 47, 1228–1235 (2015).

16. Sollis, E. et al. The NHGRI-EBI GWAS Catalog: knowledgebase and deposition resource. Nucleic Acids Res 51, D977–D985 (2023).

17. O’Connor, L. J. & Price, A. L. Distinguishing genetic correlation from causation across 52 diseases and complex traits. Nat Genet 50, 1728–1734 (2018).

18. Weissbrod, O. et al. Functionally informed fine-mapping and polygenic localization of complex trait heritability. Nat Genet 52, 1355–1363 (2020).

19. Calderon, D. et al. Landscape of stimulation-responsive chromatin across diverse human immune cells. Nat Genet 51, 1494–1505 (2019).

20. Zhang, K. et al. A single-cell atlas of chromatin accessibility in the human genome. Cell 184, 5985–6001.e19 (2021).

22. ENCODE Project Consortium. An integrated encyclopedia of DNA elements in the human genome. Nature 489, 57–74 (2012).

23. Luo, Y. et al. New developments on the Encyclopedia of DNA Elements (ENCODE) data portal. Nucleic Acids Res 48, D882–D889 (2020).

24. Wang, G., Sarkar, A., Carbonetto, P. & Stephens, M. A simple new approach to variable selection in regression, with application to genetic fine mapping. J R Stat Soc Series B Stat Methodol 82, 1273–1300 (2020).

25. Kanai, M. et al. Insights from complex trait fine-mapping across diverse populations. medRxiv 2021.09.03.21262975 (2021) doi:10.1101/2021.09.03.21262975.

26. Giambartolomei, C. et al. Bayesian test for colocalisation between pairs of genetic association studies using summary statistics. PLoS Genet 10, e1004383 (2014).

27. Wakefield, J. Bayes factors for genome-wide association studies: comparison with P-values. Genet Epidemiol 33, 79–86 (2009).

28. Plagnol, V., Smyth, D. J., Todd, J. A. & Clayton, D. G. Statistical independence of the colocalized association signals for type 1 diabetes and RPS26 gene expression on chromosome 12q13. Biostatistics 10, 327–334 (2009).

30. Hu, S. et al. Inflammation status modulates the effect of host genetic variation on intestinal gene expression in inflammatory bowel disease. Nat Commun 12, 1122 (2021).

31. Kerimov, N. et al. A compendium of uniformly processed human gene expression and splicing quantitative trait loci. Nat Genet 53, 1290–1299 (2021).

32. Eldjarn, G. H. et al. Large-scale plasma proteomics comparisons through genetics and disease associations. Nature 622, 348–358 (2023).

33. Zhu, R. et al. Exome sequencing directly implicates 68 genes in inflammatory bowel disease. medRxiv (2026) doi:10.64898/2026.05.08.26352648.

34. Stankey, C. T. et al. A disease-associated gene desert directs macrophage inflammation through ETS2. Nature 630, 447–456 (2024).

35. Bolton, C. et al. An Integrated Taxonomy for Monogenic Inflammatory Bowel Disease. Gastroenterology 162, 859–876 (2022).

36. Chen, R. et al. Expanding drug targets for 112 chronic diseases using a machine learning-assisted genetic priority score. Nat Commun 15, 8891 (2024).

