## Extended_Data for "Resolving inflammatory bowel disease risk variants to genes and cell types"

Extended Data Fig 1 | Algorithm followed to define independent 2
Extended Data Fig 2 | European and multi-ancestry meta analyses effect size estimates 3
Extended Data Fig 3 | Overview of fine-mapping results for 187 regions. 4
Extended Data Fig 4 | Heritability analysis 5
Extended Data Fig 5 | Gene rankings based on tissue enrichment analyses 6
Extended Data Fig 6 | Traits showing significant genetic correlation with IBD, CD or UC 7
Extended Data Table 1 | Meta-analysis cohorts by PC-inferred ancestry 8

**Extended Data Fig 1 | Algorithm followed to define independent**

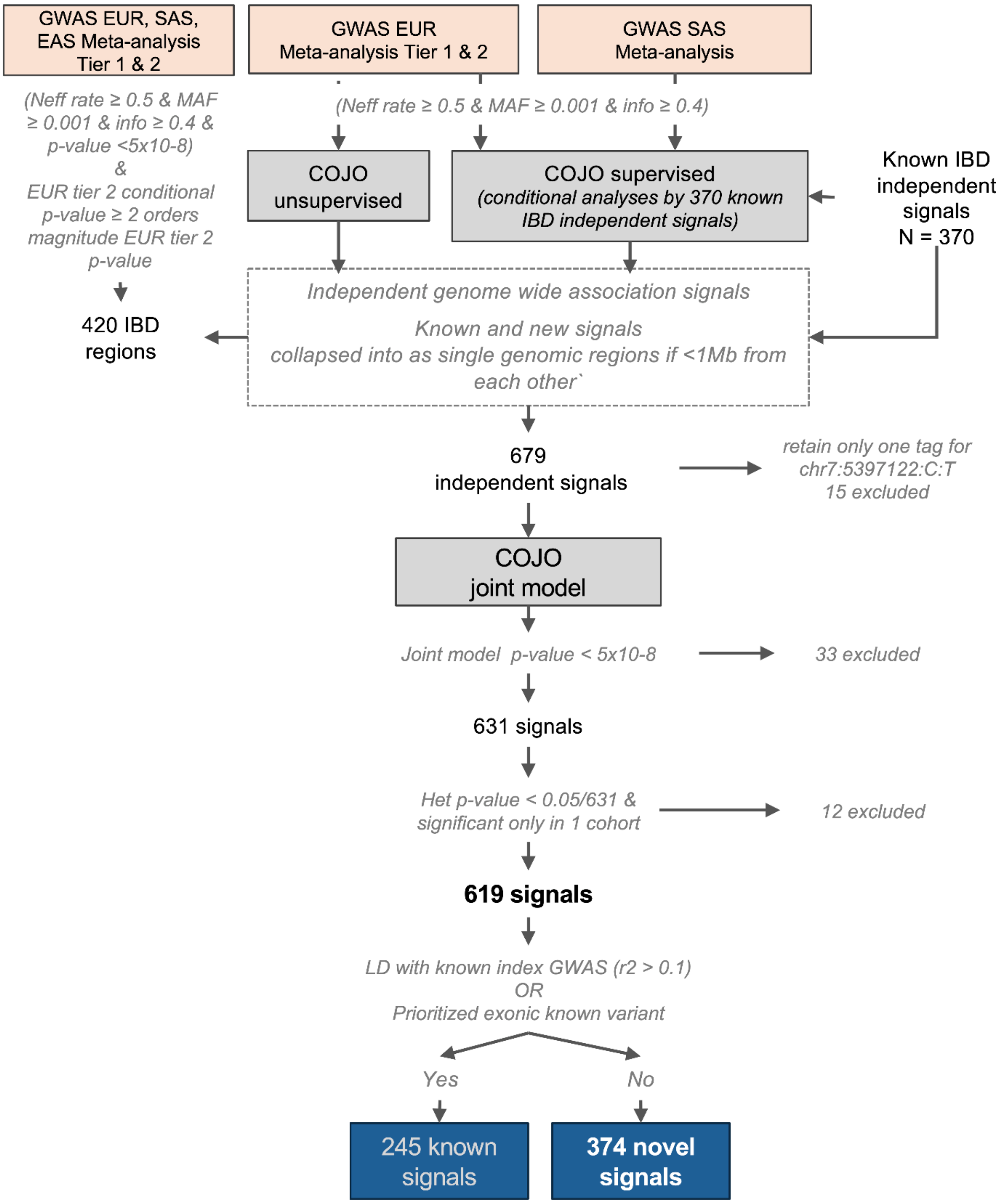

**Extended Data Fig 2 | European and multi-ancestry meta analysis effect size estimates**

x-axis: Z-score (β/SE) from the multi-ancestry meta-analysis.
**a,** y-axis: Z-score (β/SE) from the European meta-analysis. Dotted lines indicate ±4 s.d. of the mean distribution for the absolute difference between Z-scores (European versus multi-ancestry). Labels highlight five lead variants outside this interval, four of which (chr4:38333446:G:A, chr7:5433979:G:C, chr9:114801407:T:G, chr10:62710915:C:T) also show significant heterogeneity due to ancestry.
**b,** y-axis: $-\log_{10}(P)$ for ancestry-related heterogeneity estimated by MR-MEGA. The dotted horizontal line indicates the Bonferroni-corrected significance threshold ($0.05/619 = 8.08 \times 10^{-5}$). Labels highlight the four lead variants with the most significant heterogeneity across ancestries (out of 20 total).

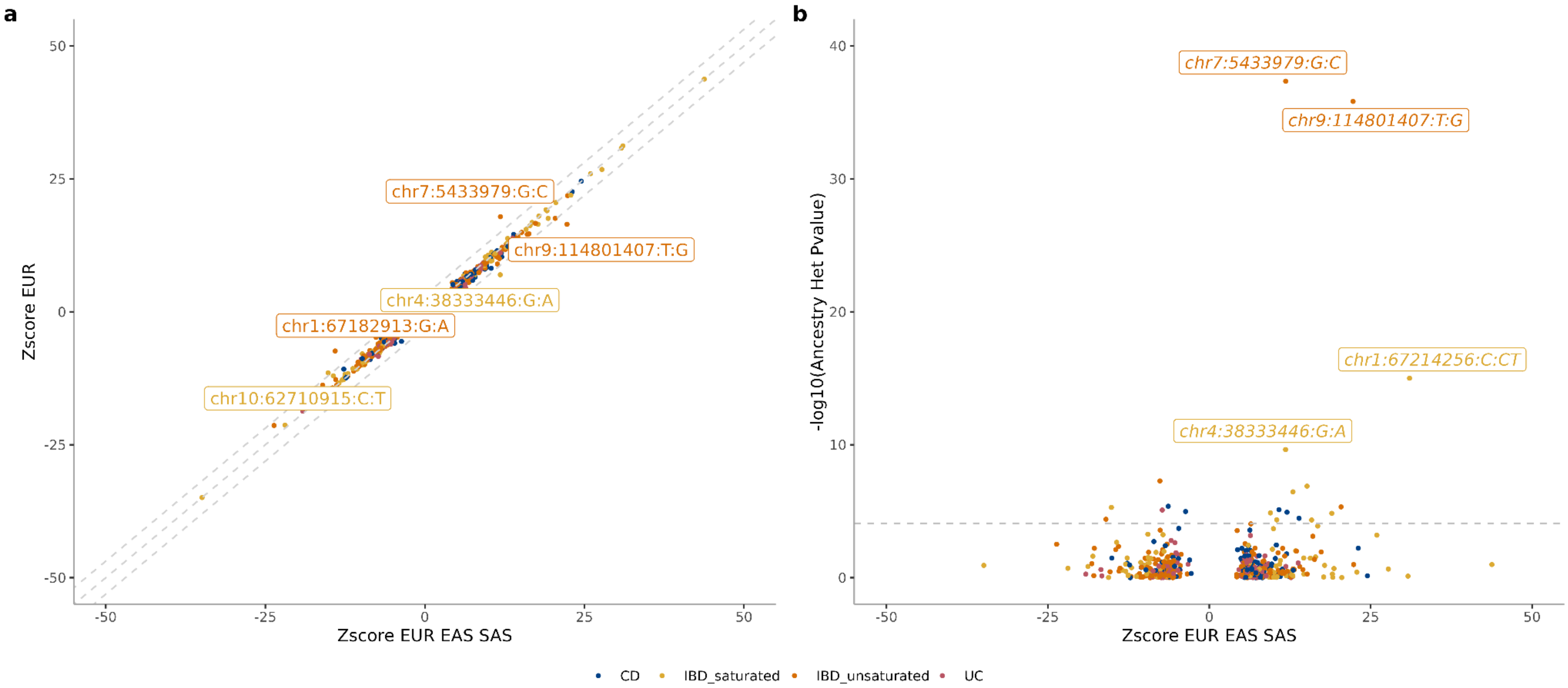

### Extended Data Fig 3 | Overview of fine-mapping results for 187 regions.

**a,** Summary of all 187 analyzed regions. **b,** Classification of the 17 regions without high-confidence fine-mapping signals. **c,** Example of a class B region. The dashed line represents the genome-wide significance threshold (p-value=5E-8). Each variant is colored by sample coverage: blue indicates high sample coverage, while red indicates low sample coverage. The shape of each variant denotes whether it was included in the initial credible set before post-filtering: squares represent variants being included in the credible set, and circles represent variants not included.

**a**

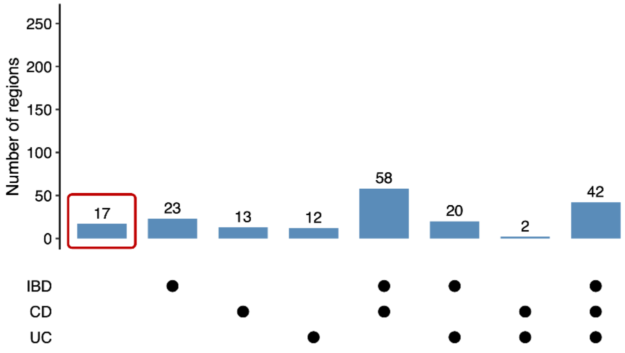


**c**

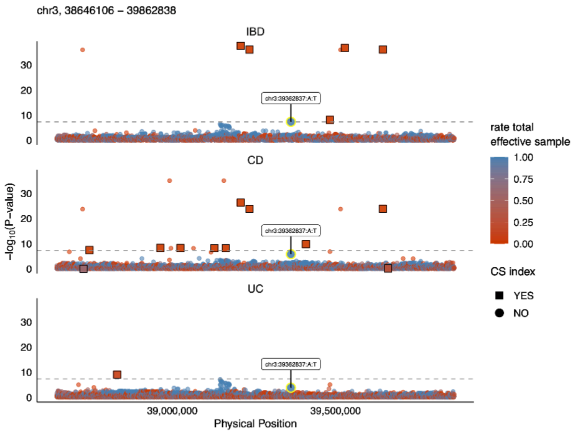


**b**

| Classification | Number of regions |
|---|---|
| Class A: no fine-mapping signal detected before filtering | 1 |
| Class B: no fine-mapping signal remained after filtering | 16 |

### Extended Data Fig 4 | Heritability analysis

Common based heritability estimates using LDSC (and S-LDSC)

a, b, common variant based heritability ($h_g^2$) on the liability scale estimates at different minor allele thresholds. a, the y axis shows the total phenotypic variance explained by the additive effects of the variants included in this study ($h_g^2$) at different minor allele frequency thresholds; error bars represent the $h_g^2$ standard error; b, number of variants contributing to the estimation of the the $h_g^2$ at each minor allele frequency threshold.

c, Stratified common variant-based heritability ($h_g^2$) estimates obtained after partitioning the genome into variants within IBD, CD and UC GWAS regions (in colour) versus variants outside those regions (grey). Old regions only include GWAS loci identified before this publication; 'all_regions' includes both old and new regions defined in this publication

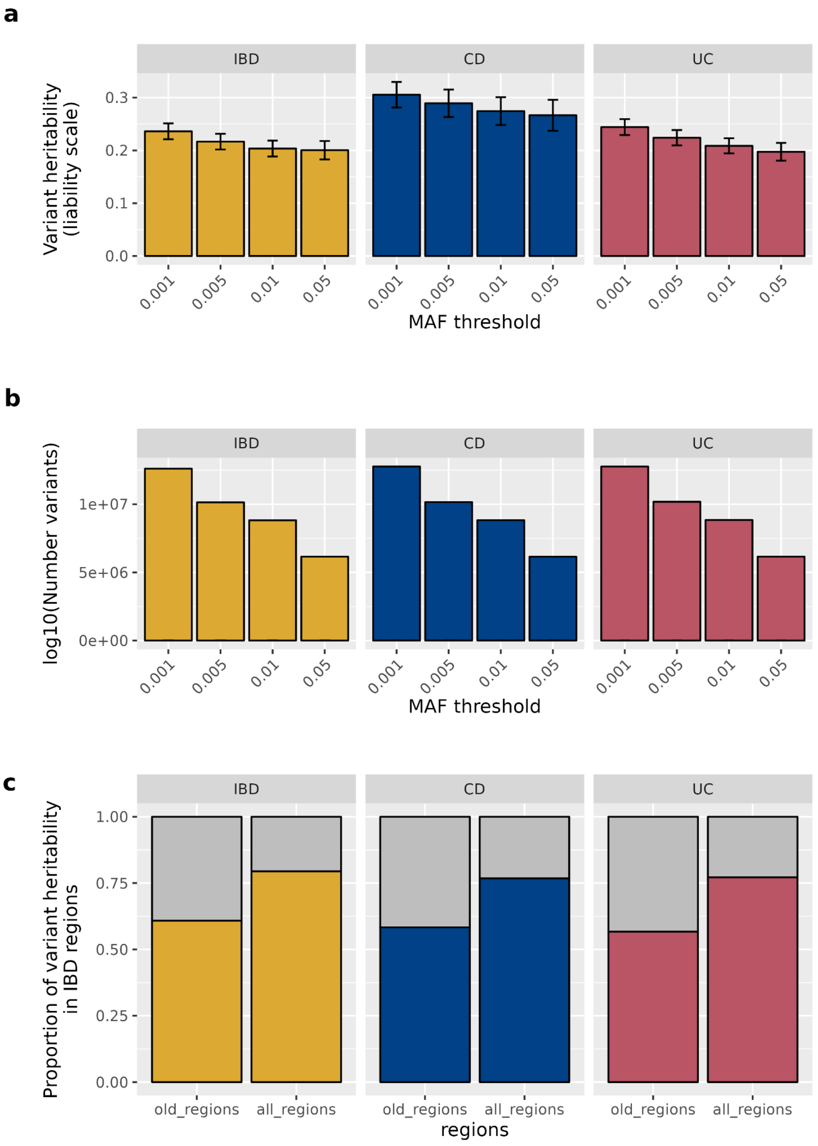

### Extended Data Fig 5 | Gene rankings based on tissue enrichment analyses

For signals with more than one nominated effector gene, we compared the rank assigned based on the eQTL tissue enrichment score (tissue_zscore) with the rank assigned based on the mean burden heritability estimates (burden_r2).

**a**: Rank reclassification between tissue enrichment scores and burden heritability. Fisher's exact test p-values are shown, from two-sided test.

**b-e**: The vertical line represents the mean burden heritability of genes nominated by, colocalization (rank 1 only), MR, and exonic variants, b; by closest gene, c; by all colocalization results, d; or by the subset of genes labelled as coloc_rank 1, e. The histogram represents the background mean heritability obtained by sampling the same number of genes 1,000,000 times.

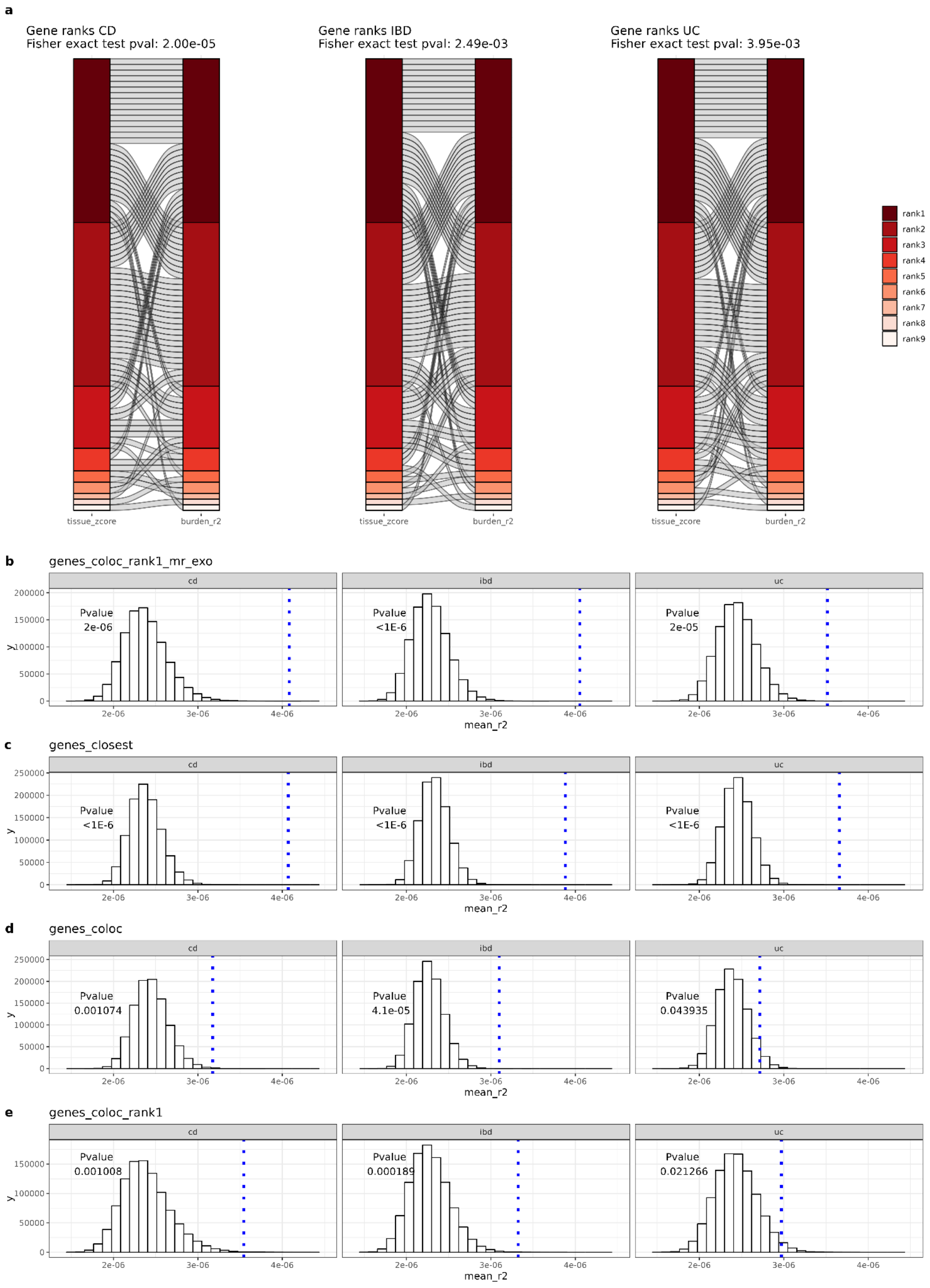

### Extended Data Fig 6 | Traits showing significant genetic correlation with IBD, CD or UC

Genetic correlation (x axis, rg),and rg 95% confidence interval (bars). In grey, pairs not reaching statistical significance (qvalue <0.01)

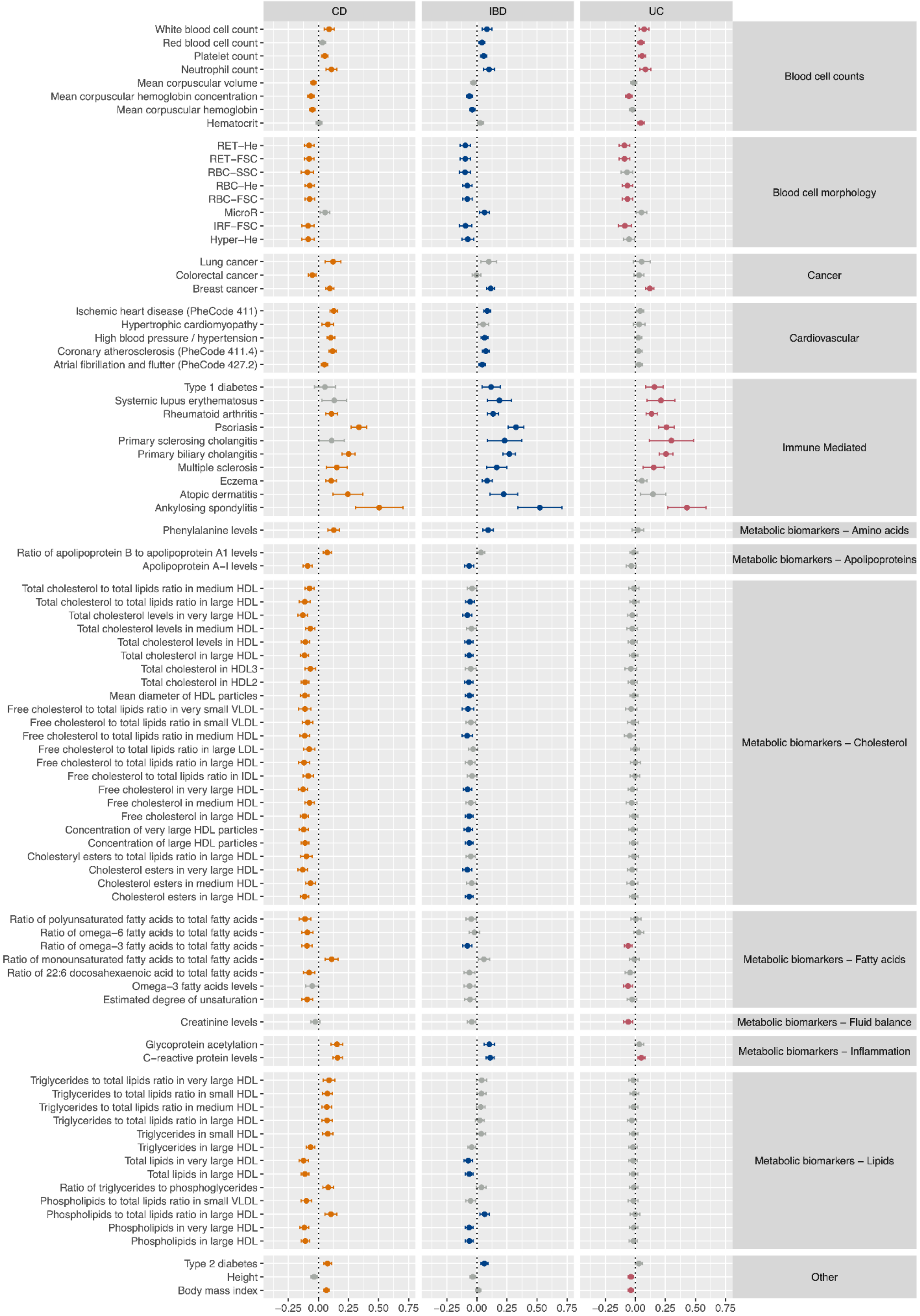

**Extended Data Table 1 | Meta-analysis cohorts by PC-inferred ancestry**

| Ancestries | Dataset | IBD | | CD | | UC | | Meta Analysis | | |
|---|---|---|---|---|---|---|---|---|---|---|
| | | Cases | Controls | Cases | Controls | Cases | Controls | | | |
| European | IIBDGC genotyped data meta-analysis | 62,501 | 40,060 | 35,560 | 37,863 | 25,132 | 39,248 | EUR Tier 1 | EUR Tier 2 | Multi ancestry |
| | UK Biobank | 8,392 | 412,120 | 1,897 | 412,120 | 4,678 | 412,120 | | | |
| | IBDBioResource | 20,914 | 35,379 | 10,074 | 35,379 | 10,178 | 35,379 | | | |
| | DAN-IBD | 3,433 | 1,628 | 1,549 | 1,628 | 1,603 | 1,628 | | | |
| | DeCode | 6,069 | 357,620 | 870 | 348,243 | 2,548 | 369,816 | | | |
| | FinnGen summary stats (r10) | 9,083 | 392,974 | 2,033 | 409,940 | 5,931 | 405,386 | | | |
| South Asian | UK Biobank | 160 | 6,166 | 36 | 6,166 | 108 | 6,166 | | SAS | |
| | IBDBioResource | 1,047 | 826 | 379 | 826 | 633 | 826 | | | |
| East Asian | EAS meta analysis (Liu, Liu, Gao et al. 2023) | 14,393 | 15,456 | 7,372 | 15,456 | 6,862 | 15,456 | | | |
| | **Total:** | **125,992** | **1,262,229** | **59,734** | | **57,565** | | | | |
| | **Total EUR only:** | **110,392** | **1,239,781** | **51,983** | | **50,070** | | | | |
| | **Total SAS only:** | **1,207** | **6,992** | **379** | | **633** | | | | |
| | **Total EAS only:** | **14,393** | **15,456** | **7,372** | | **6,862** | | | | |
