## Supplementary_Data for "Resolving inflammatory bowel disease risk variants to genes and cell types"


**Supplementary Note 1. Genotyping Array Cohorts, QC and analyses** **2**

- Genotyping data quality control 2
- Population ancestry definition and population PC estimation 3
- Imputation 3
- Analysis 4
- European Tier 1 meta-analysis 4
- Sex-Specific Analyses 4
- UK BioBank 5
  - Phenotype definitions 5
  - Evaluating the UKB case control definitions 6
  - UK Biobank Genotyping QC 7
- UK IBD BioResource 7
- FinnGen summary statistics 8
- deCODE summary statistics 8
- Dan-IBD summary statistics 9
- EUR Tier 1 and Tier 2 meta-analysis 9
- SAS tier 2 meta-analysis 9
- Trans-ancestry meta-analysis 10

**Supplementary Note 2. Sex specific results** **11**

**Supplementary Note 3. Heritability analyses informed by molecular QTL datasets** **12**

**Supplementary Note 4. Independent signals characterization** **15**

**Supplementary Note 5. Heritability captured per lead variant and power** **17**

**Supplementary Note 6 - S-LDSC analyses** **18**

**Supplementary Note 7. Explanation for replication failure of 21 high-PIP variants previous IBD fine-mapping analyses** **25**

**Supplementary Note 8. Replication of previous GWAS signals** **30**

**Supplementary Note 9. Acknowledgements** **32**

**Supplementary Note 10. IIBDGC patient-facing lay summary** **35**

**Supplementary Note 11. Lead variants showing significant ancestry-related heterogeneity in effect size, as estimated by MR-MEGA** **37**

### Supplementary Note 1. Genotyping Array Cohorts, QC and analyses

#### Genotyping data quality control

The DNA samples were genotyped across 59 cohorts with 9 different genotyping arrays (Supplementary Table 1).

Each cohort underwent the same quality control pipeline using a combination of Plink (v1.90b6.7 and v2.00a3LM)[1], bcftools (v1.16)[2], and KING (v2.2.4)[3]. In summary, we excluded (1) variants with low call rate (<0.95 for variants with MAF > 0.01; <0.98 for variants with MAF ≤ 0.01); (2) variants with a significant difference in genotype call rate missingness ($P < 1x10^{-4}$) between cases and controls; (3) variants with allele frequency differences versus those from the frequency reported in gnomAD (release v3.0) non-Finish Europeans, or TOPMed global MAF (using the criterion ((p1 − p0)^2/((p1+p0)*(2-p1-p0)) > 0.025 and > 0.125, respectively), where p0 is the MAF in the reference panel and p1 the observed MAF in the study); (4) variants with a Hardy–Weinberg equilibrium (HWE) P< 10-5 among controls and 10-12 among cases, estimated in EUR ancestry samples; and (5) monomorphic variants. For chromosome X, HWE was estimated in the subset of EUR ancestry individuals inferred as females by genotypic data.

We also excluded samples with a missing genotyping rate >0.05; a heterozygosity estimate +/- 4 standard deviations (SD) from the mean (per continental population, see below for details); or a mismatch between recorded gender and inferred genotypic sex. For this, we first identified, per cohort, a good set of ChrX and ChrY (when available in the array) markers (MAF > 0.01, genotyping call rate > 0.999 and HWE >10-4 for chrX variants, estimated in females; and genotyping call rate >0.995 for chrY variants, estimated in males, with no observed calls in females). We defined boundaries for inferred genotypic sex based on the +/- 4SD of the mean distribution of chrX F coefficient, as well as the number of ChrY genotype counts per recorded gender. We excluded samples with kinship coefficient ≥ 0.345 (defined using KING, within the cohort or between cohorts), and samples with evidence of non-European ancestry (which represented a small subset, approximately 5%, of the total number of samples included in this stage of the study).

**Population ancestry definition and population PC estimation**

To run the principal components analysis (PCA) for all participants, we combined all the cohorts (after excluding low call rate variants as well as samples with large missingness call rate). For this step, we used a set of variants available in all cohorts and in 1000GP, excluding variants associated with IBD susceptibility (P value $<10^{-4}$ in any of the cohorts), and variants located in any of the known long LD regions (as defined in https://github.com/meyer-lab-cshl/plinkQC/blob/master/inst/extdata/high-LD-regions-hg38-GRCh38.txt). This final list was pruned with the following parameters, window size = 50kb; step size = 5; r2 = 0.2. PCs were then estimated in 1000GP first, and 1000GP plus IIBDGC samples were projected afterwards.

With this information, the IIBDGC samples were assigned to global continental ancestries, and a new set of PCAs was run using only the IIBDGC samples, split by continental ancestry. The first EUR-PC was found to be informative to infer Ashkenazi Jewish (85% PPV; 95% NPV). Since we only had self-reported Ashkenazi ancestry for a subset of the IIBDGC samples, we used the EUR-PC1 to infer this EUR-specific ancestry across all the IIBDGC cohorts. The EUR samples were split accordingly into two groups (EUR-Ashkenaziy Jewish and EUR-non-Ashkenaziy) for the genotyping QC steps that were informed by population ancestry (see above).

Finally, to define the population ancestry PCs to be included in the analyses we ran a new round of PCAs per continental ancestry, and per genotyping array, first using only the non-duplicated IIBDGC samples, and projecting all IIBDGC samples (duplicates included) in a second stage. Loadings were visually inspected to confirm that none of the first 10 PCs were capturing long LD regions.

**Imputation**

Imputation of non-directly genotyped data was carried out per cohort, using the multi-ancestry TOPMed as reference panel (r2@1.0.0), through the TOPMed imputation server (imputationserver@1.5.7)[4]. After imputation, directly genotyped variants with imputation Empirical R < -0.5 were flipped, variants with Empirical Rsq ≤ 0.5 were excluded, and imputation was repeated. Post imputation, we excluded variants with strong deviation from HWE (PHWE ≤ 10-5 in controls; or alternatively ≤ 10-12 in cohorts that only included cases).

**Analysis**

Imputed data was combined by genotyping array (N = 9) and analysed together. First-degree relatives genotyped with different arrays were excluded from the analysis (kinship ≥0.177). The analyses were carried out using Regenie (v3.2.5)[5], applying Firth approximation correction to variants with P≤ 0.1, and including as covariates per-continental population ancestry PCs and sex.

**European Tier 1 meta-analysis**

We used METAL (version released on 2011-03-25) to perform a fixed-effects (standard error weighted) meta-analysis. Only variants with MAF ≥ 0.001 and imputation Rsq ≥ 0.4 per array (>13.9 million variants per array) were retained in each one of the meta-analyses (IBD, CD and UC). In total, we combined data from 102,561 individuals, 62,501 IBD participants (35,560 diagnosed with CD, 25,132 with UC), and 40,060 population controls (see Extended Data Table 1; Fig. 1).

**Sex-Specific Analyses**

Sex-specific analyses were performed for the imputed European Tier1 dataset, with exclusion of first-degree relatives across arrays, as described above. Analyses were carried out using Regenie (v3.4.1)[5], applying Firth approximation correction to variants with P≤ 0.1, with per-continental population ancestry PCs included as covariates. GWAMA (v2.2.2)[6] was used to perform fixed-effects meta-analyses, and variants with MAF ≥ 0.001 and imputation Rsq ≥ 0.4 per array were retained in each one of the meta-analyses (IBD, CD, and UC). From our combined data of 102,561 individuals, we had a total of 32,791 female and 29,710 male IBD participants, and 20,120 female and 19,940 male population controls. Four meta-analysis strategies are implemented in GWAMA: (1) sex-specific; (2) sex-combined; (3) sex-differentiated, which represents a combined P of male- and female-specific estimates while allowing for heterogeneity in effect between sexes (chi-squared distribution with 2 degrees of freedom [df]); and (4) sex-heterogeneity, a 1-df Cochran's Q-test. COJO[7] was used to identify independent signals (conditional P < 5 x 10-8) across the sex-specific summary statistics for IBD, CD, and UC. A significant sex-dimorphic association was defined by the following criteria: sex-differentiated P<5 x 10-8 and a sex-heterogeneity P < 4.17 x 10-3, accounting for four tests performed in GWAMA across three traits, and a sex-specific P<5 x 10-8 in one sex (with the other sex P>0.001). To ensure observed sex differences were not due to false associations, associations driven by a single variant and variants with

P<0.01 in a test of female versus male non-IBD controls were excluded. COJO was used to assess if the sex-dimorphic variants demonstrated conditional genome-wide significance when accounting for the effect of the 619 independent association signals.

#### UK BioBank

Phenotype definitions

We used the three sources of available clinical data to define the IBD CD and UC cases within the UK Biobank:

- Main and secondary cause of admission from hospital in-patient data, ascertained using ICD-9 and ICD-10 codes
- Self-reported ‘non-cancer illness’ data, collected at the assessment centre, when the participant was recruited
- Primary care dataset, ascertained using Read 2 and CTV3/Read 3 codes

A subset of the participants (N = 774) had in-patient ICD records for both CD and UC. In those instances, the diagnosis at the last admission prevailed, 125 were reclassified as CD, 614 as UC, and the remaining 35, with CD and UC record at last admission were excluded from the CD or UC analyses but kept in the IBD analysis.

In addition, we used the prescribed data available from primary care or self-reported, to identify IBD patients among the UK Biobank participants with an IBD-specific prescription. This included drugs such as vedolizumab, mesalazine, olsalazine and balsalazide, which according to NICE guidelines are uniquely prescribed to IBD patients, and which could help ascertain additional IBD cases. Individuals with a record of these prescriptions but not otherwise identified as CD or UC patients (N = 459) were included as IBD cases.

Finally, controls were defined as UK Biobank participants, not identified as IBD cases in the datasets described above, and with no records of ‘other noninfective gastroenteritis and colitis’ in the primary care, hospital in-patient or in the primary care data. This code, although believed to include a reasonable proportion of IBD-unclassified cases, is not specific enough to reliably identify this subset of patients.

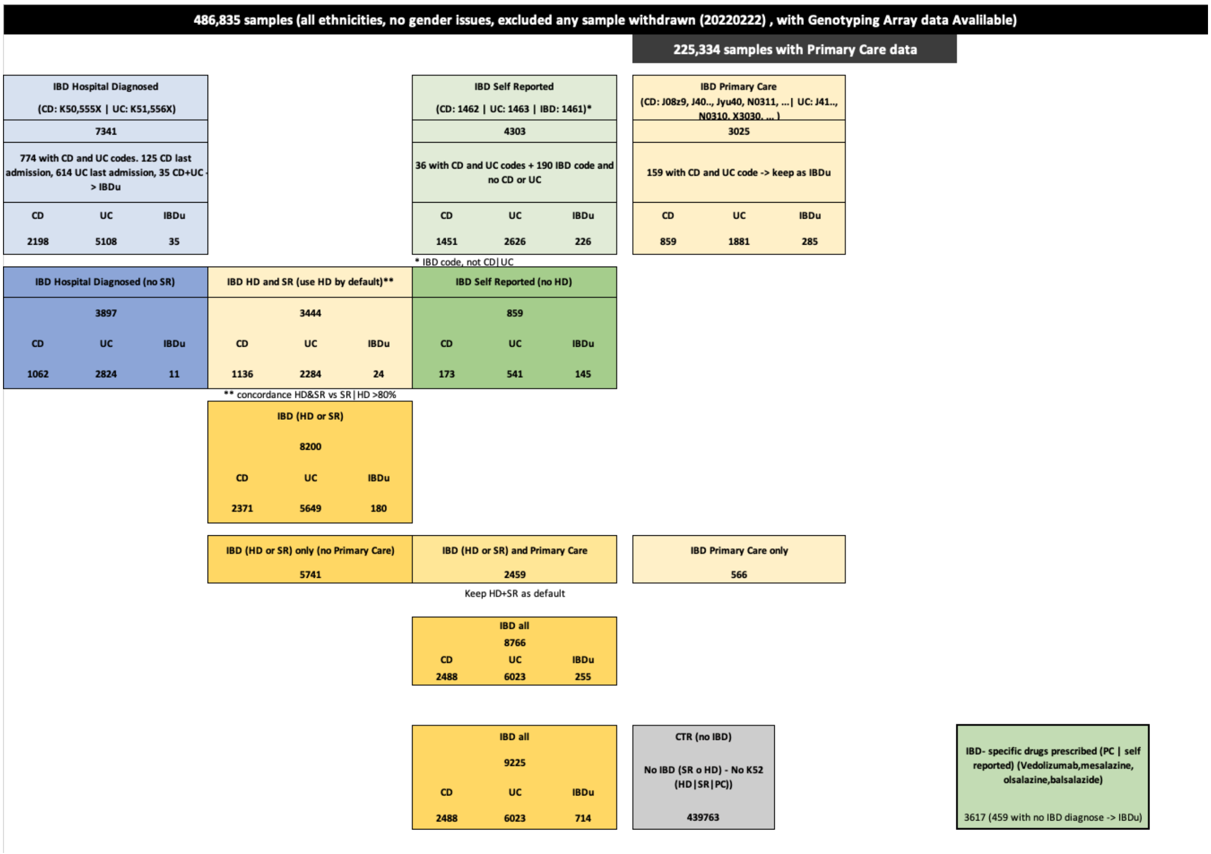


**Figure 1.1 | Algorithm to define IBD CD and UC cases and controls in UKB**

#### Evaluating the UKB case control definitions

To evaluate whether we could integrate these definitions, we derived a Genomic Risk Score (GRS) using the index variant effect sizes from[8] following the same criteria as in ref [9].

Mean GRS per disease group (CD and UC) and for the different categories was estimated for the following groups:

- HD only: cases identified in hospital in-patient data but no in the self-reported
- HD and SR: cases identified in hospital in-patient and in the self-reported dataset
- SR only: cases identified in the self-reported dataset but not in the hospital in-patient data
- Primary Care: samples identified in the primary care dataset only
- Meds: samples only identified through the IBD-specific drug prescription

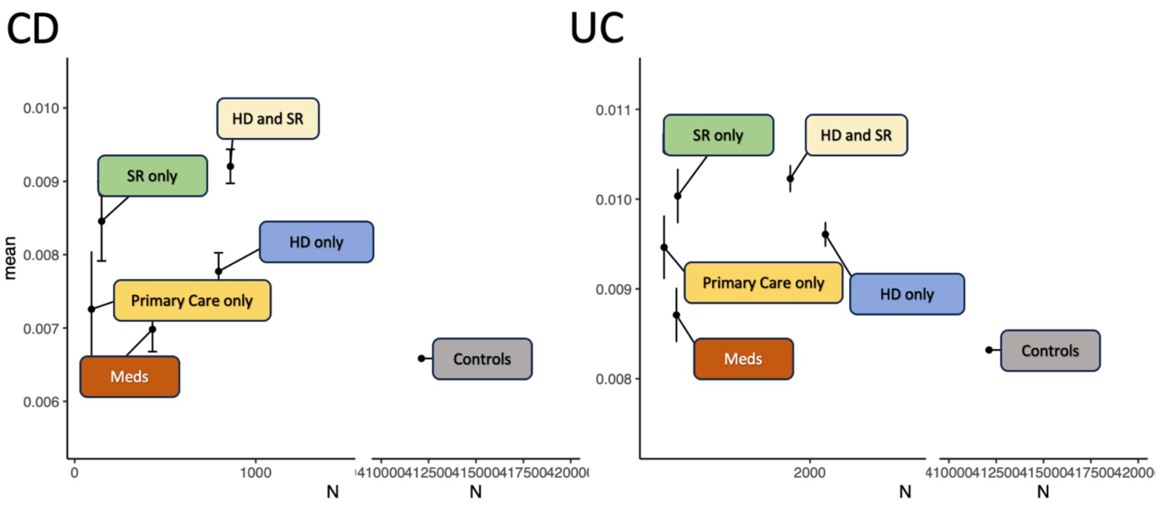


**Figure 1.2 | GRS mean per category. Mean (dot) and 95% distribution (bars) per category, versus number of UK Biobank participants in each category.**

Based on these results, CD and UC cases identified through the different clinical and prescription datasets, were combined.

##### UK Biobank Genotyping QC

Genotyping array data from the UK Biobank dataset underwent a similar QC as the IIBDGC cohorts, retaining variants by the same call rate and HWE thresholds. Samples were grouped into continental populations using the same approach as with the IIBDGC cohort. Population ancestry PCs for the analysis were re-estimated for each continental group (EUR and SAS). UK Biobank duplicated samples (N = 179, kinship ≥ 0.345), as well as participants already included within the IIBDGC dataset (kinship ≥ 0.345) and their 1st degree relatives in UKBB (kinship ≥0.177) were excluded from any further analyses.

The analysis was carried out in the UK Biobank Research Analysis Platform (RAP) with Regenie (v3.1.1; Swiss Army Knife v4.9.1) using a list of > 410,000 post-QC variants in step 1; and the UK BioBank data imputed to TOPMed in step 2 of the analyses. We included as covariates per-continental population ancestry PCs, genotyping array, and sex. Variants with HWE P≤ 10-5 in controls were also excluded from further analyses.

#### UK IBD BioResource

The cohort was genotyped with two different versions of the UKBiobank ThermoFisher genotyping array[10].

IBD BioResource duplicated samples, as well as samples from participants already included within the IIBDGC or the UK Biobank datasets (kinship ≥ 0.345) and their 1st degree relatives (kinship ≥0.177) were excluded from any further analyses.

This dataset underwent the same QC steps as the IIBDGC cohort, with the only difference that, pre-imputation, allele frequency deviation was compared to 1000GP instead. PostQC, imputation was carried out at the Sanger Imputation Server[11], using separately two reference panels, UK10K+1000GP and HRC. The imputed data from both panels was combined, so that for the set of variants present in both panels, only the data from HRC was retained. On top, we excluded a set of ~0.2% of all imputed variants showing a high discrepancy in their frequency (post imputation) between both panels (using the criterion ((p1 − p0)^2/((p1+p0)*(2-p1-p0)) > 0.00625); as well as variants with strong deviation from HWE (PHWE ≤ 10-5 in controls).

#### FinnGen summary statistics

FinnGen summary statistics (r10; more information in FinnGen webpage) were included in this study. The summary statistics included data from 2,033 CD cases and 409,940 population controls (CD_strict2 definition); 5,931 UC cases and 405,386 population controls (UC_strict2 definition); and 9,083 IBD cases and 392,974 population controls (KELAIBD definition).

#### deCODE summary statistics

49,708 samples from Icelandic participants were whole genome sequenced at deCODE using Illumina standard TruSeq methods to a mean depth of 38[12,13]. Only samples with a genome-wide average coverage of 20X and higher were used. Genotypes of single nucleotide polymorphisms (SNPs) and insertions/deletions (indels) were identified and called jointly by Graphtyper[14]. 166,281 samples from Icelandic participants had been chip-genotyped using various Illumina SNP arrays. Sample with less than 98% genotype yield, and variants with less than 95% yield and excess heterozygosity were excluded. The chip-typed individuals were long-range phased[15], and the variants identified in the whole-genome sequences of Icelanders imputed into the chip-typed individuals. Using extensive and encrypted Icelandic genealogy data, familial imputation of genotypes in first- and second-degree relatives was used to increase sample size[12,15]. The final dataset used included 24,505,450 variants with imputation information over 0.8 and minor allele frequency (MAF) over 0.1%.

Genome-wide associations were performed using software developed at deCODE Genetics, using logistic regression assuming an additive model[12]. For the Icelandic data, the model included sex, county of birth, current age or age at death (first- and second-order terms included), blood sample availability for the individual, sequencing status, and an indicator function for the overlap of the lifetime of the individual with the time span of phenotype collection. To include imputed but ungenotyped individuals in Iceland, we used county of birth as a proxy covariate for the first principal component (PC) because county of birth has been shown to be in concordance with the first PC in Iceland[16].

##### Dan-IBD summary statistics

An external Danish IBD cohort genotyped with GSA was also included in this study. Summary statistics were generated including 1,653 CD cases, 1,603 UC cases, 3,433 IBD cases, and 1,628 population controls. The dataset was imputed with TOPMed, and association testing was performed with SAIGE. The analyses included sex and population ancestry PCs as covariates.

##### EUR Tier 1 and Tier 2 meta-analysis

We used METAL (version released 2011-03-25) to perform a fixed-effects (standard error weighted) meta-analysis including the EUR summary stats from IIBDGC, UK-Biobank, IBD-Bioresource, FinnGen, deCODE and Dan-IBD. Thus, in total, this analysis included data from 1,352,042 individuals, 111,182 IBD participants (50,123 diagnosed with CD; 48,195 with UC) and 1,240,860 population controls. We assessed evidence for heterogeneity in the effects sizes by Cochran's Q statistic and the derived p-values (HetPval). We used that statistic as well to evaluate that none of the studies meta-analysed was introducing a systematic bias in the effect size estimates by using a leave-one-out approach, where the HetPval obtained when leaving one study out were compared to the HetPvals observed when all studies were combined.

##### SAS tier 2 meta-analysis

We used METAL, as described above, to perform a fixed-effects meta-analysis with the SAS summary statistics from the analysis of IBD BioResource and UK Biobank. In total we combined data from 1,215 IBD participants (338 diagnosed with CD; 613 with UC), and 6,992 population controls.

#### Trans-ancestry meta-analysis

We combined the summary statistics from the SAS and EUR cohorts together with the EAS IBD summary stats released in Liu et al. [17].These analyses included 125,992 IBD cases and 1,262,229 population controls, from three different population ancestries, EUR, EAS and SAS.

For this we followed two approaches. First, we carried out a fixed-effects meta-analysis via METAL (as described above); but we also ran a random effects meta-analysis as implemented in MR-MEGA (v0.1.5)[18]. Multidimensional scaling implemented in MR-MEGA derived the main PCs that defined the axis of genetic variation, revealing that PC1 was along sufficient to discriminate between the expected global population ancestries included in the meta-analysis (EUR, EAS, SAS; see Fig. 1.3).

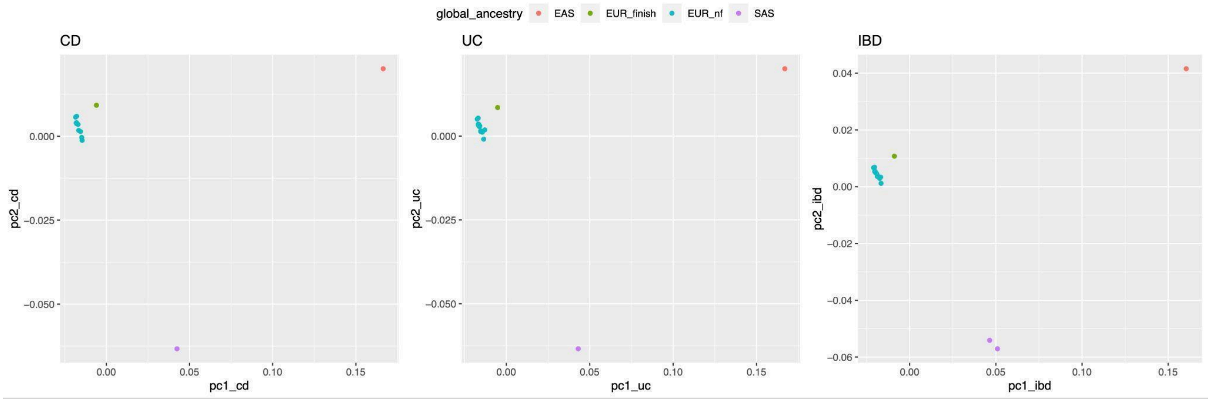


**Figure 1.3 | Two first PCs inferred by multidimensional scaling implemented in MR-MEGA**

#### Supplementary Note 2. Sex specific results

None of the 619 independent IBD signals demonstrated heterogeneity of effect between sexes for CD, UC or IBD (Supplementary Table 3). However, across the genome we observed 7 significant sex-dimorphic associations (4 female-specific and 3 male-specific), including a potentially causal novel IBD variant (Table1; chr9:114881082:C:T, intergenic to *TNFSF15*) that demonstrated a female-specific association with CD. No new IBD-associated loci were identified; the 7 observed sex-dimorphic associations all reside within genomic regions captured by the 619 independent IBD signals (Fig 2.1). Sex-dimorphic associations did not remain significant (conditional P<5 x 10-8) after accounting for the effect of the independent signals, with the exception of chr19:46640496:C:T with UC (female-specific conditional P 6.6 x10-9; OR=0.88).

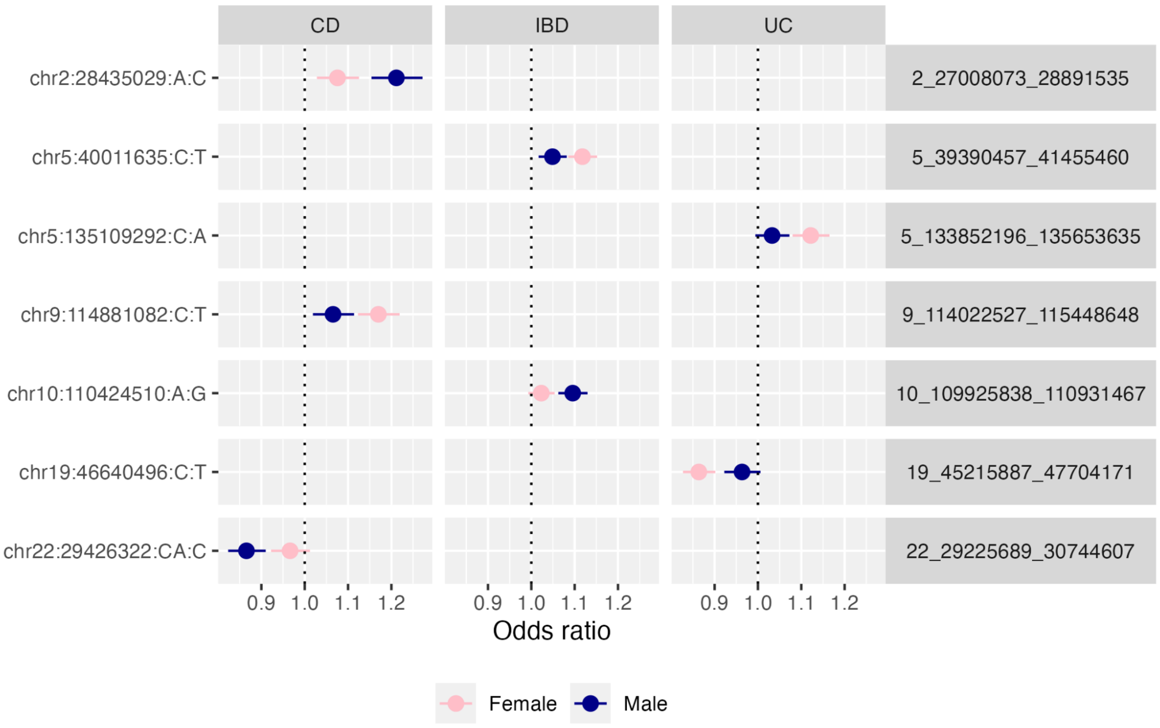


**Figure 2.1 | Sex-specific analyses per trait**

X-axis shows odds ratio for sex-dimorphic associations per trait, residing within genomic regions (right) corresponding to the independent IBD signals

### Supplementary Note 3. Heritability analyses informed by molecular QTL datasets

To refine colocalization-based candidates, cell-type enrichment Z-scores were calculated for each QTL dataset, and for each signal-gene pair we assigned the maximum cell-type Z-score observed across all colocalizing QTLs (PP H4) for that gene. For each eQTL or pQTL dataset (one condition at a time), we computed enrichment Z-scores using S-LDSC[19], annotating the lead index variant significantly associated with gene expression or plasma protein levels and extending this annotation to variants in LD with the index variant. To assess robustness to LD definition, we evaluated the stability of the Z-score estimates at different LD thresholds ($r^2$ = 0.25, 0.5, 0.75) in a representative dataset (Macromap). Z-scores were largely consistent across thresholds (Figure 1 a, c), but the statistical significance of the estimates increased systematically as the threshold decreased (Figure 1 b). On this basis, we selected an LD threshold of $r^2$ = 0.25 for all subsequent analyses.

As an external calibration, we included height as an independent trait[20]. eQTLs from cell types identified as disease-relevant in our analyses, such as macrophages (Fig 3.1a), exhibited higher enrichment Z-scores for IBD subtypes relative to height (Figure 3.1b, c). However, the enrichment specificity (IBD vs. height) based on eQTLs alone was lower than for other omics modalities (ATAC-seq or DNase), with several tissues and cell types showing significant enrichment for both height and IBD-related traits.

Among signals with multiple nominated effector genes, rankings based on tissue enrichment and coding-burden heritability showed significant concordance, indicating convergence between these independent prioritization approaches and providing further support to this strategy.

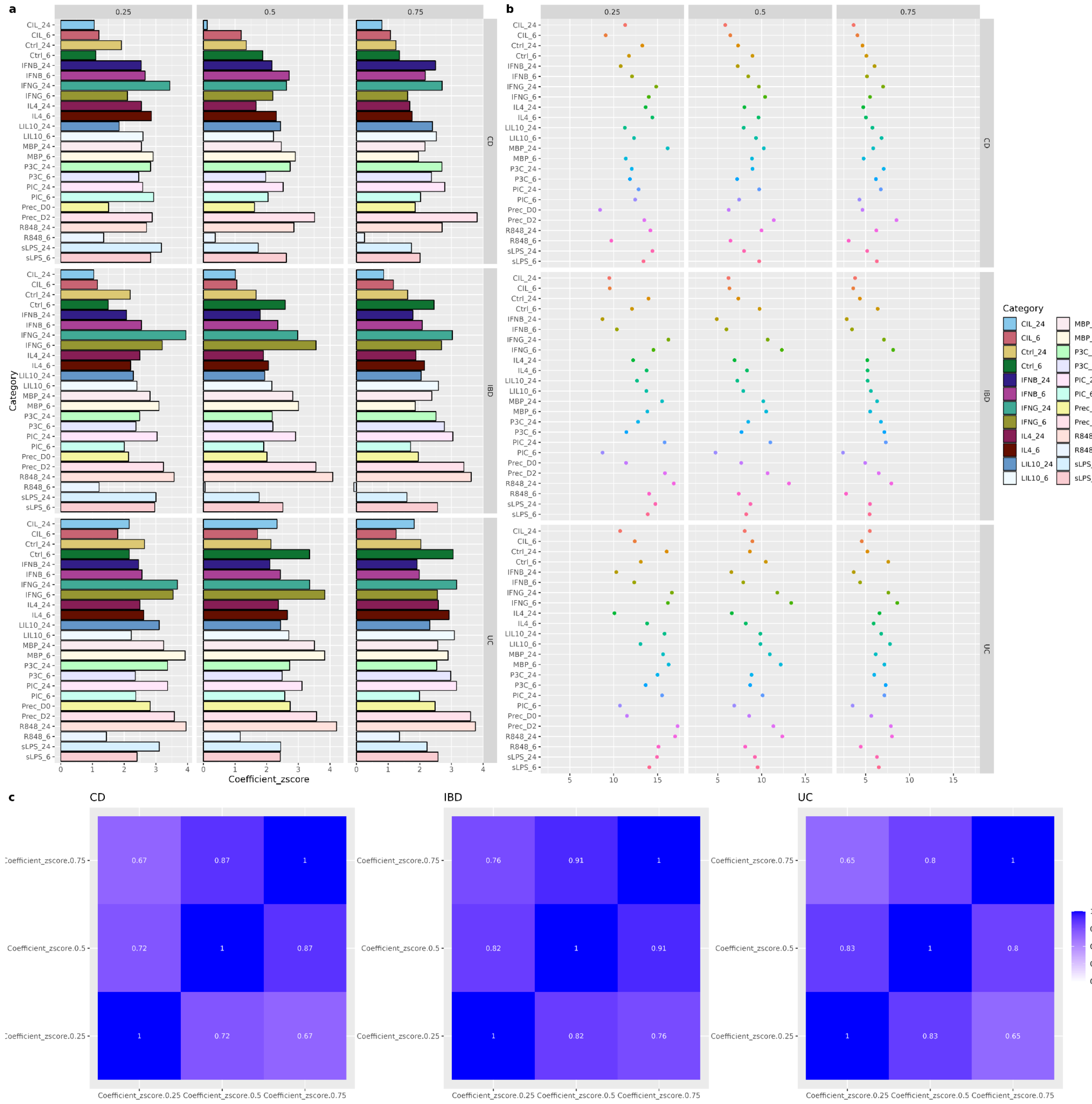


**Figure 3.1 | Heritability analyses at different LD thresholds to define informed weights per cell type, condition and IBD subtype**

We tested the performance of different LD thresholds to build the eQTL and pQTL annotations. The plots show the impact of using LD r2 threshold on 0.25, 0.5 and 0.75, using data from Macromap as reference.

a) on the z-score estimations; and (b) on the p-values.  c) correlation estimates between the z-score estimates at different LD thresholds threshold for each IBD phenotype.

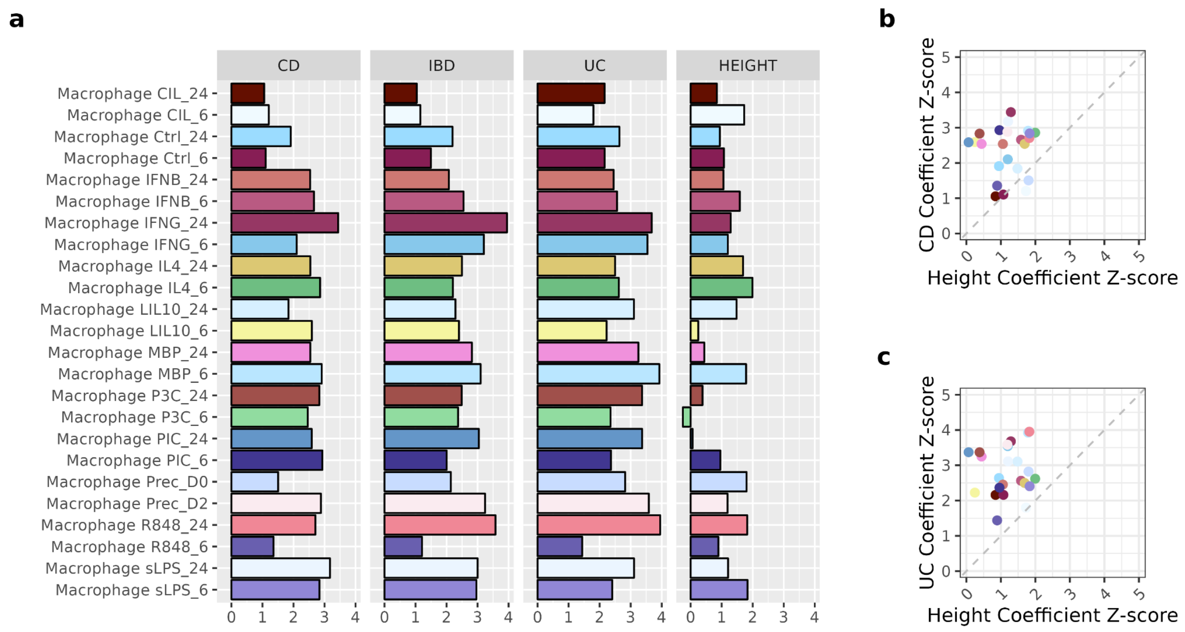


**Figure 3.2 | Heritability analyses to define informed weights per cell type, condition and IBD subtype**

a, eQTL enrichment z scores; b,c, z-score comparisons between IBD subtypes (CD and UC) and height

### Supplementary Note 4. Independent signals characterization

We assessed whether characteristics of the independent signals (N = 619) differed according to whether effector genes were successfully assigned. Signal characteristics were derived from the lead variant for each signal. Of the 619 signals, 341 were assigned effector genes. Signals without nominated genes had significantly lower minor allele frequencies than those with nominated genes, particularly for colocalization-based assignments (Fig. 1a). Although all signals had high imputation quality (INFO score > 0.6), those without nominated genes showed significantly lower imputation INFO scores.

No significant differences were observed for the effective sample size contributing to the analysis (Neff; Fig. 4.1c), distance to the transcription start site of the closest gene (TSS; Fig. 4.1d), or effect size (Fig. 4.1e).

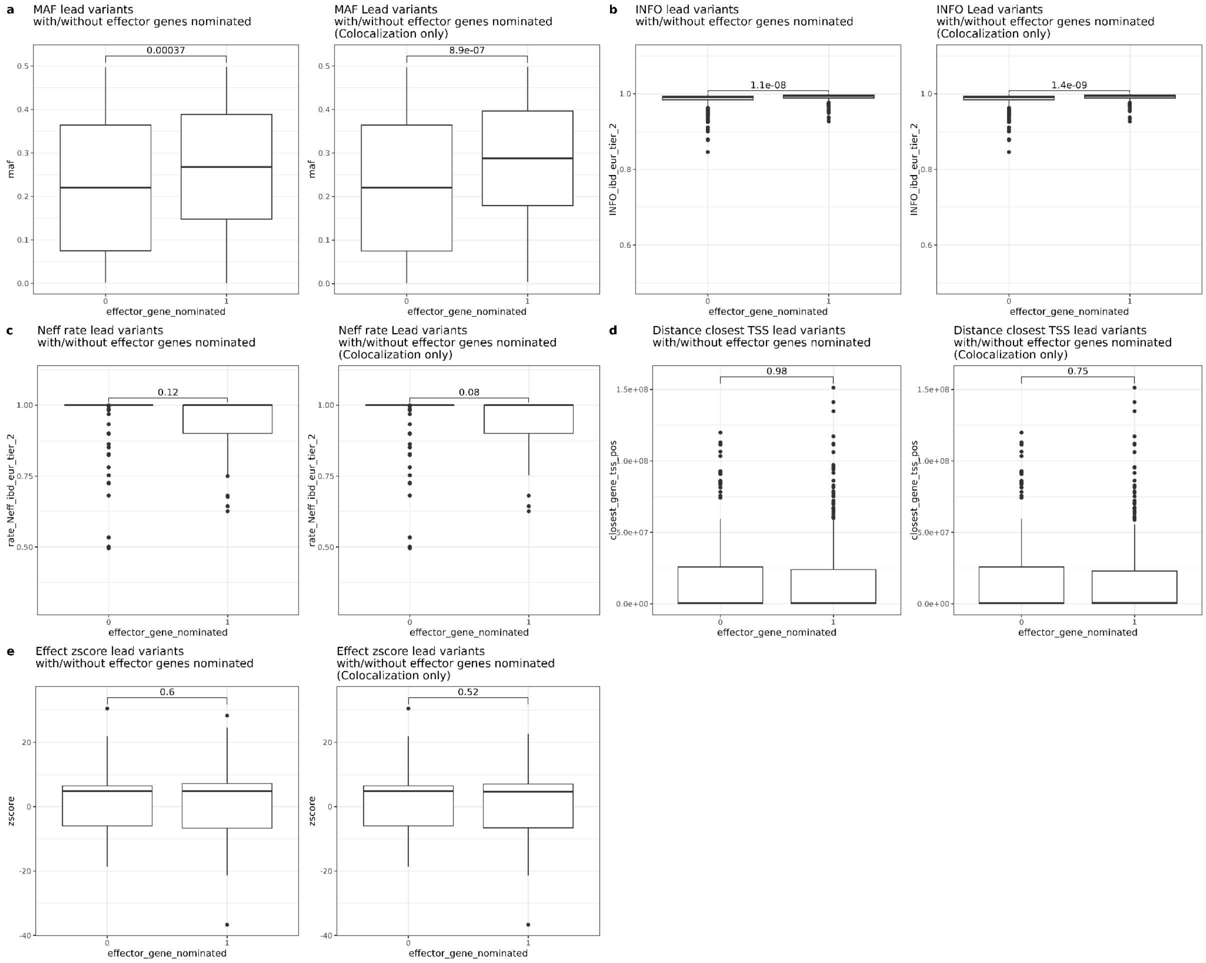


**Figure 4.1 | Characterization of the independent signals.**

Y axis categorises the signal with regard to whether an effector gene could be linked to the signal (0 = No; 1 = Yes). Y axis a: Minor allele frequency of the lead variant; b: imputation INFO score; c: N effective rate; d: distance to closest transcription start site (TSS); e: IBD risk Z score (beta/se). All tests were done using a Wilcoxon rank sum test, reported P values are from two-sided tests.

### Supplementary Note 5. Heritability captured per lead variant and power

We estimated the proportion of heritability (on the liability scale, as in https://cloufield.github.io/gwaslab/PerSNPh2/) explained by each independent lead variant (Figure 5.1a,b). We also estimated the sample size required to detect variants at different heritability thresholds (Figure 1c).

To detect variants explaining variances < 1.75x10$^{-4}$ we would need considerably larger sample sizes than our current study (Neff IBD = 240386; Neff CD = 118922; Neff UC 145099).

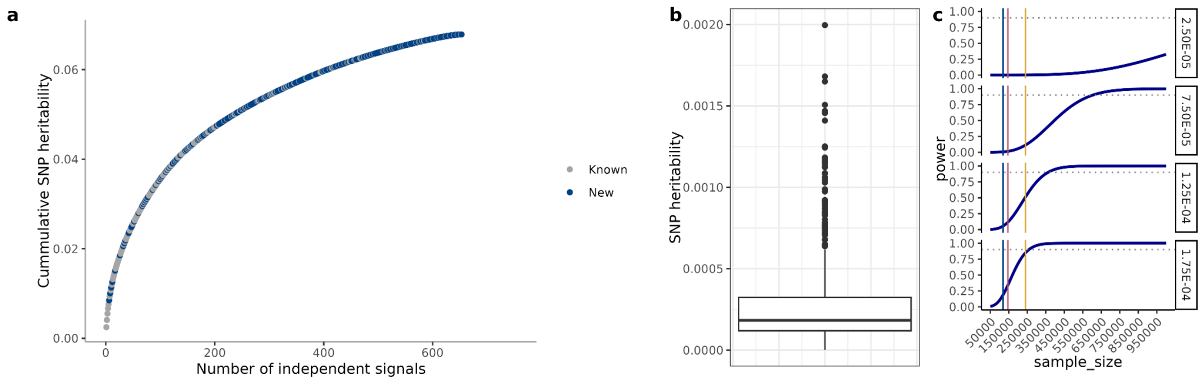


**Figure 5.1. Variance explained per lead variant**

a, proportion of the heritability explained by the independent signals lead variants (N = 621); b. Distribution of genetic variance per lead variant; c, estimates to detect genetic variants by heritability estimates (right x axis). The dotted horizontal line shows power = 0.9. Vertical lines indicate Neff of current study (yellow IBD; red UC; blue CD)

### Supplementary Note 6 - S-LDSC analyses

We applied S-LDSC[19] to identify the cell types that play a role in IBD, CD and UC disease susceptibility. Five independent functional genomics resources were included in this analysis: the immune cell atlas[21] (Fig. 6.1); the cis-elements atlas of human tissues[22] (Fig. 6.2); two single-nucleus open chromatin studies across eight different intestinal sites, Hickey et al.[23] (Fig. 6.3); a chromatin accessibility map of T cells and macrophages under different stimuli, Soskic et al.[24] (Fig. 6.4); and primary immune cell types profiled via DNase, intact Hi-C and ATAC-seq as part of the ENCODE project[25](Fig. 6.5). To assess the specificity of the selected cell types and datasets, and to validate the S-LDSC-derived annotations and weights generated for this study, we performed the same analyses using height GWAS summary statistics as a negative control (Fig. 6.6; Methods). The enrichment analyses for the variables included in the baseline model are represented in Fig 6.7.

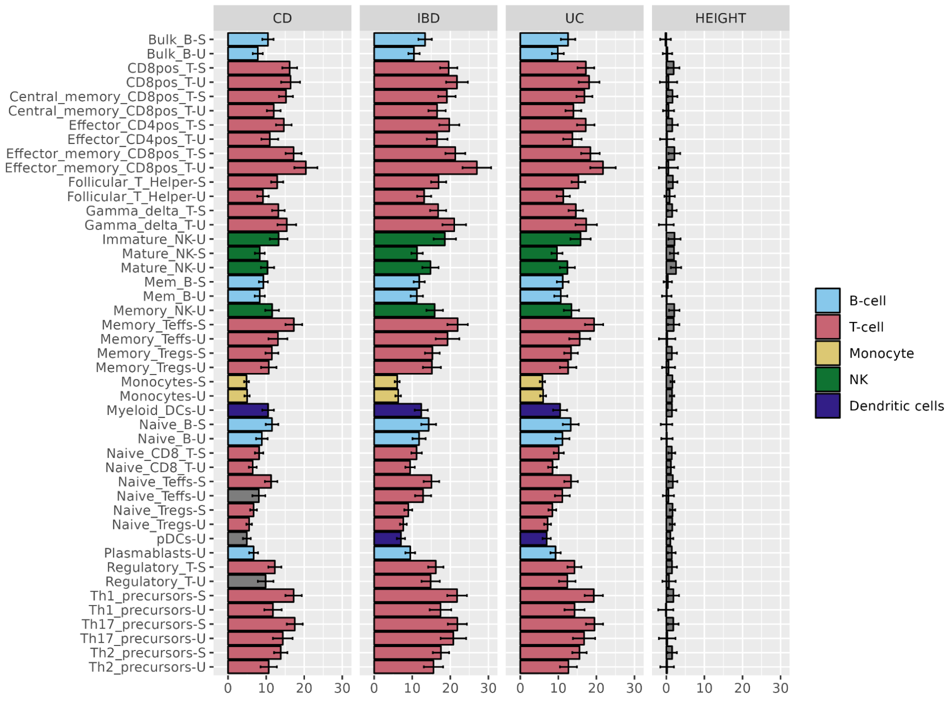


**Figure 6.1 | Enrichment analyses across the Immune Cell Atlas (Calderon et al. 2019)**[21]

The x axis shows the enrichment (Prop.h2/Prop.SNPs) per annotation, whereas the y axis indicates the cell types. Error bars represent the enrichment standard error. Cell types represented under stimulated (-S) and resting (-U) conditions). Results have been coloured by cell type, grouping different cell types in a broader category (ie, Bulk Naive and Memory B cells into B-cell). Coloured bars represent the significant results after multiple testing correction (Bonferroni correction, two sided p value distribution); in grey, non significant results

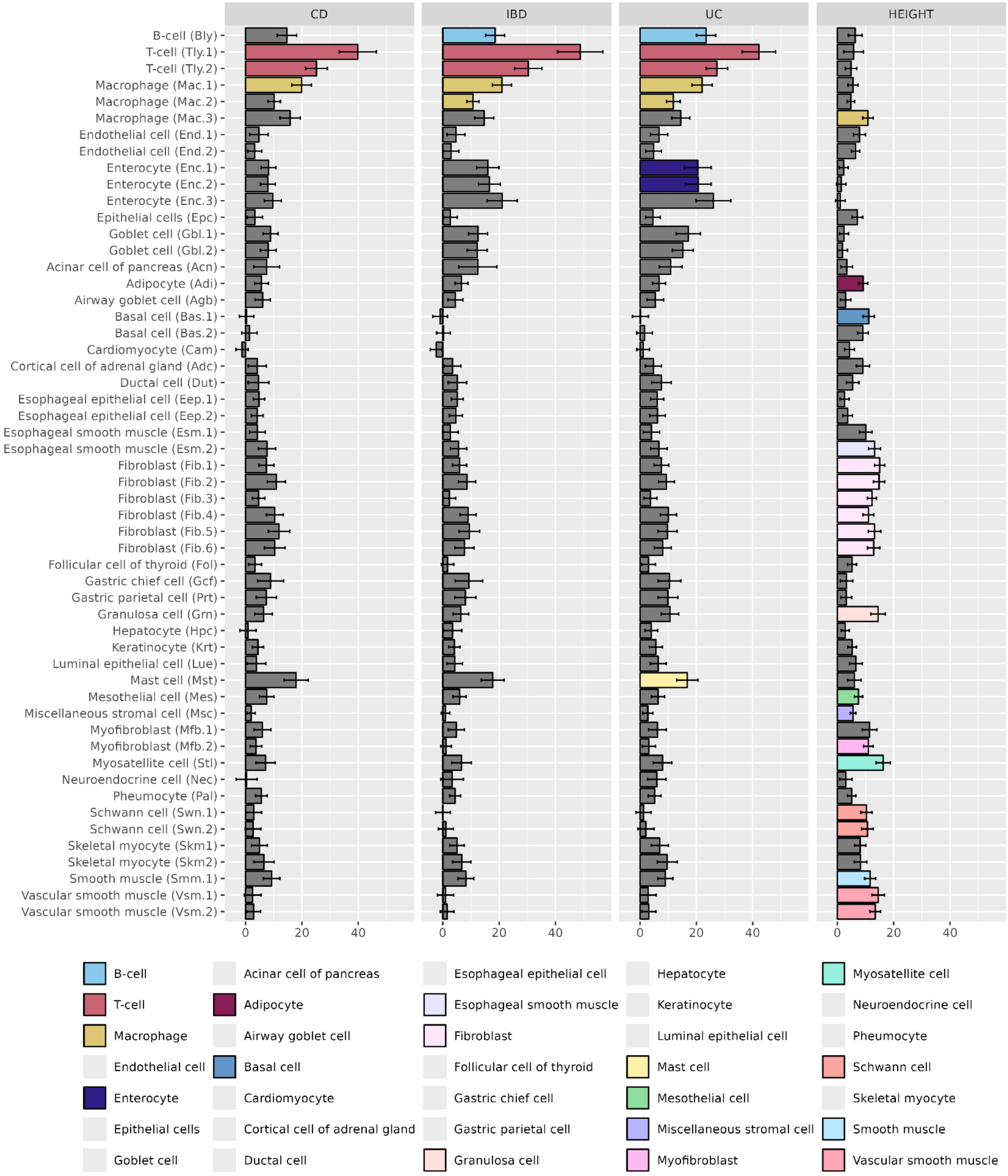


**Figure 6.2 | Enrichment analyses across the cis-element atlas (Zhang et al. 2021)[22]**

The x axis shows the enrichment (Prop.h2/Prop.SNPs) per annotation, whereas the y axis indicates the cell types. Error bars represent the enrichment standard error. Results have been coloured by cell type, grouping different cell types in a broader category (ie, Goblet cells Gbl.1 and Gbl.2 into Goblet). Coloured bars represent the significant results after multiple testing correction (Bonferroni correction, two sided p value distribution); in grey, non significant results

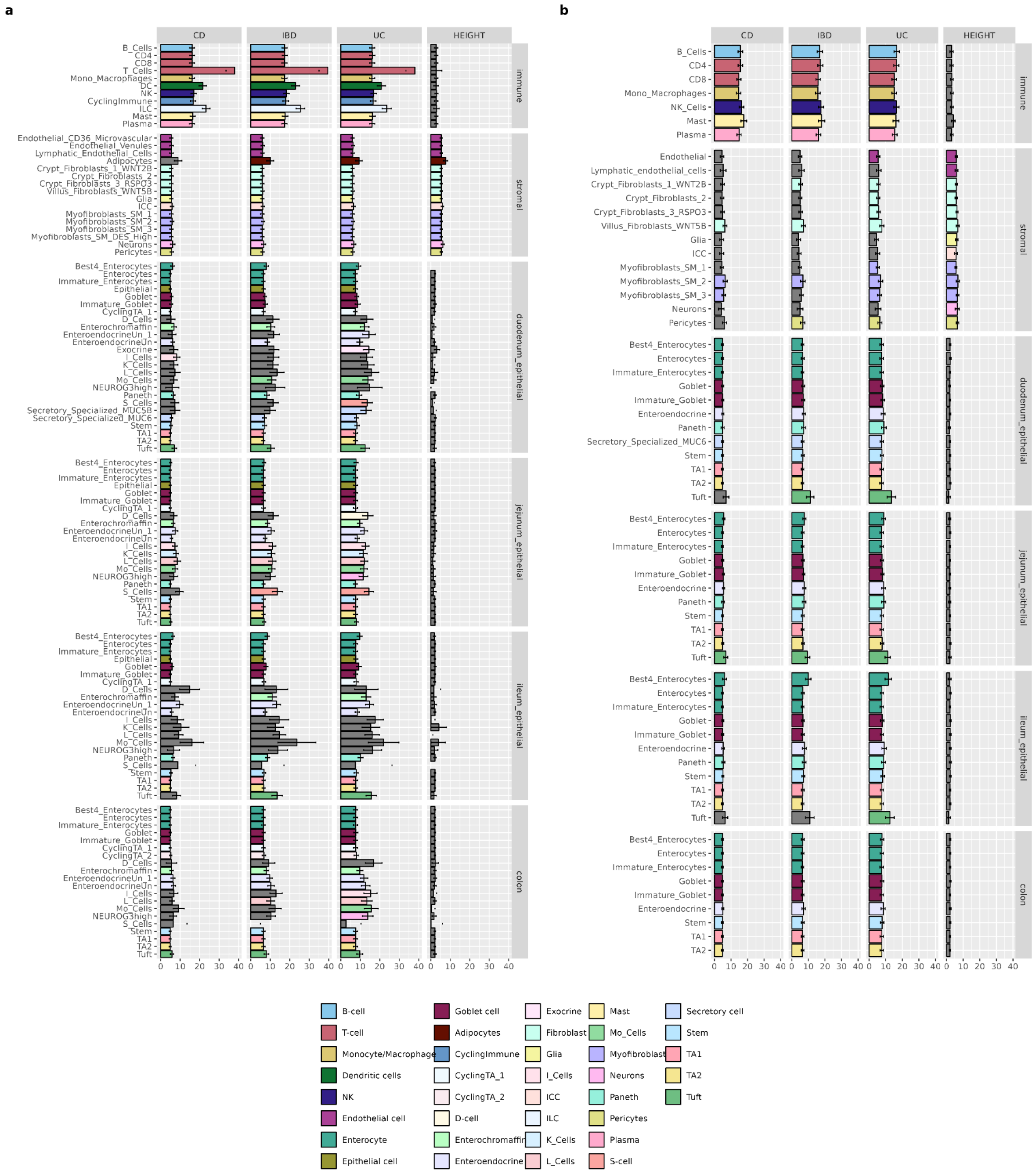


**Figure 6.3 | Enrichment analyses in intestinal cell types (Hickey et al. 2023)**[23]

Results using Hickey et al. multiome data (panel a) and non-multiome data (panel b)

The x axis shows the enrichment (Prop.h2/Prop.SNPs) per annotation, whereas the y axis indicates the cell types. Error bars represent the enrichment standard error. Results have been coloured by cell type, grouping different cell subtypes in a broader category (ie, Goblet and Immature Goblet cells into Goblet). Coloured bars represent the significant results after multiple testing correction (Bonferroni correction, two sided p value distribution); in grey, non significant results

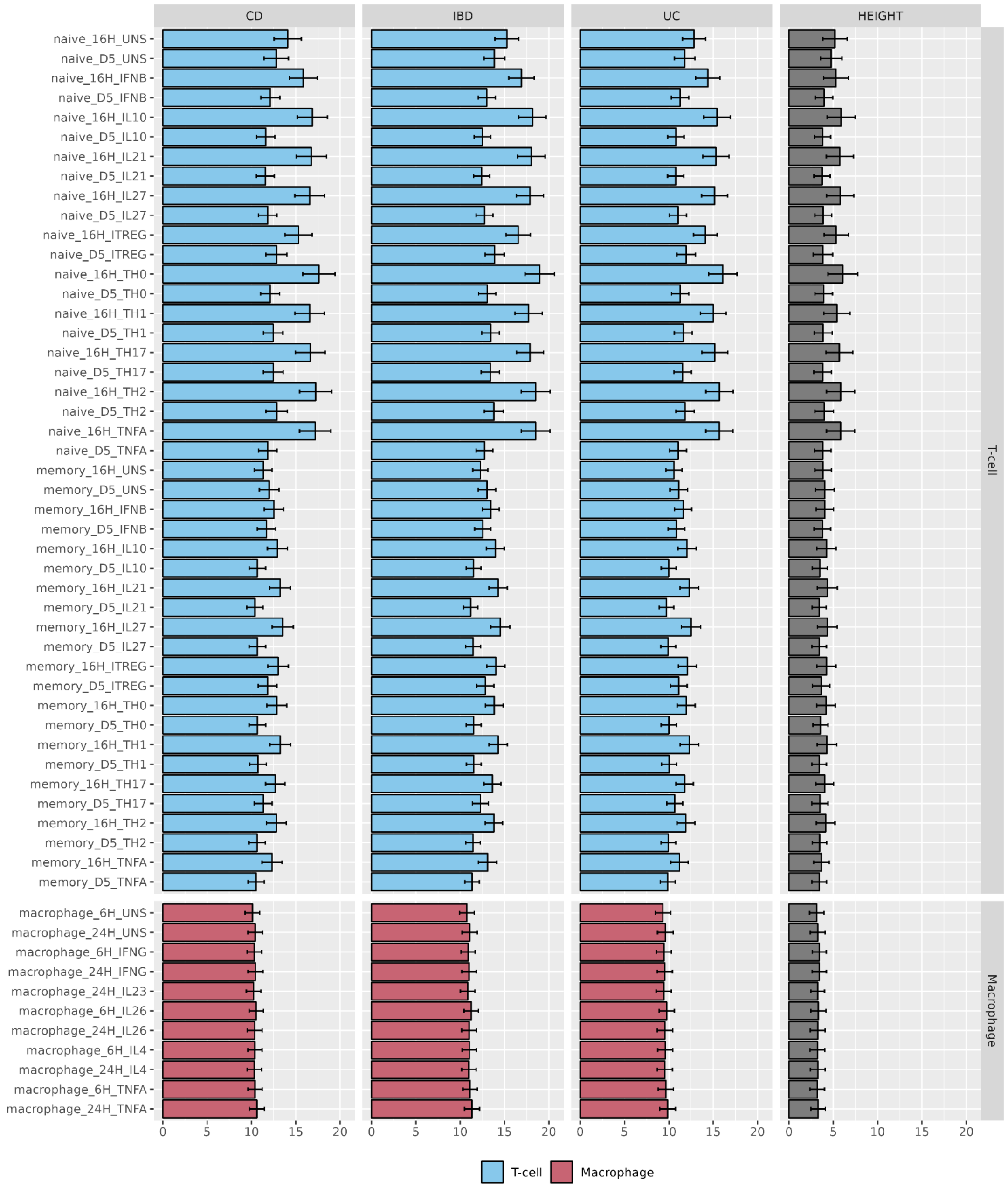


**Figure 6.4 | Enrichment analyses in T cells and Macrophages under different stimuli (Soskic et al. 2019)**[24]

The x axis shows the enrichment (Prop.h2/Prop.SNPs) per annotation, whereas the y axis indicates the cell types. Error bars represent the enrichment standard error. Results have been coloured by cell type, grouping different cell types in a broader category (ie, memory and naive T-cells into T-cells). Coloured bars represent the significant results after multiple testing correction (Bonferroni correction, two sided p value distribution); in grey, non significant results

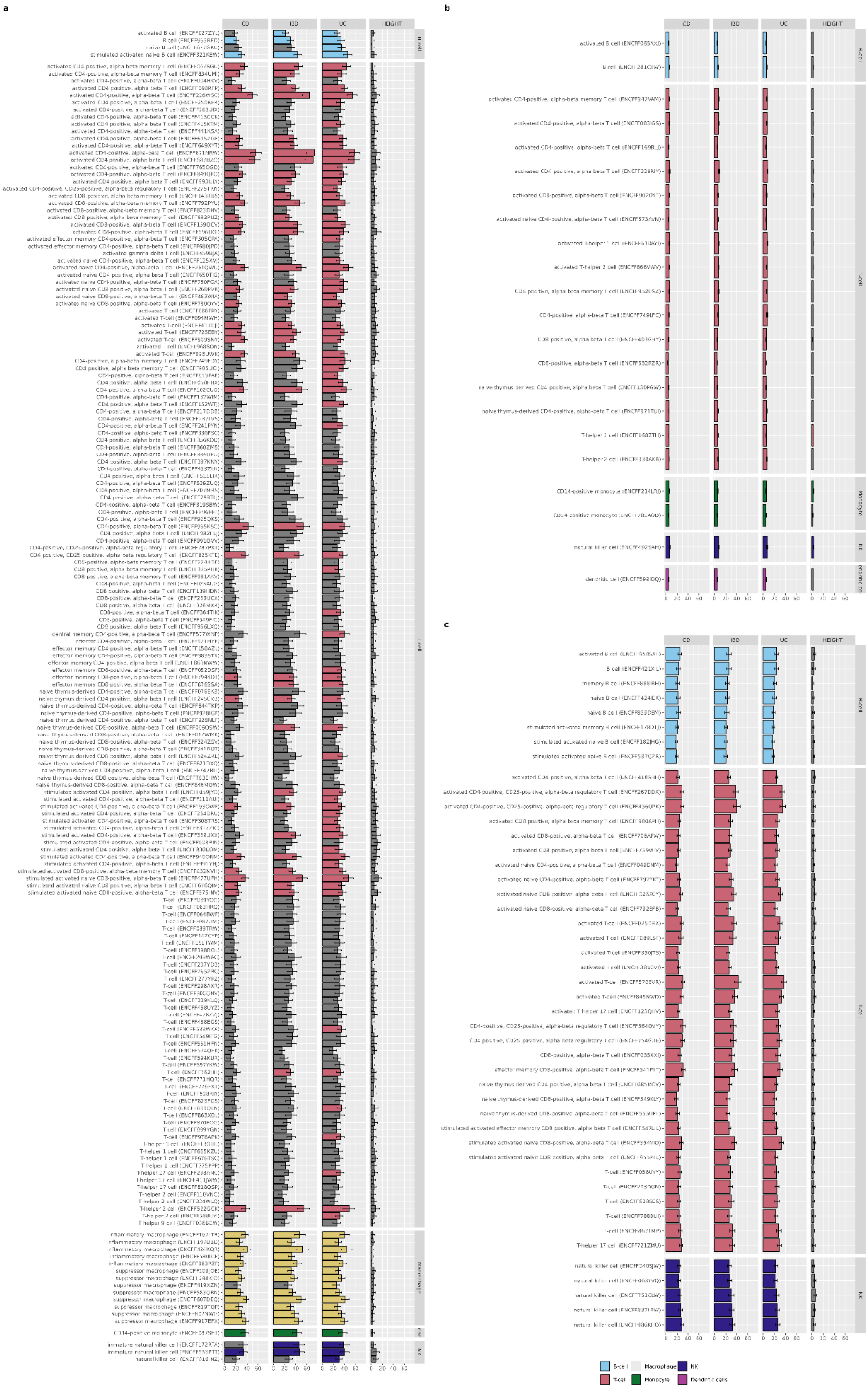

**Figure 6.5 | Enrichment analyses in immune cell types (ENCODE)**[26]

Results using ENCODE DNAse (panel a), intact HiC (panel b) and ATAC seq (panel c) data

The y axis shows the enrichment (Prop.h2/Prop.SNPs) per annotation, whereas the y axis indicates the cell types. Error bars represent the enrichment standard error. Results have been coloured by cell type, grouping different cell types in a broader category (ie, memory and naive T-cells into T-cells).

Coloured bars represent the significant results after multiple testing correction (Bonferroni correction, two sided p value distribution); in grey, non significant results

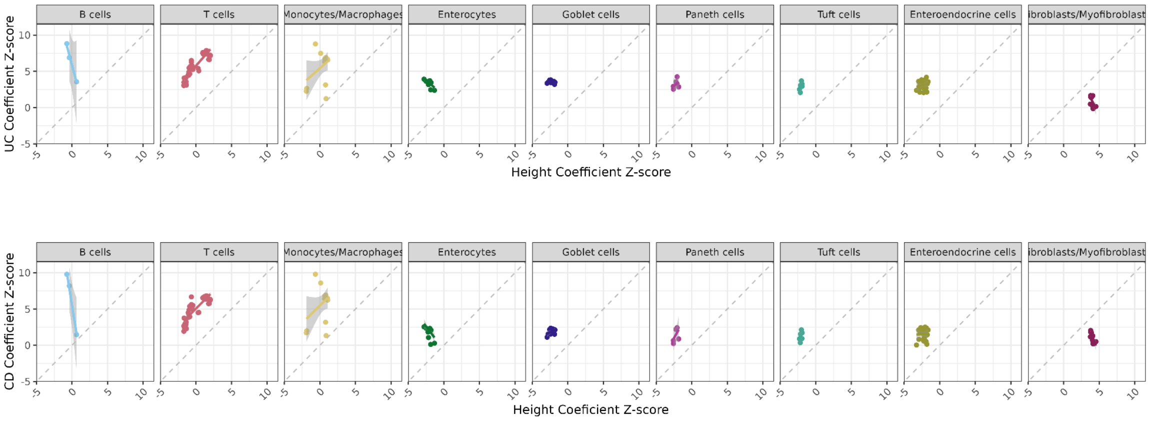


**Supplementary Fig 6.6 | Cell type enrichment analyses.**

Pair comparison between coefficient z-score across a subset of cell types from all studies. Only cell types with more than 2 independent estimates are included. The x axis shows the S-LDSC coefficient z-score for Height, whereas the y axis shows the z-score for either UC (top panel), or CD (bottom panel).

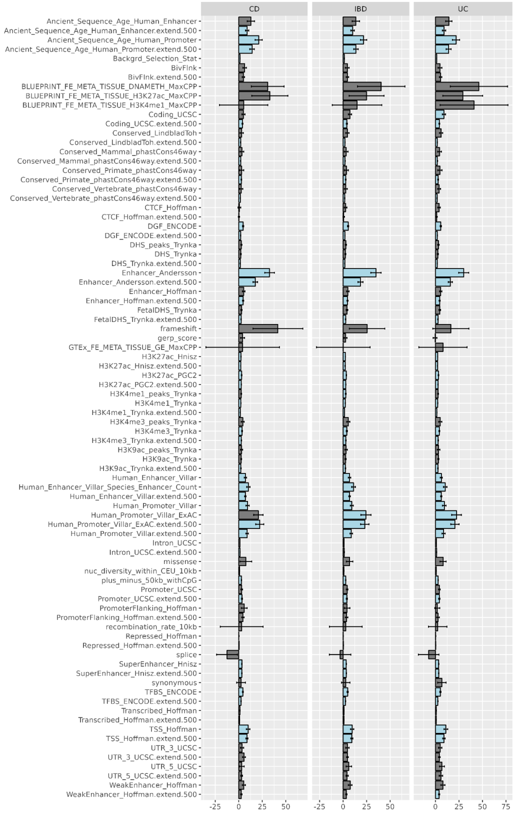


**Supplementary Fig 6.7 | Enrichment analyses using the baseline annotations**

The y axis shows the enrichment (Prop.h2/Prop.SNPs) per annotation, whereas the y axis indicates the cell types. Error bars represent the enrichment standard error. Coloured bars represent the significant results after multiple testing correction (Bonferroni correction, two sided p value distribution); in grey, non significant results; in light blue, significantly enriched annotations

### Supplementary Note 7. Explanation for replication failure of 21 high-PIP variants previous IBD fine-mapping analyses

We further assessed the replication of 45 high-PIP variants reported in Huang et al.[27] Of the 45 variants, 24 were replicated in our analysis, including 18 that remained lead variants and 6 that were retained within credible sets (Supplementary Table 13). Non-replicated variants were largely explained by differences in data availability, allele frequency, statistical significance or refined fine-mapping resolution in the current study.

We next assessed whether the independent signals identified by conditional analysis using COJO recapitulated the fine-mapped credible sets. Of the 268 credible sets, 249 were captured by conditional analysis ($r^2 > 0.1$), 216 of them in high linkage disequilibrium ($r^2 > 0.6$) (Supplementary Table 12).

Among the 21 variants that failed replication, 3 were not present in our EUR tier 1 data, 5 had a rare MAF (< 0.1%), and 8 were not genome-wide significant in our EUR tier 1 dataset (Supplementary Table 6). The detailed evaluation of the remaining five variants is provided below.

1. **rs4655215; chr1:19811221:T:C; n_signal=3, signal=3; RNF186 downstream; UC phenotype**

For this region (1_19238082_20420495), the first signal reported in Huang et al.[27] was a credible set including variants chr1:19845367:G:A (lead variant) and chr1:19839478:C:T, associated with the UC phenotype (upper right corner in the UC inset). In our new fine-mapping results, these two variants are included in the IBD credible set but not in UC. Moreover, chr1:19845367:G:A is no longer the lead variant. Instead, the new lead variant is chr1:19901230:G:C in both IBD and UC, and these two credible sets can be aligned. Therefore, the final assigned phenotype for this signal is UC.

- Variants being included in the credible set of UC: chr1:19901230:G:C
- Variants being included in the credible set of IBD: chr1:19901230:G:C, chr1:19845367:G:A, chr1:19839478:C:T, chr1:19845021:T:G

As such, the first signal reported in Huang et al.[27] is replicated in our new fine-mapping results.

For the target variant chr1:19811221:T:C, it is the third signal in this region. The LD (r) between this variant and a newly identified causal variant chr1:19805278:C:T is about 0.56. In the Huang et al. data, chr1:19811221:T:C showed a stronger association, whereas in our new data, chr1:19805278:C:T is more significant. Therefore, we consider the failure to replicate this variant to be data-driven.

2. **rs77981966; chr2:43550825:C:T; n_signal=1, signal=1; THADA, intronic; CD**

In this region (2_43050825_44050826), Huang et al. identified a single signal in CD with lead variant chr2:43550825:C:T. In our new fine-mapping results, we also identified a signal present in both IBD and CD (with CD as the final assigned phenotype), with lead variant chr2:43541406:T:G. The LD (r) between these two variants chr2:43550825:C:T and chr2:43541406:T:G is approximately 0.7. In our data, chr2:43541406:T:G shows stronger association than chr2:43550825:C:T, whereas the opposite was observed in the Huang et al. dataset, Therefore, we consider the failure to replicate the target variant chr2:43550825:C:T to be data-driven.

3. **rs28701841, chr6:106082455:G:A, n_signal=2, signal=2, PRDM1, upstream, CD**

In this region (6_104959734_106582456), Huang et al.[27] reported two signals associated with the CD phenotype. The lead variant of the first signal was chr6:106026874:C:T (not included in the list of 45 high PIP variants, PIP_Huang = 0.31), and the lead variant of the secondary signal was chr6:106082455:G:A. Neither of these two signals was reported in our new fine-mapping results. Instead, we identified three new credible sets in this locus:

- one in CD with lead variant chr6:106013960:T:C (LD r is about 0.37 with the target variant)
- one in IBD: chr6:105994221:G:C (LD r is about 0.47 with the target variant)
- one in IBD and UC: chr6:106016006:T:C (LD r is about -0.32 with the target variant)

We consider the failure to replicate the target variant chr6:106082455:G:A to be data-driven, as the newly identified credible sets in our analysis show stronger association than the previously reported signals.

4. **rs4728142; chr7:128933913:G:A; n_signal=1, signal=1; IRF5, upstream; UC**

In this region (7_128242231_129433914), both our new fine-mapping analysis and Huang et al. identified a single signal. The lead variant in the credible set reported by Huang et al.[27] was chr7:128933913:G:A, associated with the UC phenotype. In our new results, the lead variant is chr7:128941781:A:G, identified in both IBD and UC. The two credible sets are perfectly aligned, as they contain exactly the same variants, with UC assigned as the final phenotype. The variant, chr7:128941781:A:G, was also included in the credible set from Huang et al., though it was not the lead variant. The LD (r) between chr7:128933913:G:A and chr7:128941781:A:G is approximately 0.82.

Notably, the newly identified credible set is a subset of the one reported by Huang et al.[27], consisting of only three variants (compared to ten previously). The cumulative PIP of these three variants exceeds 0.95, indicating no need to include additional variants in the credible set. Moreover, these three variants show stronger association signals in our data than the other variants, which is not the case in Huang et al.[27] data. We therefore consider the failure to replicate the target variant chr7:128933913:G:A to be data-driven.

5. **rs146029108; chr9:136435514:GTTAT:G; n_signal=3, signal=3; INPP5E, intronic; CD**

In this region (9_135865140_137434173), Huang et al.[27] identified three signals:

- The first credible set, with lead variant chr9:136378050:T:C, associated with the IBD phenotype
- The second credible set, with lead variant chr9:136365140:C:G, associated with IBD
- The third credible set, with lead variant chr9:136435514:GTTAT:G, associated with CD; this is also the target variant in this evaluation

Compared to our new fine-mapping results:

- The lead variant of the first signal (chr9:136378050:T:C) is included in the credible sets for CD and UC, with the lead variant chr9:136369439:C:G (the LD r between these two variants is 0.92), which is also the lead variant of a credible set for IBD. These three credible sets are aligned, and the final assigned phenotype is IBD. Thus, the variant chr9:136378050:T:C can be replicated in our current results.
- The lead variant of the second signal (chr9:136365140:C:G) is a splicing variant in the gene CARD9 and is also identified as the lead variant in the UC credible set in our new fine-mapping results.

In total, we identified four credible sets in this region:

- One with lead variant chr9:136365140:C:G in UC phenotype, corresponding to the second signal in Huang et al.
- One with lead variant chr9:136533458:C:T in IBD
- One with lead variant chr9:136369439:C:G in IBD, CD, and UC, with IBD as the final assigned phenotype, corresponding to the first signal in Huang et al.
- One with lead variant chr9:136928090:C:T in IBD and CD, with CD as the final assigned phenotype (not shown in figure, as it is not present in the ImmunoChip data of Huang et al.)

In our EUR tier1 data, the P value for the newly identified CD lead variant chr9:136928090:C:T is $6.45 \times 10^{-10}$, while the P value of target variant chr9:136435514:GTTAT:G is $4.89 \times 10^{-13}$. Since the first and second signals reported by Huang et al. are replicated in our results, and the P value of the newly identified lead variant is comparable to that of the lead variant of the third signal reported by Huang et al.[27], we consider the failure to replicate the third signal to be algorithm- driven.

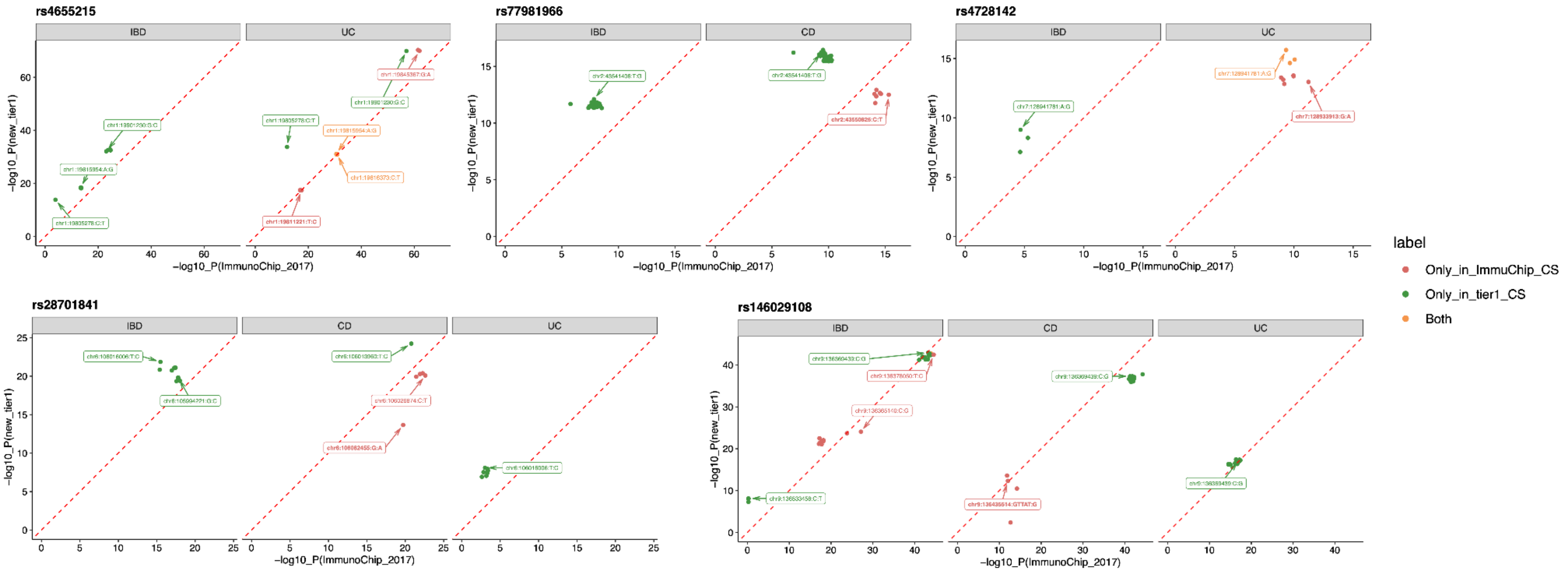


**Figure 7.1 | Explanation of replication failure for five variants**

### Supplementary Note 8. Replication of previous GWAS signals

We were unable to replicate previously reported associations at 47 known IBD regions. Five reached genome-wide significance in the Multiancestry analyses, but were not identified as independent signals with COJO using EUR TIer 2 summary statistics. Of the remaining 42 signals, 21 were first reported in Liu et al.[17] (Figure); a multi-ancestry meta-analysis combining European and East Asian cohorts. Among the remaining 21 regions identified prior to Liu et al.[17], only 4 had previously achieved genome-wide significance in previous IIBDGC analyses [8]. Three of these (led by chr1:209797265:G:A, chr10:124750812:G:A and chr20:6113242:T:G) reached $P < 1\times10^{-4}$ in our multi-ancestry meta-analysis, whereas one signal (led by chr10:26890667:T:G) could not be replicated.

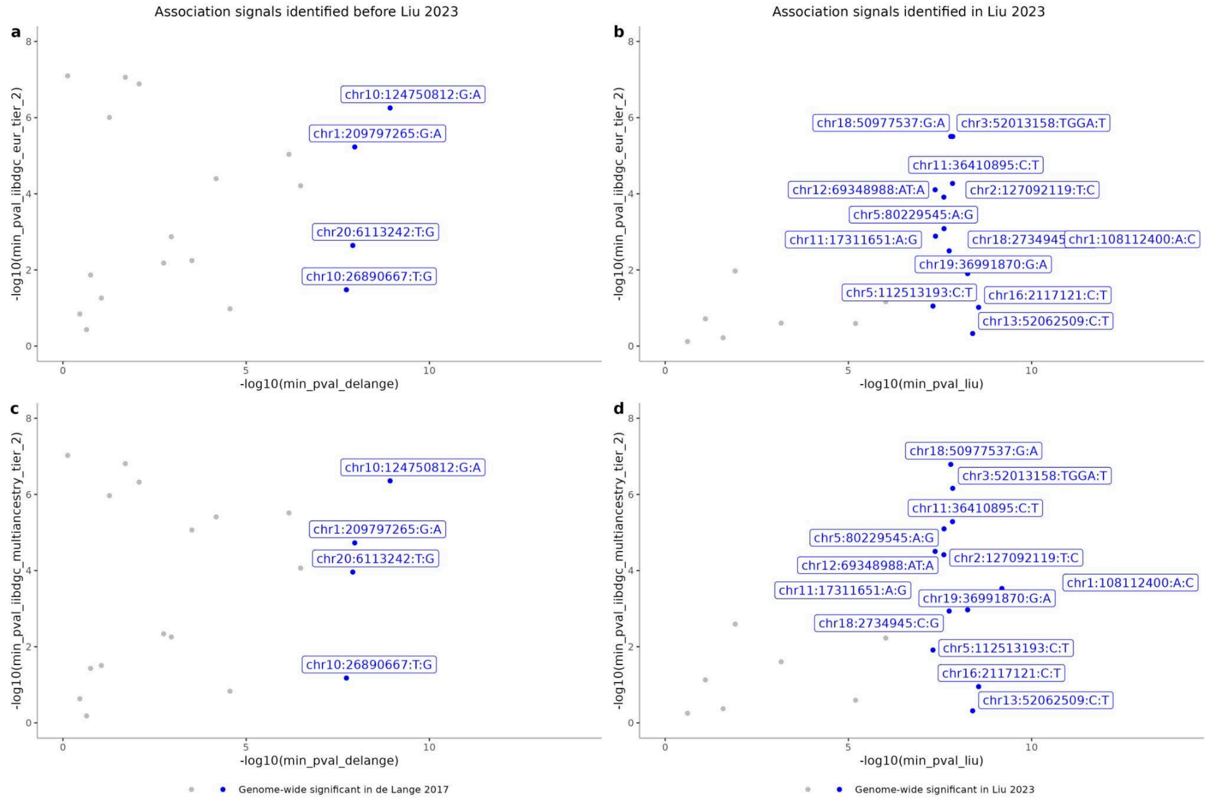


**Figure 8.1 | Non replicated SNPs**

The plot represents the most significant P(CD, UC or IBD) for 42 index variants, at 42 IBD regions, not replicated in the current study.

X axis (a, c) p-values in de Lange et al.[8]; (b, d) Liu et al.[17] Y axis (a, b) P in the EUR Tier 2 meta-analyses; (c, d) Multiancestry Tier 2 (EUR, EAS, SAS) meta-analyses. SNPs with a label indicate those that reach genome-wide significance levels in the previous study.

#### Supplementary Note 9. Acknowledgements

We thank all of the principal investigators, local staff from individual cohorts, and all of the patients who kindly donated samples used in the study for making this global collaboration and resource possible to advance IBD genetics research. We thank Tobi Alegbe, Bradley Harris for helpful discussions. We thank the Wellcome Sanger Institute's Human Genetics Informatics (HGI) and Scientific Operations teams for their support, and Kavyaa Venkat for administrative support. We wish to thank our funders: The National Institute for Health and Care Research Centres in Cambridge (NIHR203312), Newcastle, University College London and Exeter; the Medical Research Council, the Wellcome Trust, Open Targets, Crohn's & Colitis UK, the Helmsley Charitable Trust, Crohn's in Childhood Research Association (CICRA) and our Industry partners. We thank NIHR BioResource volunteers for their participation, and gratefully acknowledge NIHR BioResource centres, NHS Trusts and staff for their contribution. We thank the National Institute for Health and Care Research, NHS Blood and Transplant, and Health Data Research UK as part of the Digital Innovation Hub Programme. The views expressed are those of the authors and not necessarily those of the NIHR or the Department of Health and Social Care. This research was funded in whole, or in part, by the Wellcome Trust [Grant numbers 206194 and 220540/Z/20/A]. We would like to thank the clinical researchers and patients participating in the IMID Consortium for their collaboration. This work was supported by Health Data Research UK, which is funded by the UK Medical Research Council, Engineering and Physical Sciences Research Council, Economic and Social Research Council, Department of Health and Social Care (England), Chief Scientist Office of the Scottish Government Health and Social Care Directorates, Health and Social Care Research and Development Division (Welsh Government), Public Health Agency (Northern Ireland), British Heart Foundation and Wellcome. This research has been conducted using the UK Biobank Resource under Application Number 45669. Slovenian study was supported by Slovenian Research and Innovation Agency research (core funding P3-0427 and grant no. I0-0029) and by the Republic of Slovenia Ministry of Higher Education, Science, and Innovation and the European Union through the European Regional Development Fund (grant RIUM). The authors would like to thank the patients for participating. The results published here are in part from the Study of a Prospective Adult Research Cohort with IBD (SPARC IBD), a component of the Crohn's & Colitis Foundation IBD Plexus program. Support by NIH Washington University DDRCC Grant No. NIDDK P30

DK052574 We want to acknowledge all investigators in the SNP-IBD and BIO-IBD programs for their contribution to the recruitment of patients.

Lundbeckfonden R24620171, Health Research Fund of Central Denmark Region, The A.P. Møller Foundation, Colitis-Crohn Foreningen, Aase og Ejnar Danielsens Fond, Beckett-Fonden, Novo Nordisk Fonden NNF15OC0016932. The Swiss IBD Cohort Study (SIBDCS) is supported by Swiss National Science Foundation grant # 3347CO-108792/1. Samples were provided by the Inflammatory Bowel in South Eastern Norway (IBSEN) study group and the Norwegian Bone Marrow Donor Registry (NMBDR). The study received infrastructure support from the Deutsche Forschungsgemeinschaft (DFG, German Research Foundation) Cluster of Excellence 2167 “Precision Medicine in Chronic Inflammation (PMI)” (EXC 2167-390884018) and acknowledges the EU IMI programm ("3TR"), and the European Regional Development Fund 01.2.2-LMT-K-718-04-0003. The study received infrastructure support from the Deutsche Forschungsgemeinschaft (DFG, German Research Foundation) Cluster of Excellence 2167 “Precision Medicine in Chronic Inflammation (PMI)” (EXC 2167-390884018). Our current work is supported by funding to Jacob L. McCauley and Maria T. Abreu through the NIH/NIDDK (U01DK134201). Italian Minister of Health, Ricerca Corrente program 2022–2024, Division of Gastroenterology, Fondazione IRCCS “Casa Sollievo della Sofferenza” Hospital, San Giovanni Rotondo (Italy); “5 × 1000” voluntary contribution. The study received infrastructure support from the Deutsche Forschungsgemeinschaft (DFG, German Research Foundation) Cluster of Excellence 2167 “Precision Medicine in Chronic Inflammation (PMI)” (EXC 2167-390884018). This work was supported by internal funds from the F. Widjaja Foundation Inflammatory Bowel Disease Institute; NIH/NIDDK U01 DK062413; and the Leona M and Harry B Helmsley Charitable Trust. To Be Confirmed with Dr. Silverberg. We wish to acknowledge all the participants who took part in the study and the clinicians, clinical nurses, administrative staff and research nurses who assisted in the study. The IMAGINE Network is supported by Canadian Institutes for Health Research (CIHR)'s Strategy for Patient Oriented Research (SPOR) (Funding Reference Number: 1715-000-001) with funding from several other partners. We are grateful for the help with biobanking in Leuven provided by Vera Ballet and Justien Degry; and we thank Helene Blevi, Tamara Coopmans, Arno Cuvry, and Sophie Organe for processing the Leuven patient samples.

We want to acknowledge the participants and investigators of the FinnGen study. The FinnGen project is funded by two grants from Business Finland (HUS 4685/31/2016 and UH 4386/31/2016) and the following industry partners: AbbVie Inc., Alnylam Pharmaceuticals, Inc., AstraZeneca UK Ltd, Bayer AG, Biogen MA Inc., Boehringer Ingelheim International

GmbH, Bristol Myers Squibb Inc. (and Celgene Corporation & Celgene International II Sàrl), Genentech Inc., GlaxoSmithKline Intellectual Property Development Ltd., Johnson&Johnson Innovative Medicine Inc., Maze Therapeutics Inc., Merck Sharp & Dohme LCC, Novartis AG, Pfizer Inc. and Sanofi US Services Inc. Following biobanks are acknowledged for delivering biobank samples to FinnGen: Auria Biobank (www.auria.fi/biopankki), THL Biobank (www.thl.fi/biobank), Helsinki Biobank (www.helsinginbiopankki.fi), Biobank Borealis of Northern Finland (https://www.ppshp.fi/Tutkimus-ja-opetus/Biopankki/Pages/Biobank-Borealis-briefly-in-English.aspx), Finnish Clinical Biobank Tampere (www.tays.fi/en-US/Research_and_development/Finnish_Clinical_Biobank_Tampere), Biobank of Eastern Finland (www.ita-suomenbiopankki.fi/en), Central Finland Biobank (www.ksshp.fi/fi-FI/Potilaalle/Biopankki), Finnish Red Cross Blood Service Biobank (www.veripalvelu.fi/verenluovutus/biopankkitoiminta), Terveystalo Biobank (www.terveystalo.com/fi/Yritystietoa/Terveystalo-Biopankki/Biopankki/) and Arctic Biobank (https://www.oulu.fi/en/university/faculties-and-units/faculty-medicine/northern-finland-birth-cohorts-and-arctic-biobank). All Finnish Biobanks are members of BBMRI.fi infrastructure (https://www.bbmri-eric.eu/national-nodes/finland/). Finnish Biobank Cooperative -FINBB (https://finbb.fi/) is the coordinator of BBMRI-ERIC operations in Finland. The Finnish biobank data can be accessed through the Fingenious® services (https://site.fingenious.fi/en/) managed by FINBB.

#### Supplementary Note 10. IIBDGC patient-facing lay summary

Inflammatory bowel disease is a long-term condition characterised by inflammation in the bowel. The two main types are Crohn's disease and ulcerative colitis. Crohn's disease can affect any part of the digestive tract, while ulcerative colitis affects the large bowel. People develop inflammatory bowel disease for many reasons, including processes in the immune system, things in the environment, chance, and inherited factors. For many years, scientists and clinicians have worked to understand how inherited factors may increase or decrease the risk of the disease.

This study was carried out by a large international group of scientists and clinicians working together to combine knowledge gained from previous research into the inherited risk of inflammatory bowel disease. This is important because no single hospital, university or country could have done research on this scale alone. By bringing together a large body of genetic information from many earlier studies, the team was able to use DNA information from more than 125,000 people living with inflammatory bowel disease and more than 1.2 million people without it. The researchers could then compare these two groups, looking for DNA markers that were more common in people with inflammatory bowel disease.

As a result, this research has greatly expanded the genetic map of inflammatory bowel disease. It has doubled the number of known regions of DNA linked with the condition. A key step was to compare this new genetic map with existing information about gene activity, proteins, and different types of cell. This enabled the researchers to explore what the DNA findings might reveal about what may be happening in the bodies of people with inflammatory bowel disease.

The findings draw attention to the importance of the bowel lining, especially in people with ulcerative colitis. This matters because many current treatments for inflammatory bowel disease work by calming or suppressing parts of the immune system. Better understanding of how inherited factors may affect the bowel lining could help researchers explore parts of the disease process beyond the immune system alone.

The researchers also compared the genetic findings from inflammatory bowel disease with those seen in people with other long-term conditions. Some common genetic links were already known, especially with conditions where the immune system plays a role, such as coeliac disease and arthritis. The researchers also found other connections that were more unexpected, including links with metabolism. This new insight may help researchers understand more about how the body and the environment interact in inflammatory bowel disease.

This research does not mean that a person can take a genetic test to tell whether they will or will not develop Crohn's disease or ulcerative colitis. Nor does it change what treatments are currently available. However, these new findings do mark a major step forward in understanding the genetic map of inflammatory bowel disease. The team estimates that this study can now explain over 80% of the inherited risk measured by this type of genetic research. This gives scientists and doctors a much clearer understanding of how inherited factors contribute to the development of inflammatory bowel disease, and will help guide future work towards better treatments.

### Supplementary Note 11. Lead variants showing significant ancestry-related heterogeneity in effect size, as estimated by MR-MEGA

As part of the QC process to evaluate the independent signals, we interrogated the meta-analysis heterogeneity P values from the fixed-effects analyses and those derived by MR-MEGA as ancestry-related. Those lead variants showcasing a high ancestry related heterogeneity are likely reflecting a failure to capture the causal variant as the lead variant; and instead capturing tag variants with different LD patterns with the causal variant across ancestries. This is exemplified by the known IBD signal at the TNRC18 locus[28], and supported by FinnGen fine-mapping results. The lead variant chr7:5397122:C:T is excluded from most EUR studies due to its low frequency in non-Finnish Europeans (gnomAD EUR MAF ~ 0.001 versus Finnish MAF ~ 0.02). This causes an alternative variant tagging chr7:5433979:G:C to be selected by COJO instead.

| MarkerName | Multiancestry HetPval | Multiancestry HetPval (ancestry related) | SAS A2 Freq | EAS A2 Freq | EUR A2 Freq | Finnish A2 Freq |
|---|---|---|---|---|---|---|
| **chr7:5433979:G:C** | **1.84E-16** | **4.52E-38** | 0.1189 | 0.5350 | 0.0255 | 0.0468 |
| **chr9:114801407:T:G** | 1.47E-02 | **1.49E-36** | 0.7585 | 0.6426 | 0.7165 | 0.7387 |
| **chr1:67214256:C:CT** | 4.44E-02 | **9.87E-16** | 0.5423 | 0.5443 | 0.2764 | 0.2438 |
| **chr4:38333446:G:A** | 1.57E-01 | **2.32E-10** | 0.2407 | 0.2058 | 0.0639 | 0.0380 |
| **chr9:114851353:GA:G** | 4.50E-01 | **5.32E-08** | 0.0292 | 0.0334 | 0.0192 | 0.0186 |
| **chr13:26957130:T:C** | 3.00E-01 | **1.30E-07** | 0.2836 | 0.2277 | 0.1778 | 0.1801 |
| **chr9:136395559:G:A** | 3.07E-03 | **3.48E-07** | 0.3254 | 0.3031 | 0.4150 | 0.4051 |
| **chr9:114845889:A:G** | 8.41E-01 | **4.24E-06** | 0.2860 | 0.0238 | 0.2823 | 0.2751 |
| **chr5:159402286:C:T** | 1.18E-04 | **4.72E-06** | 0.3656 | 0.3592 | 0.3162 | 0.2993 |
| **chr10:62710915:C:T** | 1.69E-01 | **5.16E-06** | 0.2726 | 0.2656 | 0.1802 | 0.2161 |
| **chr16:50500888:G:T** | 7.33E-01 | **7.70E-06** | NA | NA | 0.0060 | 0.0069 |
| **chr7:27192143:C:T** | 5.34E-01 | **8.14E-06** | 0.8236 | 0.5820 | 0.8042 | 0.7919 |
| **chr5:59021173:A:G** | 4.94E-01 | **1.06E-05** | 0.0958 | 0.4682 | 0.0817 | 0.1011 |
| **chr10:110425838:G:A** | 4.21E-03 | **1.18E-05** | 0.1976 | 0.1696 | 0.3524 | 0.3613 |
| **chr12:40337211:A:G** | 5.26E-01 | **1.35E-05** | 0.1048 | 0.0437 | 0.0202 | 0.0251 |
| **chr5:40498420:T:TG** | 1.13E-01 | **1.45E-05** | 0.4235 | 0.4872 | 0.6475 | 0.5950 |
| **chr19:1127616:G:C** | 1.19E-01 | **3.41E-05** | 0.2561 | 0.2117 | 0.2170 | 0.2323 |
| **chr17:42345821:G:C** | 6.66E-01 | **4.04E-05** | 0.3764 | 0.3374 | 0.2767 | 0.2818 |
| **chr1:161547427:G:A** | 4.04E-01 | **4.48E-05** | 0.0920 | 0.1880 | 0.0856 | 0.0629 |

| | | | | | | |
|---|---|---|---|---|---|---|
| **chr5:150872803:G:A** | 1.27E-01 | **4.58E-05** | 0.2064 | 0.4188 | 0.0801 | 0.0840 |

**Table 12.1 | List of variants demonstrating a significant ancestry-related heterogeneity in effect size, as estimated by MR-Mega**

References

1. Purcell, S. *et al.* PLINK: a tool set for whole-genome association and population-based linkage analyses. *Am J Hum Genet* **81**, 559–575 (2007).
2. Danecek, P. *et al.* Twelve years of SAMtools and BCFtools. *Gigascience* **10**, (2021).
3. Manichaikul, A. *et al.* Robust relationship inference in genome-wide association studies. *Bioinformatics* **26**, 2867–2873 (2010).
4. Das, S. *et al.* Next-generation genotype imputation service and methods. *Nat Genet* **48**, 1284–1287 (2016).
5. Mbatchou, J. *et al.* Computationally efficient whole-genome regression for quantitative and binary traits. *Nat Genet* **53**, 1097–1103 (2021).
6. Magi, R., Lindgren, C. M. & Morris, A. P. Meta-analysis of sex-specific genome-wide association studies. *Genet Epidemiol* **34**, 846–853 (2010).
7. Yang, J. *et al.* Conditional and joint multiple-SNP analysis of GWAS summary statistics identifies additional variants influencing complex traits. *Nat Genet* **44**, 369–75, S1–3 (2012).
8. de Lange, K. M. *et al.* Genome-wide association study implicates immune activation of multiple integrin genes in inflammatory bowel disease. *Nat Genet* **49**, 256–261 (2017).
9. Serra, E. G. *et al.* Author Correction: Somatic mosaicism and common genetic variation contribute to the risk of very-early-onset inflammatory bowel disease. *Nat Commun* **13**, 3576 (2022).
10. Parkes, M. & IBD BioResource Investigators. IBD BioResource: an open-access platform of 25 000 patients to accelerate research in Crohn's and Colitis. *Gut* **68**, 1537–1540 (2019).
11. McCarthy, S. *et al.* A reference panel of 64,976 haplotypes for genotype imputation. *Nat Genet* **48**, 1279–1283 (2016).
12. Gudbjartsson, D. F. *et al.* Large-scale whole-genome sequencing of the Icelandic

population. *Nat Genet* **47**, 435–444 (2015).

13. Jónsson, H. *et al.* Whole genome characterization of sequence diversity of 15,220 Icelanders. *Scientific Data* **4**, 170115 (2017).
14. Eggertsson, H. P. *et al.* Graphtyper enables population-scale genotyping using pangenome graphs. *Nat Genet* **49**, 1654–1660 (2017).
15. Kong, A. *et al.* Detection of sharing by descent, long-range phasing and haplotype imputation. *Nat Genet* **40**, 1068–1075 (2008).
16. Price, A. L. *et al.* The impact of divergence time on the nature of population structure: an example from Iceland. *PLoS Genet* **5**, e1000505 (2009).
17. Liu, Z. *et al.* Genetic architecture of the inflammatory bowel diseases across East Asian and European ancestries. *Nat Genet* **55**, 796–806 (2023).
18. Mägi, R. *et al.* Trans-ethnic meta-regression of genome-wide association studies accounting for ancestry increases power for discovery and improves fine-mapping resolution. *Hum Mol Genet* **26**, 3639–3650 (2017).
19. Finucane, H. K. *et al.* Partitioning heritability by functional annotation using genome-wide association summary statistics. *Nat Genet* **47**, 1228–1235 (2015).
20. Sakaue, S. *et al.* A cross-population atlas of genetic associations for 220 human phenotypes. *Nat Genet* **53**, 1415–1424 (2021).
21. Calderon, D. *et al.* Landscape of stimulation-responsive chromatin across diverse human immune cells. *Nat Genet* **51**, 1494–1505 (2019).
22. Zhang, K. *et al.* A single-cell atlas of chromatin accessibility in the human genome. *Cell* **184**, 5985–6001.e19 (2021).
23. Hickey, J. W. *et al.* Organization of the human intestine at single-cell resolution. *Nature* **619**, 572–584 (2023).
24. Soskic, B. *et al.* Chromatin activity at GWAS loci identifies T cell states driving complex immune diseases. *Nat Genet* **51**, 1486–1493 (2019).
25. ENCODE Project Consortium. An integrated encyclopedia of DNA elements in the

human genome. *Nature* **489**, 57–74 (2012).

26. ENCODE Project Consortium. An integrated encyclopedia of DNA elements in the human genome. *Nature* **489**, 57–74 (2012).
27. Huang, H. *et al.* Fine-mapping inflammatory bowel disease loci to single-variant resolution. *Nature* **547**, 173–178 (2017).
28. Rahimov, F. *et al.* A genome-wide CRISPR screen identifies the TNRC18 gene locus as a regulator of inflammatory signaling. *Nat Commun* **16**, 10346 (2025).
